# Adjusting for residual confounding using high-dimensional propensity scores in a study of inhaled corticosteroids and COVID-19 outcomes

**DOI:** 10.1101/2025.02.04.25321459

**Authors:** Marleen Bokern, John Tazare, Christopher T. Rentsch, Jennifer K. Quint, Ian Douglas, Anna Schultze

**Author notes:** Corresponding author; Marleen Bokern.

## Abstract

In pharmacoepidemiologic studies of COVID-19, there were concerns about bias from residual confounding. We applied high-dimensional propensity scores (HDPS) to a case study investigating the role of inhaled corticosteroids (ICS) in COVID-19 to adjust for unmeasured confounding.

We selected patients with chronic obstructive pulmonary disease on 01 March 2020 from Clinical Practice Research Datalink (CPRD) Aurum, comparing ICS/LABA/(+-LAMA) and LABA/LAMA users. ICS effects on the outcomes COVID-19 hospitalisation and death were assessed through weighted and unweighted Cox proportional hazards models. HDPS were estimated from primary care clinical records, prescriptions and hospitalisations. SNOMED-CT codes and dictionary of medicines and devices codes from CPRD Aurum were mapped to International Classification of Disease 10^th^ revision codes and British National Formulary paragraphs respectively. We estimated propensity scores (PS) combining prespecified and HDPS covariates, selecting the top 100, 250, 500, 750 and 1000 covariates ranked by confounding potential.

When excluding triple therapy users, the conventional PS-weighted estimates showed weak evidence of increased risk of COVID-19 hospitalisation among ICS users (HR 1.19 (95% CI 0.92-1.54)). Results varied slightly based on the number of covariates included in HDPS (HR using 100 HDPS covariates 1.01 (95% CI 0.76-1.33), HR using 250 HDPS covariates 1.24 (95% CI 0.83-1.87)).

For COVID-19 death, conventional PS-weighted models showed weak evidence of harm of ICS when excluding triple therapy users (HR 1.24 (95% CI 0.87-1.75)). HDPS-weighting moved estimates toward the null, suggesting no effect of ICS (HR using 250 HDPS covariates excluding triple therapy 1.08 (95% CI 0.73- 1.59)).

HDPS may have provided better confounding control for COVID-19 deaths and may be able to partially compensate for suboptimal comparison groups. HDPS results can be sensitive to the number of covariates included, highlighting the importance of sensitivity analyses.

**Key points:**

- Residual confounding, including residual confounding by indication, is a major concern in pharmacoepidemiologic studies of COVID-19 outcomes.
- We apply high-dimensional propensity scores (HDPS) to adjust for residual confounding in a case study of inhaled corticosteroids (ICS) on COVID-19 hospitalisation and death in CPRD Aurum.
- Conventional PS-weighted analyses suggested harmful effects of ICS on COVID-19 hospitalisation and, to a lesser extent, deaths.
- HDPS weighted analyses of COVID-19 hospitalisations were sensitive to the number of covariates included, with results moving towards the null for smaller number of covariates and away from the null when including more covariates, while for deaths, estimates moved towards the null consistently.
- HDPS demonstrated promise in addressing confounding even when comparison groups are suboptimal, but its performance depends on the careful selection and ranking of covariates.

**Plain Language Summary:** A key challenge when researching the effects of medications using electronic health records is accounting for the fact that people who receive different medications often differ in important ways. Such differences, called confounding, is typically accounted for using statistical methods which require researchers to pre-specify all important confounders. A newer method, called high-dimensional propensity scores (HDPS), uses a data-driven approach to select what confounders to account for instead. These methods have not yet been applied to studies of inhaled corticosteroids and COVID-19 outcomes, an area where studies have found conflicting findings. We used electronic health records from the UK to compare the risk of COVID-19 hospitalisation and death among patients with chronic obstructive pulmonary disease taking two different treatments (ICS/LABA and LABA/LAMA) using both conventional and HDPS methods. Our findings showed that HDPS can reduce important differences between patients (confounding), but that the results can be sensitive to the number of covariates included. This demonstrates the value of HDPS and the need for researchers to run their analysis using several different assumptions.

## 1. Introduction

In non-interventional research, it can be difficult to control for hard-to-measure concepts such as frailty or disease severity. Many methods exist to mitigate residual confounding, for example using quantitative bias analysis (QBA) and e-values. Most QBA methods rely on pre-specifying the prevalence of and exposure/outcome association with a single confounder. High-dimensional propensity scores (HDPS) are an alternative method aiming to optimise confounding adjustment by considering recorded data as proxy variables.^1^

Inhaled corticosteroids (ICS) are commonly used anti-inflammatory therapies for COPD and were explored as a potential treatment for COVID-19. Randomized controlled trials (RCTs) indicated that one inhaled ICS, budesonide, had a protective effect against severe COVID-19 in patients with mild COVID-19^2,3^, but found no effect of ciclesonide or fluticasone.^4–6^ Observational studies also examined the relationship between ICS and COVID-19 outcomes, but findings were inconsistent and potentially influenced by biases including residual confounding.^7,8^

We aimed to control for residual confounding using HDPS when investigating the association between ICS and COVID-19 hospitalisation and death in CPRD Aurum.

## 2. Methods

The study protocol was registered on ENCEPP EU PAS (register number 47885). This analysis was planned post-hoc after results^9^ suggesting residual confounding may explain observed harmful associations.

### Data

The data sources, study population, and exposure, outcome and covariate definitions have previously been described^9^, and are summarised below.

#### Data source

This study used UK primary care data recorded in the Clinical Practice Research Datalink (CPRD) Aurum. CPRD Aurum includes data on 41 million patients (May 2022 build), with >13 million patients currently registered (20% of UK population)^10^ from >1,300 practices using EMIS management software.^11^ CPRD Aurum is broadly representative of the English population.^11^

CPRD Aurum was linked to Hospital Episode Statistics (HES) Admitted Patient Care (APC) and Office for National Statistics (ONS) Death Registry.^11,12^ HES APC holds information on all in-patient contacts at NHS hospitals in England.^12,13^ The ONS death registry contains information on deaths occurring in England and Wales, including cause of death documented using International Classification of Disease 10^th^ revision (ICD-10) codes.^13,14^ Data was also linked to the Index of Multiple Deprivation (IMD), a postcode-level indicator of socioeconomic status.

#### Study population

We defined a cohort diagnosed with COPD before 01^st^ March 2020 (index date) based on a validated algorithm.^15^ Patients had to be alive aged ≥35, and registered in CPRD Aurum on the index date. Patients needed to have 12 months’ continuous registration and a record of current/former smoking before the index date. We excluded people with asthma recorded within three years before the index date, leukotriene receptor antagonist use within 4 months before the index date as this indicates asthma, or other chronic respiratory disease at any point before the index date. Patients were followed until death (recorded in ONS or CPRD), deregistration, or 31^st^ August 2020, whichever came first. If death was registered in ONS, we used that date as the date of death. If death was missing in ONS but registered in CPRD, we used the date recorded in CPRD as the date of death.

##### Exposure

Continuous treatment episodes were generated based on the recorded prescription issue date and information on intended duration, prescribed amount, and dosage.^9^

We used treatment episodes to categorise people as exposed to ICS/LABA or LABA/LAMA on the index date. We conducted two separate analyses. In the first, people using ICS+LABA+LAMA (i.e., triple therapy) were included in the ICS/LABA group. In the second, triple therapy users were excluded as we expected these patients to be sicker than those on dual therapy. ICS/LABA was the exposure of interest and LABA/LAMA was the active comparator. We did not censor at the estimated discontinuation dates due to low discontinuation rates and concern that discontinuations would be inaccurately estimated due to pandemic-related healthcare disruptions.^16^

##### Outcome

Outcomes were i) hospitalisation with a primary diagnosis code for COVID-19 recorded in HES APC, and ii) death with a diagnosis code for COVID-19 as cause of death anywhere on the death certificate in the ONS Death Registry. COVID-19 hospitalisation and death were identified with ICD-10 codes U07.1 and U07.2.

#### Prespecified Covariates

The following covariates, selected based on input from a practising clinician, were automatically included in propensity scores (PSs): age, gender, BMI (most recent within 10 years, categorised as underweight (<18.5), normal (18.5-24.9), overweight (25-29.9), or obese (≥30)), smoking (current vs former), ethnicity, cancer (ever), diabetes (ever), chronic kidney disease (ever), cardiovascular disease (ever), hypertension (ever), asthma (>3 years ago), immunosuppression, influenza vaccination (past year), pneumococcal vaccination (past 5 years), IMD quintile, and number of COPD exacerbations in the past 12 months, based on a validated algorithm.^17^

### Statistical analyses

Cohort characteristics were summarised using descriptive statistics by exposure group. There were missing data for BMI (n = 550), ethnicity (n = 7785) and IMD (n = 42). Missing BMI was assumed to be normal.^18^ Missing ethnicity was treated as a separate category^19^, and patients with missing IMD were excluded as this indicates poor quality records.

We estimated PSs and used inverse probability of treatment weighting (IPTW) to estimate the average treatment effect (ATE) adjusting for potential confounders. Conventional PSs were estimated using logistic regression including the covariates listed above. Weights were calculated as 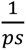 (ICS/LABA) and 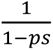 (LABA/LAMA), where the PS is the probability of receiving ICS. Overlap of the PSs across treatment groups was assessed graphically and by summarising PSs by treatment group. PSs were trimmed to the region of common support.^20^ This was applied for all analyses.

Cox regression was used to estimate hazard ratios (HRs) and 95% confidence intervals (CIs) for the association between exposure and outcomes. The proportional hazards assumption was assessed using Schoenfeld residuals.

### High-dimensional propensity scores

When applying HDPS, data is separated into dimensions reflecting different care facets.^1^ We separated data into clinical (CPRD Aurum), prescription (CPRD Aurum), and hospitalisation (HES APC) information.^21^ In CPRD Aurum, clinical observations are coded using SNOMED-CT, while prescriptions are coded using dictionary of medicines and devices (dm+d) codes. Observations in HES are recorded using ICD-10 codes. ICD-10 codes are hierarchical, with the first three characters denoting clinical categories. To avoid sparsity, SNOMED-CT codes were mapped to ICD-10 using a mapping file supplied by the NHS, and subsequently truncated to three characters.^22,23^ dm+d codes were mapped to British National Formulary (BNF) paragraphs using a mapping file from the Bennett Institute.^24^ For unmatched dm+d codes, the first 6 characters of the BNF chapter variable supplied in the CPRD code browser were used. Unmapped codes that had over 1,000,000 recordings in CPRD Aurum were mapped manually (n = 16). The top 100 unmapped codes for clinical observations and prescriptions are in Supplementary Table 1 and 2. BNF paragraphs correspond to drug classes or treatment indications rather than specific mechanisms of action (e.g., paragraph 030101 corresponds to bronchodilators, specifically adrenoceptor agonists).^21^ We excluded medications used to define the treatment groups (ICS, LABA and LAMA, as single-ingredient or combined inhalers), and ICD-10 codes used to define outcomes (U07) from the HDPS.

We assessed occurrence of codes in the 12 months before the index date. The original algorithm applies a prevalence filter which includes the top codes ranked based on marginal prevalence.^1^ In line with Schuster et al., we did not apply this filter as variables with a low marginal prevalence can still be important confounders.^25^

For each patient, we generated variables indicating if a code was recorded once (≥1), sporadically (≥median) or frequently (≥75%-ile) in the year before the index date, relative to the number of occurrences of the relevant code per patient among those who had ≥1 occurrence of the code. For the clinical dimension, we considered codes that were ever-present, reflecting the fact that conditions, particularly chronic ones, may not be recorded at every consultation. Consistently with other applications of HDPS in CPRD, this information was included by redefining “once” variables for this dimension.^21^

Covariates were prioritised using the Bross formula, which using covariate prevalence in exposed (P_C1_) and unexposed (P_C0_), and the relative risk between confounder and outcome (RR_CD_) to rank covariates by their potential to bias the exposure-outcome association.^26^

We excluded variables that behaved like instrumental variables, i.e. strongly associated with exposure but not outcomes; defined using cut-offs based on the relative risk between confounder and exposure (RR_CE_) and the RR_CD_ (|log(RR_CE_)|>1.1 and |log(RR_CD_)|<0.5).^23^

Covariates identified using HDPS were included in logistic regression models alongside predefined covariates to estimate PSs. Subsequently, IPTWs were used to weight logistic regression and Cox models to estimate the impact of ICS on the outcomes. We calculated standardised mean differences (SMDs) in covariate distributions between treatment groups before PS-weighting, after conventional PS-weighting using, and after HDPS-weighting.

We varied the number of covariates included in the HDPS (100, 250, 500, 750, 1000), with no predefined primary number specified. The top 30 ranked covariates for each cohort and outcome are available in Supplementary Tables 3-6. Post-hoc, we conducted an analysis where covariates ranked 1^st^-250^th^ were added one by one to assess the sensitivity of results to the inclusion of particular covariates.

Data was managed using Stata version 17.0^27^ and analysis carried out using R (version 4.3.3)^28^. Code lists and data management and analysis code is on GitHub (https://github.com/bokern/ics_hdps).

## 3. Results

Cohort characteristics have previously been described.^9^ We included 56,029 patients using ICS/LABA (14,905 excluding triple therapy users) and 22,307 patients using LABA/LAMA at baseline (Table 1).

**Table 1.** baseline cohort characteristics.

|  | Including triple therapy users |  |  |  |  |  | Excluding triple therapy users |  |  |  |  |  |
| --- | --- | --- | --- | --- | --- | --- | --- | --- | --- | --- | --- | --- |
|  |  |  | Standardised mean differences |  |  |  |  |  | Standardised mean differences |  |  |  |
|  | LABA/LAMA<br>(n = 22307) | ICS/LABA<br>(n = 56029) | unweighted | weighted<br>conventional<br>PS | 250 HDPS<br>covariates<br>(Hospitalisation) | 250 HDPS<br>covariates<br>(Death) | LABA/LAMA<br>(n = 22307) | ICS/LABA<br>(n = 14905) | unweighted | weighted<br>conventional<br>PS | 250 HDPS<br>covariates<br>(Hospitalisation) | 250 HDPS<br>covariates<br>(Death) |
| Gender = Female | 10068 (45.1) | 26217 (46.8) | 0.033 | 0.004 | 0.002 | 0.001 | 10068 (45.1) | 7089 (47.6) | 0.049 | 0.001 | <0.001 | 0.001 |
| Age (mean (SD)) | 70.83 (10.23) | 71.32 (10.47) | 0.048 | 0.002 | 0.01 | 0.007 | 70.83 (10.23) | 71.38 (11.29) | 0.051 | 0.002 | 0.002 | 0.002 |
| IMD |  |  | 0.045 | 0.004 | 0.005 | 0.007 |  |  | 0.027 | 0.002 | 0.002 | 0.002 |
| 1 | 3048 (13.7) | 7178 (12.8) |  |  |  |  | 3048 (13.7) | 2021 (13.6) |  |  |  |  |
| 2 | 3813 (17.1) | 9213 (16.4) |  |  |  |  | 3813 (17.1) | 2562 (17.2) |  |  |  |  |
| 3 | 4077 (18.3) | 9971 (17.8) |  |  |  |  | 4077 (18.3) | 2759 (18.5) |  |  |  |  |
| 4 | 5012 (22.5) | 12722 (22.7) |  |  |  |  | 5012 (22.5) | 3473 (23.3) |  |  |  |  |
| 5 | 6357 (28.5) | 16945 (30.2) |  |  |  |  | 6357 (28.5) | 4090 (27.4) |  |  |  |  |
| Diabetes | 5515 (24.7) | 14074 (25.1) | 0.009 | 0.003 | 0.01 | 0.011 | 5515 (24.7) | 3684 (24.7) | <0.001 | 0.001 | <0.001 | 0.001 |
| Hypertension | 11312 (50.7) | 28592 (51.0) | 0.006 | 0.003 | 0.002 | 0.006 | 11312 (50.7) | 7689 (51.6) | 0.018 | 0.001 | <0.001 | <0.001 |
| Cardiovascular disease | 6538 (29.3) | 16766 (29.9) | 0.013 | 0.002 | 0.003 | 0.005 | 6538 (29.3) | 4387 (29.4) | 0.003 | <0.001 | <0.001 | <0.001 |
| Cancer | 4413 (19.8) | 10539 (18.8) | 0.025 | 0.001 | 0.005 | 0.008 | 4413 (19.8) | 2827 (19.0) | 0.021 | 0.001 | 0.001 | 0.001 |
| Past asthma | 2664 (11.9) | 15339 (27.4) | 0.396 | 0.002 | 0.022 | 0.011 | 2664 (11.9) | 4134 (27.7) | 0.404 | 0.001 | 0.002 | 0.002 |
| Kidney impairment | 6737 (30.2) | 16719 (29.8) | 0.008 | 0.007 | 0.012 | 0.012 | 6737 (30.2) | 4567 (30.6) | 0.01 | 0.001 | 0.002 | 0.001 |
| Immunosuppression | 274 (1.2) | 664 (1.2) | 0.004 | <0.001 | 0.003 | 0.004 | 274 (1.2) | 188 (1.3) | 0.003 | <0.001 | <0.001 | <0.001 |
| Influenza vaccine | 17955 (80.5) | 44884 (80.1) | 0.01 | 0.001 | <0.001 | 0.002 | 17955 (80.5) | 11389 (76.4) | 0.099 | <0.001 | <0.001 | <0.001 |
| Pneumococcal vaccine | 3236 (14.5) | 5983 (10.7) | 0.116 | 0.001 | 0.001 | <0.001 | 3236 (14.5) | 1497 (10.0) | 0.136 | 0.001 | <0.001 | 0.001 |
| COPD exacerbation in past 12 months | 6221 (27.9) | 22793 (40.7) | 0.272 | 0.002 | 0.005 | 0.001 | 6221 (27.9) | 4638 (31.1) | 0.071 | 0.001 | <0.001 | 0.001 |
| Former smoking | 12239 (54.9) | 33280 (59.4) | 0.092 | 0.002 | 0.01 | 0.008 | 12239 (54.9) | 8942 (60.0) | 0.104 | 0.001 | 0.001 | 0.001 |
| Ethnicity (%) |  |  | 0.051 | 0.009 | 0.015 | 0.009 |  |  | 0.101 | 0.002 | 0.004 | 0.004 |
| White | 19574 (87.7) | 49366 (88.1) |  |  |  |  | 19574 (87.7) | 12890 (86.5) |  |  |  |  |
| South Asian | 197 (0.9) | 742 (1.3) |  |  |  |  | 197 (0.9) | 292 (2.0) |  |  |  |  |
| Black | 130 (0.6) | 351 (0.6) |  |  |  |  | 130 (0.6) | 138 (0.9) |  |  |  |  |
| Mixed | 49 (0.2) | 142 (0.3) |  |  |  |  | 49 (0.2) | 44 (0.3) |  |  |  |  |
| Unknown | 2357 (10.6) | 5428 (9.7) |  |  |  |  | 2357 (10.6) | 1541 (10.3) |  |  |  |  |
| BMI (%) |  |  | 0.07 | 0.004 | 0.009 | 0.009 |  |  | 0.038 | 0.001 | 0.002 | 0.002 |
| Underweight (<18.5) | 969 (4.3) | 3138 (5.6) |  |  |  |  | 969 (4.3) | 619 (4.2) |  |  |  |  |
| Normal (18.5-24.9) | 6922 (31.0) | 18153 (32.4) |  |  |  |  | 6922 (31.0) | 4629 (31.1) |  |  |  |  |
| Overweight (25-29.9) | 7167 (32.1) | 17339 (30.9) |  |  |  |  | 7167 (32.1) | 5020 (33.7) |  |  |  |  |
| Obese (>=30) | 7249 (32.5) | 17399 (31.1) |  |  |  |  | 7249 (32.5) | 4637 (31.1) |  |  |  |  |

### Propensity Score Diagnostics

Figure 1 shows unweighted and weighted PS distributions. For all analyses, trimming PSs to the region of common support did not exclude many patients (<1%). Prespecified covariates were balanced after weighting (SMD <0.1). Covariate balance was better for the predefined covariates when estimating the PS using only those covariates. However, weighting using HDPS achieved adequate balance (SMD <0.1) of predefined covariates while achieving better balance for the additional HDPS covariates (Figure 2, Table 1). For illustration, we present diagnostics for 250 covariates for COVID-19 hospitalisations below (Figure 3, Figure 2, Table 1); diagnostics for other implementations of the HDPS showed similar results and are in supplementary material.

**Figure 1.**
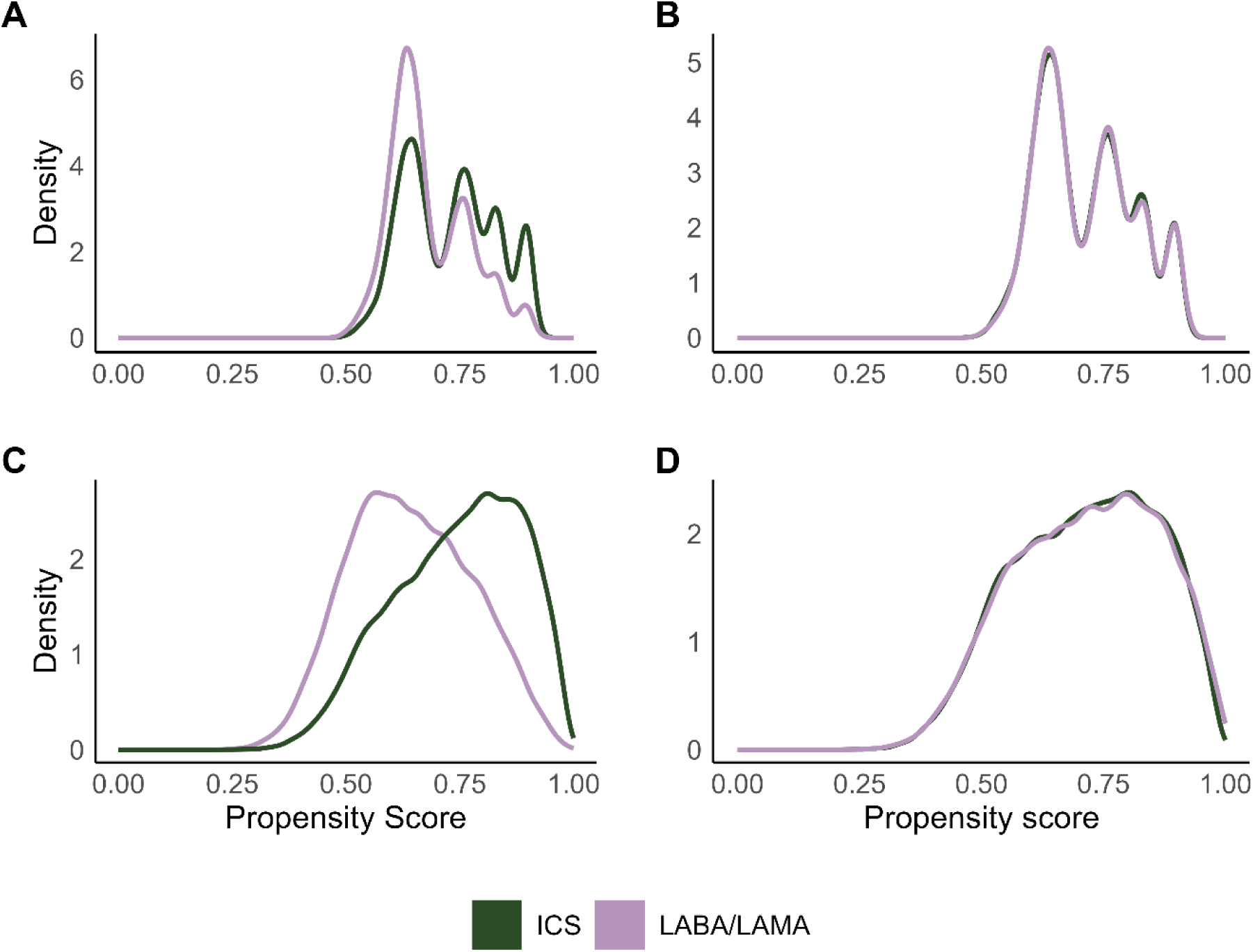
Density distributions of propensity scores for ICS and LABA/LAMA treatment groups before and after weighting. A) unweighted propensity score distributions using predefined covariates, B) weighted propensity score distribution, C) high-dimensional propensity score (HDPS) distribution for 250 covariates, for the outcome COVID-19 hospitalisation, D) weighted HDPS distribution for 250 covariates, for the outcome COVID-19 hospitalisation

**Figure 2.**
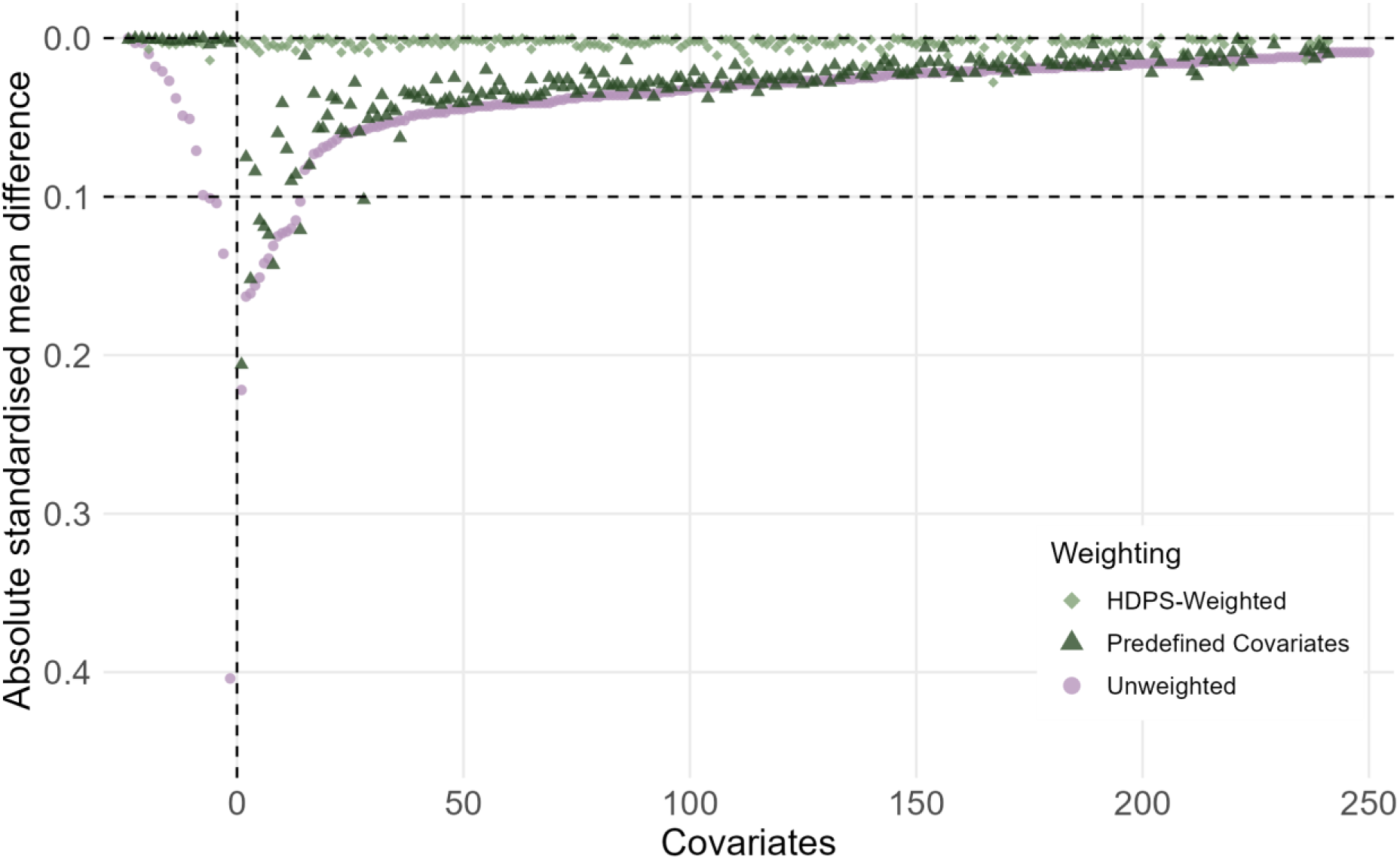
Comparison of absolute standardised differences in the predefined and top 250 high-dimensional propensity score covariates between unweighted, predefined and HDPS weighted samples for COVID-19 hospitalisations, excluding triple therapy users

**Figure 3.**
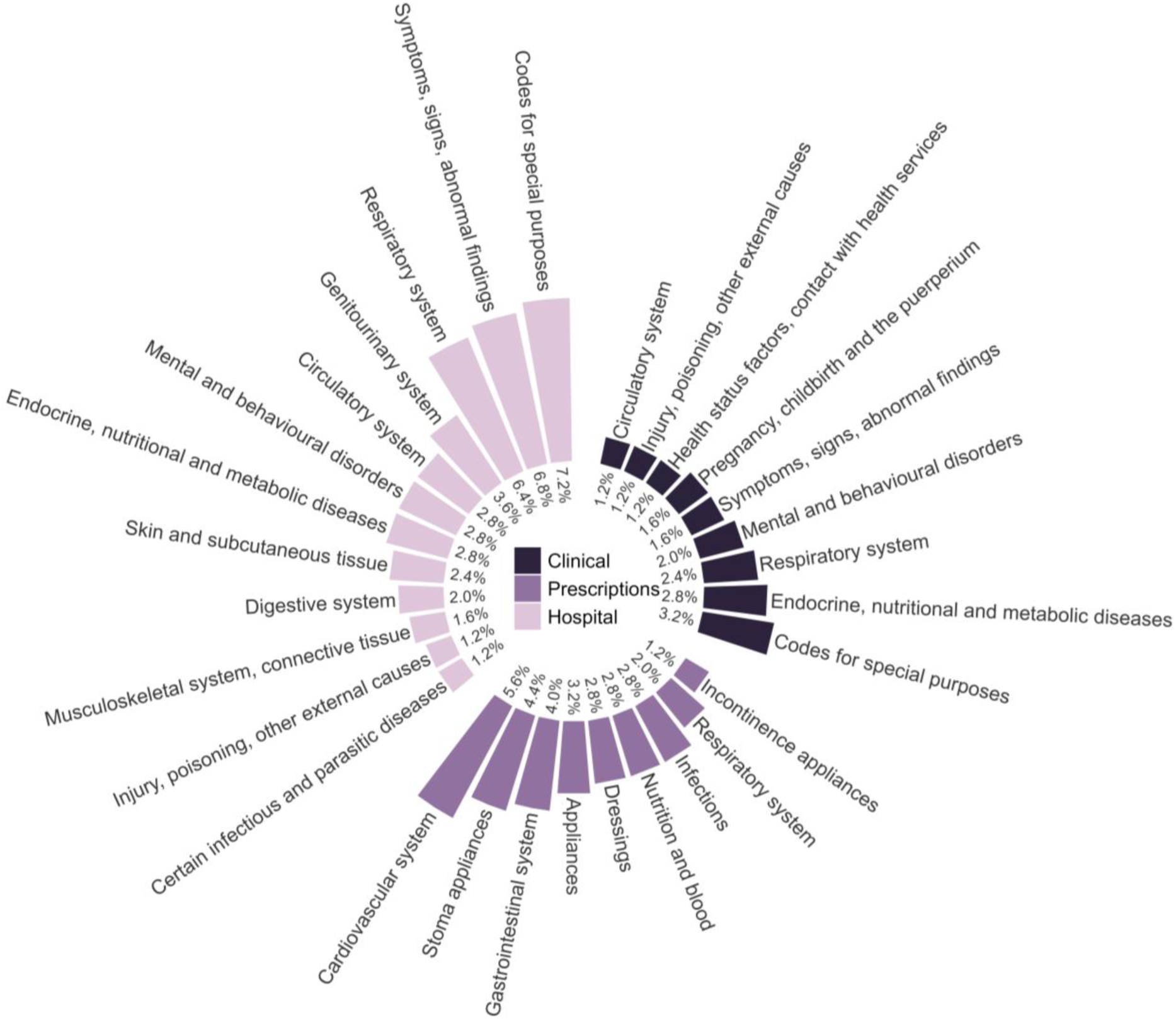
Summary of high-level concepts captured in the top 250 Bross-prioritised high-dimensional propensity score pre-exposure covariates colour-coded by data dimension for COVID-19 hospitalisations, excluding triple therapy users

For COVID-19 hospitalisations including triple therapy users, the top 250 codes included 145 (58.0%) from hospital data, 82 prescription (32.8%), and 23 primary care observations (9.2%) (Figure 3).

### Effects of ICS on COVID-19 hospitalisations

Conventional PS-weighted analyses including triple therapy users suggested an increased risk of COVID- 19 hospitalisation among ICS users (HR 1.46 (95% CI 1.19-1.79)), which was attenuated when excluding triple therapy users (HR 1.19 (95% CI 0.92-1.54)). All HDPS applications moved effect estimates towards null (Figure 4). Point estimates varied slightly depending on the number of included covariates (HR using 100 HDPS-covariates 1.19 (95% CI 0.93 – 1.52), HR using 250 HDPS-covariates 1.22 (0.96 – 1.56)). When excluding triple therapy users, results were more sensitive to the number of covariates included in the HDPS. The HDPS calculated using the top 100 covariates gave an almost exact null result (HR 1.01 (95% CI 0.76 – 1.33)). Including more covariates shifted the effect estimate away from null (HR using 250 HDPS- covariates 1.24 (95% CI 0.83-1.87)) although CIs were wide and remained consistent with null effects.

**Figure 4.**
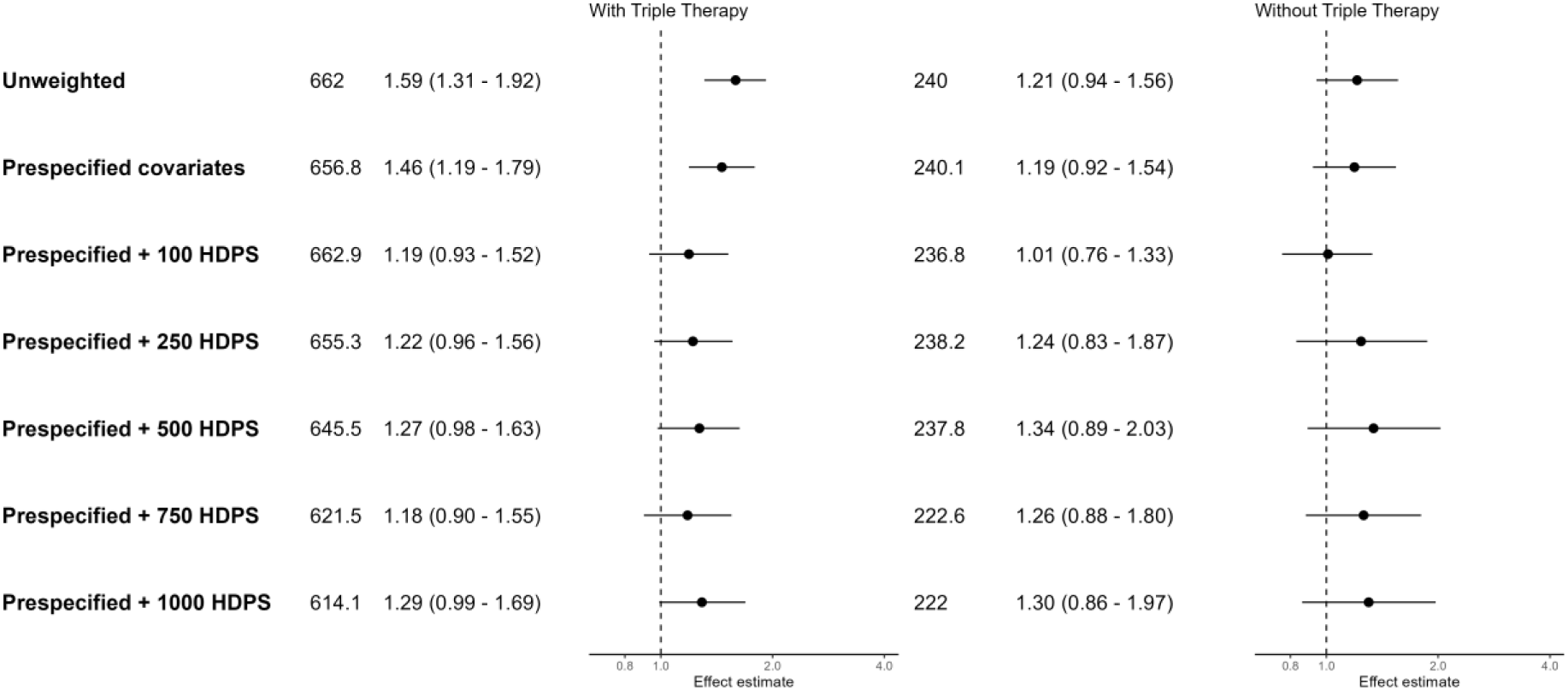
Forest plot of hazard ratios and 95% confidence intervals for COVID-19 hospitalisations, comparing ICS/LABA (+/- LAMA) users to LABA/LAMA users. Effect estimates >1 indicate an increased risk in the ICS group.

### Effects of ICS on COVID-19 death

Conventional PS-weighted models showed evidence of harm of ICS when including triple therapy users (HR 1.42 (95% CI 1.08-1.87)) and very weak evidence of a harm (HR 1.24 (95% CI 0.87-1.75)) when excluding triple therapy users. Using HDPS shifted estimates towards the null (HR using 100 HDPS covariates including triple therapy users 1.15 (95 % CI 0.82-1.60), HR using 100 HDPS covariates excluding triple therapy users 1.05 (95% CI 0.72-1.53)) (Figure 5). For both cohorts, the CIs for all applications of HDPS crossed the null and were insensitive to the number of covariates. With more covariates, the estimates for the cohort including triple therapy users became very close to those in the cohort excluding triple therapy users (HR using 1000 HDPS covariates including triple therapy 1.15 (95% CI 0.78-1.69), excluding triple therapy 1.12 (95% CI 0.75-1.66)).

**Figure 5.**
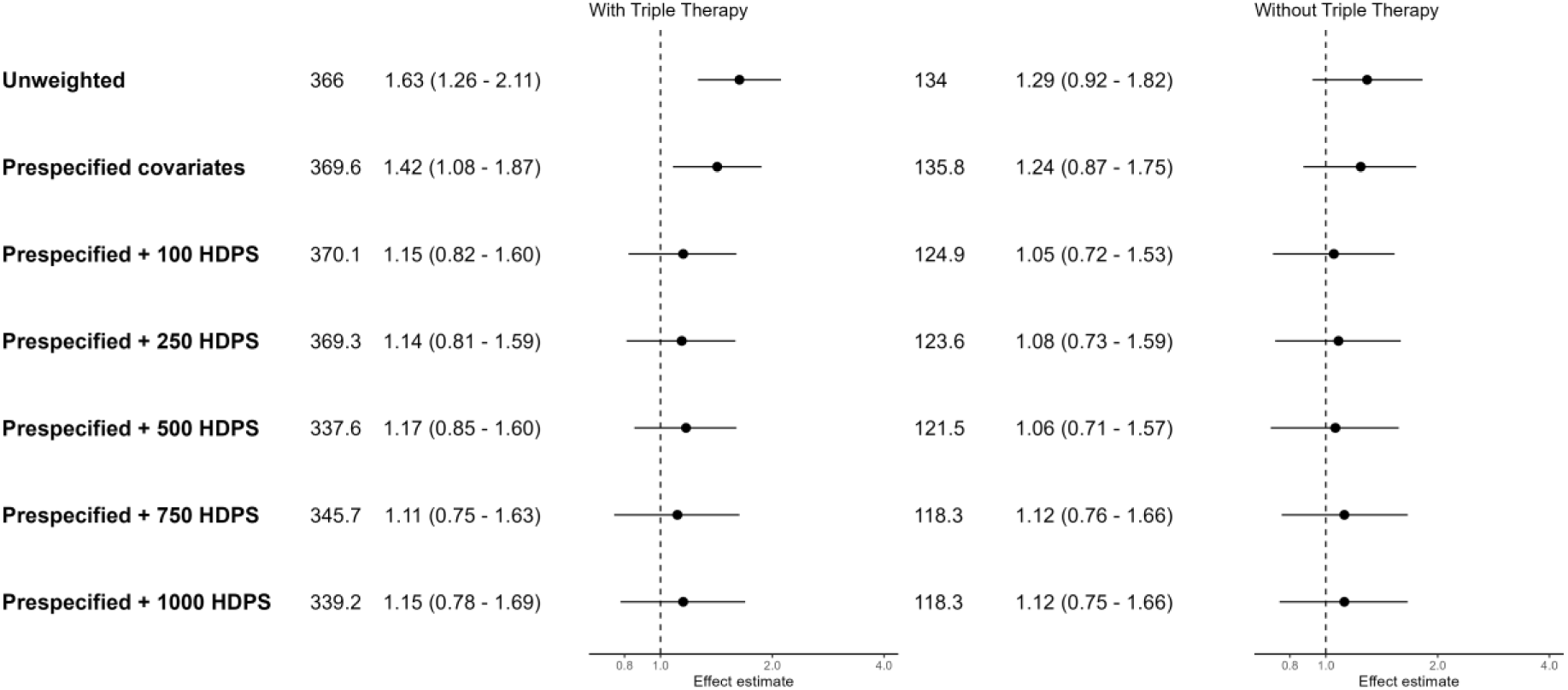
Forest plot of hazard ratios and 95% confidence intervals for COVID-19 deaths, comparing ICS/LABA (+/- LAMA) users to LABA/LAMA users. Effect estimates >1 indicate an increased risk in the ICS group.

### Sensitivity analyses

Adding covariates one by one showed that certain covariates made substantial changes in the effect estimates (Figure 6). The largest changes occurred when individuals with rare combinations of covariates received a large weight and experienced the outcome. These combinations of covariates cannot be disclosed for data privacy reasons.

**Figure 6.**
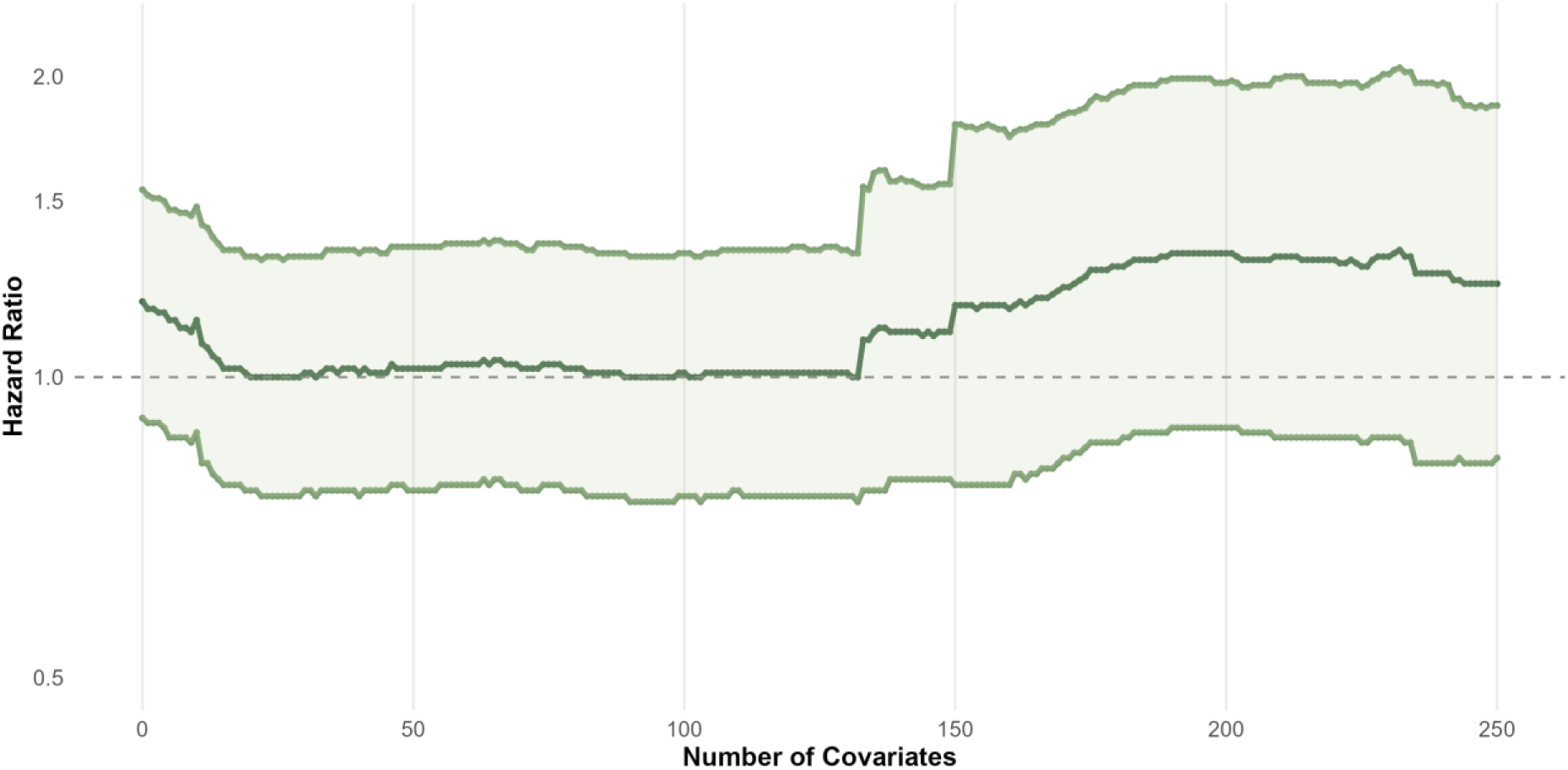
Sensitivity analyses assessing the impact of the number of high-dimensional propensity score covariates selected on the effect estimate, adding the top 250 Bross-ranked covariates one by one. Propensity scores were estimated using logistic regression and treatment effects were estimated using an inverse probability of treatment weighted Cox regression model. Outcome was COVID-19 hospitalisation, excluding triple therapy users.

For both outcomes, point estimates were identical when using Cox and logistic regression models. However, CIs were consistently narrower when using logistic regression models. Results of the logistic regression models are in Supplementary Figures 41 and 42.

## 4. Discussion

Using conventional IPT-weighted models, we observed an increased risk of COVID-19 hospitalisation and death among ICS users when including triple therapy users, but no evidence of increased risk when excluding triple therapy users. For COVID-19 hospitalisations, results were sensitive to the number of covariates included in HDPS, with the point estimate moving closer to the null or further away depending on the comparison group used. For COVID-19 deaths, HDPS consistently moved the point estimate closer to the null, with no evidence against the null hypothesis. The analyses including and excluding triple therapy users gave very similar results after HDPS weighting. This is the first study to implement HDPS in CPRD Aurum, building upon previous work to implement HDPS in CPRD GOLD.^21^

### Comparison to previous studies

Many studies have applied HDPS in CPRD GOLD, with most using 500 covariates to estimate HDPS^29–39^, although some used as few as 40^40,41^. Most present results applying HDPS, although one presented results comparing conventional PS-weighting to HDPS-weighting, finding superior confounding control with HDPS.^32^ Another study compared HDPS-decile adjusted results to results adjusted for prespecified covariates only and found similar effect estimates in the two analyses.^38^

No pharmacoepidemiologic study applied HDPS to UK electronic health record (EHR) data during the COVID-19 pandemic. However, several studies investigated the same clinical question. A cohort study using the EHR platform OpenSAFELY^7^ found an aHR for COVID-19 death 1.39 (1.10–1.76) among patients with COPD. Using E-values, the authors found that an unmeasured confounder would need to be associated with either exposure or outcome by a risk ratio of at least 1.43 to explain the association.

A QResearch study ^8^ found a more modest risk of severe COVID-19 and COVID-19 death associated with ICS use (COVID-19 hospitalisation aHR 1.13 (1.03-1.23), COVID-19 death aHR 1.15 (1.01-1.31)). Compared with our study, this study included more prespecified confounders. Our results after HDPS are very similar to the fully-adjusted results from this study.

### Interpretation

For both cohorts and outcomes, the highest-ranked empirically identified covariates included codes relating to nicotine dependence and lung disease (Supplementary tables 3-6). This suggests that variables related to respiratory disease were potentially important confounders, and HDPS may have provided better adjustment for this than conventional PS-weighting. Compared to other approaches such as e-values or QBA, HDPS offers the advantage of not having to specify a single unmeasured confounder and its strength of association with the exposure and outcome, and can adjust for multiple hard-to-measure concepts including confounding by indication.

For hospitalisations, results were sensitive to the number of HDPS covariates included, illustrating the importance of varying the number of covariates in sensitivity analyses. A sensitivity analysis that included covariates one by one showed that even low-ranked covariates can result in substantial changes in the effect estimates if patients with unusual combinations of covariates have the outcome, especially if the outcome is rare. It is possible that hospitalisation is a paradoxical outcome, with a selection process occurring regarding which patients are admitted to hospital with COVID-19. We assumed patients in the ICS group were sicker and would have more severe COVID-19 compared to those using LABA/LAMA, and therefore better control for confounding by disease severity would move effect estimates towards the null. However, it is possible that for COVID-19 hospitalisations, confounding operated in both directions, with sicker or frail patients less likely to be hospitalised if they were deemed unlikely to survive.^42^ The extent of this selection process may have varied over time depending on changes hospital pressures or triaging processes. Similar issues were identified when using intensive care unit admission as an outcome.^42^ ONS data on place of death indicates that during our study period around 28% of COVID-19 deaths occurred outside hospitals^43^, suggesting that not everyone with severe COVID-19 was admitted to hospital and there may have been selection into hospitals. Therefore, COVID-19 hospitalisation may not have been a suitable outcome to represent severe COVID-19 if those with the most severe COVID-19 died without hospitalisation.

For deaths, point estimates for the two treatment cohorts were similar. Inclusion of the 100 highest-ranked covariates in the HDPS shifted the effect estimate towards null, while inclusion of additional covariates did not substantially change effect estimates. With inclusion of more covariates, effect estimates in the two cohorts became more similar. The cohort including triple therapy had more precision due to larger sample size. In the UK, LABA/LAMA and ICS/LABA are commonly used treatment combinations for COPD. Treatment should be stepped up to triple therapy if COPD remains uncontrolled. We therefore expect patients using triple therapy to have more severe COPD and be sicker than patients on dual therapy. As a result, excluding triple therapy from the ICS/LABA group should provide a better comparison to the LABA/LAMA group. Taken together, this indicates that HDPS may offer better confounding control than conventional PS-weighting and may be able to compensate for suboptimal comparator groups in some cases.

### Strengths and Limitations

Strengths of this study include the size and representativeness of CPRD Aurum and comprehensive capture of hospitalisations and deaths.

Due to short follow-up, we could not use a new-user design, and assumed any effect of ICS on the outcomes was independent of treatment history and length. Stockpiling of medications in the early months of the pandemic may have led to inaccurate estimations of exposure durations^16^, potentially causing exposure misclassification. Additionally, we did not have information on in-hospital prescriptions. We observed minor deviations from proportional hazards, but graphical inspection indicated this was likely due to low numbers of outcomes and the characteristics of people with severe COVID-19 changing throughout the study period. The HRs presented should be interpreted as an average over the time period.

The Bross formula used to prioristise variables for HDPS is suited to binary covariates only. Continuous confounders such as laboratory or diagnostic test results do not naturally get considered in the HDPS. Therefore, if these factors are important confounders, they will not directly be accounted for. In this cohort, codes relating to forced expiratory volume in one second measurements or Medical Research Council Dyspnoea Scale may have additionally controlled for confounding by COPD severity.^44^

### Conclusions

The results provide weak evidence of a harmful association between ICS and COVID-19 hospitalisation, but no effect on COVID-19 death. The analysis of deaths suggests that HDPS can achieve better confounding control than conventional PS weighting and compensate for suboptimal comparator groups in some instances. In cases where the confounding structure is unclear, such as for the outcome COVID-19 hospitalisations, using a data-driven approach such as HDPS can lead to inconsistent results that may be sensitive to the inclusion of even low-ranked covariates, highlighting the importance of sensitivity analyses.

## Data Availability

No additional data available. Data management and analysis code and all code lists are available on our GitHub repositories. (https://github.com/bokern/ics_hdps and https://github.com/bokern/ics_covid)

https://github.com/bokern/ics_covid

## Acknowledgments

This study is based in part on data from the Clinical Practice Research Datalink obtained under licence from the UK Medicines and Healthcare products Regulatory Agency. The data is provided by patients and collected by the NHS as part of their care and support. The interpretation and conclusions contained in this study are those of the author/s alone. The study was approved by the Independent Scientific Advisory Committee (approval number: 22_001876).

## Ethics Approval

The study was approved by the London School of Hygiene and Tropical Medicine Research Ethics Committee (Reference: 27896).

## Author contributions

MB, JT, AS, CTR, IJD contributed to the study design. MB conducted the data management and analysis and drafted the manuscript.

All authors contributed to reviewing and editing of the manuscript. All authors were involved in design and conceptual development and reviewed and approved the final manuscript.

