## Supplement for "Adjusting for residual confounding using high-dimensional propensity scores in a study of inhaled corticosteroids and COVID-19 outcomes"

1. Supplementary tables

Supplementary Table 1 100 most frequent unmatched SNOMED-CT codes.

| Rank | SnomedCTConceptId | N | Term |
| --- | --- | --- | --- |
| 1 | 1572871000006100 | 2646163 | Awaiting clinical code migration to EMIS Web |
| 2 | 279991000000102 | 2551983 | SMS text message sent to patient |
| 3 | 279991000000102 | 2551983 | SMS (short message service) text message sent to patient |
| 4 | 279991000000102 | 2551983 | Short message service text message sent to patient |
| 5 | 498521000006103 | 1525015 | Attachment |
| 6 | 72313002 | 668205 | Systolic arterial pressure |
| 7 | 72313002 | 668205 | Systolic blood pressure |
| 8 | 1091811000000100 | 667919 | Diastolic arterial pressure |
| 9 | 163020007 | 660161 | O/E - blood pressure reading |
| 10 | 163020007 | 660161 | O/E - blood pressure |
| 11 | 163020007 | 660161 | O/E - BP reading |
| 12 | 163020007 | 660161 | O/E-blood pressure reading NOS |
| 13 | 163020007 | 660161 | On examination - blood pressure reading |
| 14 | 78564009 | 433362 | Pulse rate |
| 15 | 78564009 | 433362 | PR - Pulse rate |
| 16 | 1000731000000100 | 415607 | Serum creatinine NOS |
| 17 | 1000731000000100 | 415607 | Serum creatinine level |
| 18 | 1000661000000100 | 388221 | Serum sodium level |
| 19 | 1000651000000100 | 386693 | Serum potassium level |
| 20 | 27113001 | 356678 | Body weight |
| 21 | 27113001 | 356678 | O/E - weight NOS |
| 22 | 27113001 | 356678 | Weight |
| 23 | 60621009 | 345560 | Body mass index |
| 24 | 60621009 | 345560 | Weight: body mass |
| 25 | 60621009 | 345560 | BMI - Body mass index |
| 26 | 1022431000000100 | 331357 | Haemoglobin estimation NOS |
| 27 | 1022431000000100 | 331357 | Hb estimation |
| 28 | 1022431000000100 | 331357 | Haemoglobin estimation |
| 29 | 1022541000000100 | 330596 | Total white cell count NOS |
| 30 | 1022541000000100 | 330596 | Total white blood count |
| 31 | 1022541000000100 | 330596 | Total white blood count |
| 32 | 1022541000000100 | 330596 | White blood count |
| 33 | 1022541000000100 | 330596 | White cell count |
| 34 | 1022541000000100 | 330596 | Total white cell count |
| 35 | 1022651000000100 | 327523 | Platelet count |
| 36 | 1022651000000100 | 327523 | Platelet count NOS |
| 37 | 1022551000000100 | 325184 | Neutrophil count |
| 38 | 1022551000000100 | 325184 | Granulocyte count |
| 39 | 1022551000000100 | 325184 | Granulocyte count |
| 40 | 1022491000000100 | 323940 | Mean cell volume |
| 41 | 1022491000000100 | 323940 | MCV - Mean corpuscular volume |
| 42 | 1022491000000100 | 323940 | Mean cell volume |
| 43 | 1022581000000100 | 321991 | Lymphocyte count |
| 44 | 1022561000000100 | 321775 | Eosinophil count |
| 45 | 1022561000000100 | 321775 | Eosinophil count NOS |
| 46 | 1022591000000100 | 321339 | Monocyte count |
| 47 | 1022591000000100 | 321339 | Monocyte count NOS |
| 48 | 1022471000000100 | 321091 | Mean cell haemoglobin |
| 49 | 1022471000000100 | 321091 | MCH - Mean corpuscular haemoglobin |
| 50 | 1022471000000100 | 321091 | Mean cell haemoglobin |
| 51 | 1022291000000100 | 318114 | Packed cell volume |
| 52 | 1022291000000100 | 318114 | Haematocrit - PCV |
| 53 | 1022291000000100 | 318114 | Haematocrit |
| 54 | 1022291000000100 | 318114 | Packed cell volume - PCV |
| 55 | 1022291000000100 | 318114 | Haematocrit - packed cell volume |
| 56 | 1022291000000100 | 318114 | Packed cell volume |
| 57 | 1000821000000100 | 317583 | Serum albumin level |
| 58 | 1022451000000100 | 316277 | RBC count NOS |
| 59 | 1022451000000100 | 316277 | Red blood cell count |
| 60 | 1022451000000100 | 316277 | RBC (red blood cell) count |
| 61 | 1022451000000100 | 316277 | Erythrocyte count |
| 62 | 1022571000000100 | 313054 | Basophil count |
| 63 | 1000951000000100 | 309980 | Serum urea level |
| 64 | 1000621000000100 | 287786 | Serum alkaline phosphatase level |
| 65 | 25611000000107 | 271414 | Referral letter |
| 66 | 1022441000000100 | 270864 | FBC - full blood count |
| 67 | 1018251000000100 | 270030 | ALT/SGPT serum level |
| 68 | 1018251000000100 | 270030 | Serum alanine aminotransferase level |
| 69 | 14734007 | 262161 | Administrative procedure |
| 70 | 14734007 | 262161 | Administrative procedures |
| 71 | 428481002 | 249980 | Patient mobile telephone number |
| 72 | 1022481000000100 | 244162 | MCHC - Mean corpuscular haemoglobin concentration |
| 73 | 431314004 | 241071 | Peripheral oxygen saturation |
| 74 | 431314004 | 241071 | Pulse oximetry monitoring |
| 75 | 431314004 | 241071 | SpO2 - oxygen saturation at periphery |
| 76 | 431314004 | 241071 | Pulse oximetry |
| 77 | 431314004 | 241071 | SpO2 - saturation of peripheral oxygen |
| 78 | 997531000000108 | 228728 | Liver function test |
| 79 | 394703002 | 228234 | Chronic obstructive pulmonary disease annual review |
| 80 | 999791000000106 | 227487 | Haemoglobin A1c level - International Federation of Clinical Chemistry and Laboratory Medicine standardised |
| 81 | 313334002 | 222174 | Blood sample taken |
| 82 | 313334002 | 222174 | Nursing care blood sample taken |
| 83 | 1000971000000100 | 211145 | Urea and electrolytes level |
| 84 | 248333004 | 208537 | Standing height |
| 85 | 248333004 | 208537 | O/E - height NOS |
| 86 | 997591000000109 | 208053 | Serum total bilirubin level |
| 87 | 1020291000000100 | 204903 | GFR (glomerular filtration rate) calculated by abbreviated Modification of Diet in Renal Disease Study Group calculation |
| 88 | 2051000000104 | 201713 | Letter sent to patient |
| 89 | 993501000000105 | 201207 | Red blood cell distribution width |
| 90 | 993501000000105 | 201207 | RBC (red blood cell) distribution width |
| 91 | 1000811000000100 | 194578 | Serum total protein |
| 92 | 1011481000000100 | 192330 | eGFR (estimated glomerular filtration rate) using creatinine Chronic Kidney Disease Epidemiology Collaboration equation per 1.73 square metres |
| 93 | 713636003 | 175318 | Frailty Index score |
| 94 | 1005681000000100 | 174486 | Serum high density lipoprotein cholesterol level |
| 95 | 1005681000000100 | 174486 | Serum HDL (high density lipoprotein) cholesterol level |
| 96 | 270426007 | 172311 | Did not attend - no reason |
| 97 | 270426007 | 172311 | DNA - Did not attend - no reason |
| 98 | 415974002 | 167484 | Tympanic temperature |
| 99 | 1022791000000100 | 166360 | Serum TSH (thyroid stimulating hormone) level |
| 100 | 1005671000000100 | 163080 | Serum cholesterol NOS |

Supplementary Table 2 Top 100 unmapped product codes

|  | ProdCodeId | Term.from.EMIS | DrugIssues |
| --- | --- | --- | --- |
| 1 | 1572871000006117 | Awaiting clinical code migration to EMIS Web | 30000000 |
| 2 | 294711000000118 | Transfer-degraded medication entry | 6000000 |
| 3 | 619841000033115 | Fybogel Granules 3.5 grams/sachet | 2000000 |
| 4 | 643541000033114 | Glyceryl Trinitrate Spray 400 micrograms/dose | 2000000 |
| 5 | 1274141000033118 | Senna Oral Solution, Sugar Free 7.5 mg/5 ml | 2000000 |
| 6 | 2295041000033110 | Doublebase Gel (Pump Dispenser) | 2000000 |
| 7 | 2750641000033112 | Cetraben Emollient Cream | 2000000 |
| 8 | 783241000033112 | Ispaghula Husk Sachets (orange) 3.5 g/sachet | 1000000 |
| 9 | 1023241000033115 | Oxygen Cylinder 1360 litres | 1000000 |
| 10 | 1413741000033113 | Temazepam Capsules 10 mg | 1000000 |
| 11 | 1426941000033114 | Terfenadine 60mg tablets | 1000000 |
| 12 | 1843241000033118 | Olive oil ear drops | 1000000 |
| 13 | 2147541000033115 | Rosiglitazone 4mg tablets | 1000000 |
| 14 | 3232941000033115 | Liquid Paraffin And Isopropyl Myristate Gel (Pump Dispenser) 15 % + 15 % | 1000000 |
| 15 | 457641000033117 | Dioralyte Oral powder | 900000 |
| 16 | 783141000033117 | Ispaghula Husk Sachets 3.5 g/sachet | 900000 |
| 17 | 1803941000033119 | Rofecoxib 12.5mg tablets | 900000 |
| 18 | 1514841000033110 | Vitamin Capsules Bpc Capsules | 800000 |
| 19 | 4036141000033112 | Sharpsafe Container 1 litre | 800000 |
| 20 | 941541000033112 | Multivitamin Capsules | 700000 |
| 21 | 1431841000033111 | Thick And Easy Powder | 700000 |
| 22 | 1433641000033111 | Thioridazine 10mg tablets | 700000 |
| 23 | 1433841000033112 | Thioridazine 25mg tablets | 700000 |
| 24 | 1804041000033117 | Rofecoxib 25mg tablets | 700000 |
| 25 | 2221841000033111 | Advantage Ii Test strips | 700000 |
| 26 | 3996241000033117 | Olanzapine 20mg tablets | 700000 |
| 27 | 7687241000033110 | Sitagliptin 50mg tablets | 700000 |
| 28 | 468841000033112 | Diltiazem Hydrochloride Tablets 60 mg | 600000 |
| 29 | 621541000033110 | Gamolenic acid 40mg capsules | 600000 |
| 30 | 1470941000033111 | Triludan Tablets 60 mg | 600000 |
| 31 | 2571841000033116 | Cavilon Durable Barrier cream 3392E | 600000 |
| 32 | 2720841000033114 | Ispaghula Husk Sugar and Gluten Free Effervescent granules (orange) 3.5 grams/sachet | 600000 |
| 33 | 3159241000033117 | Dermol Cream 500 gram bottle | 600000 |
| 34 | 3196841000033116 | Fortisip Bottle Liquid Feed (Mixed Flavours) Bottle 200 ml | 600000 |
| 35 | 133841000033111 | Beconase Nasal spray 50 micrograms/dose | 500000 |
| 36 | 188241000033114 | Calcipotriol 50micrograms/g cream | 500000 |
| 37 | 444941000033119 | Disposable Insulin syringe with needle 0.5 ml | 500000 |
| 38 | 590341000033112 | Flucloxacillin Oral suspension 125 mg/5 ml | 500000 |
| 39 | 647341000033113 | Glucotrend Test strips | 500000 |
| 40 | 930641000033116 | Monomax Sr 60 M/R capsules 60 mg | 500000 |
| 41 | 974041000033118 | Nitrolingual Spray 400 micrograms/dose | 500000 |
| 42 | 1043841000033118 | Pasteur Merieux Inactivated Influenza Vaccine 0.5 ml | 500000 |
| 43 | 1413941000033111 | Temazepam Capsules 20 mg | 500000 |
| 44 | 2147641000033119 | Rosiglitazone 8mg tablets | 500000 |
| 45 | 4438341000033116 | Ensure Plus Liquid feed (mixed flavours) Milkshake Style | 500000 |
| 46 | 196441000033112 | Calpol Paediatric Suspension 120 mg/5 ml | 400000 |
| 47 | 287341000033117 | Clarityn Tablets 10 mg | 400000 |
| 48 | 586341000033110 | Fluticasone Propionate Inhaler 250 micrograms/puff | 400000 |
| 49 | 596541000033119 | Fluzone Vaccine 0.5 ml | 400000 |
| 50 | 652141000033110 | Graduated Compression Hosiery below knee class 2 | 400000 |
| 51 | 726341000033119 | Human Mixtard 30 Penfill cartridges (3 ml) | 400000 |
| 52 | 792241000033116 | Juvela gluten free loaf sliced (Hero UK Ltd) | 400000 |
| 53 | 906041000033116 | Mfv-Ject Prefilled syringe | 400000 |
| 54 | 1252341000033111 | Salbutamol Rotacaps 400 micrograms | 400000 |
| 55 | 1358941000033119 | Sodium Valproate Sugar-free liquid 200 mg/5 ml | 400000 |
| 56 | 1804941000033116 | Ensure Plus Liquid Feed (Mixed Flavours) Tetrapak 220 ml | 400000 |
| 57 | 2207041000033119 | Glyceryl Trinitrate Cfc-free pump spray 400 micrograms/dose (180 dose) | 400000 |
| 58 | 3232341000033119 | White Soft Paraffin And Liquid Paraffin Light Cream (Pump Dispenser) 13.2 % + 10.5 % | 400000 |
| 59 | 5733041000033111 | Hypodermic insulin needles for pre-filled / reusable pen injectors screw on 4mm/32gauge | 400000 |
| 60 | 6381541000033111 | Vita-Pos Eye ointment (preservative-free) | 400000 |
| 61 | 288341000033118 | Clostet Vaccine | 300000 |
| 62 | 421741000033116 | Depo-Provera Injection 150 mg/1 ml | 300000 |
| 63 | 497541000033112 | Efamast 40 Capsules 40 mg | 300000 |
| 64 | 525441000033115 | Epogam Capsules 40 mg | 300000 |
| 65 | 644241000033114 | Glycerol Suppositories | 300000 |
| 66 | 651841000033113 | Graduated Compression Hosiery thigh length class 2 | 300000 |
| 67 | 773641000033116 | Ipratropium Bromide Nebuliser solution 250 micrograms/ml | 300000 |
| 68 | 791141000033115 | Juvela gluten free fibre loaf sliced (Hero UK Ltd) | 300000 |
| 69 | 791741000033116 | Juvela gluten free mix (Hero UK Ltd) | 300000 |
| 70 | 943141000033116 | Mucaine Suspension | 300000 |
| 71 | 970441000033115 | Nicotine Inhalation cartridge with mouthpiece (refill) 10 mg/cartridge | 300000 |
| 72 | 1044341000033113 | Peppermint Oil Capsules 0.2 ml | 300000 |
| 73 | 1064641000033111 | Penicillin Vk Tablets 250 mg | 300000 |
| 74 | 1264041000033110 | Scanpor Adhesive tape 2.5 cm x 5 m | 300000 |
| 75 | 1420541000033118 | Tetavax Injection | 300000 |
| 76 | 1420641000033117 | Tetavax Injection | 300000 |
| 77 | 1433941000033116 | Thioridazine 50mg tablets | 300000 |
| 78 | 1477641000033115 | Tubigrip Elasticated Support Bandage Stockinette 8.75 cm x 1 m (e) | 300000 |
| 79 | 1618941000033118 | Instillagel Gel (11 Ml Syringe) | 300000 |
| 80 | 1619041000033110 | Instillagel Gel (6 Ml Syringe) | 300000 |
| 81 | 1728741000033117 | Elasticated Viscose Stockinette 10.75 cm x 5 m (yellow line) | 300000 |
| 82 | 1728941000033119 | Elasticated Viscose Stockinette 7.5 cm x 5 m (blue line) | 300000 |
| 83 | 1754341000033113 | Tubifast 2-Way Stretch Stockinette 7.5 cm x 5 m (blue line) | 300000 |
| 84 | 2206941000033115 | Magnesium Hydroxide Mixture BP | 300000 |
| 85 | 2207241000033110 | Nutriprem 2 Powder 900 grams | 300000 |
| 86 | 2271241000033111 | Carmellose Sodium Eye-drops (unit dose) 1 % | 300000 |
| 87 | 2527941000033118 | Penfine Needles for insulin pens 31g, 8 mm | 300000 |
| 88 | 3160241000033116 | Sodium Alginate And Potassium Bicarbonate Oral Suspension Sugar Free Peppermint, 500 mg + 100 mg/5 ml | 300000 |
| 89 | 3169041000033119 | Thick And Easy Powder 225 gram tin | 300000 |
| 90 | 3335541000033112 | Senna 15mg tablets | 300000 |
| 91 | 3346841000033117 | Losartan 100mg / Hydrochlorothiazide 25mg tablets | 300000 |
| 92 | 3925841000033118 | Juvela gluten free fresh white loaf sliced (Hero UK Ltd) | 300000 |
| 93 | 4387641000033111 | Pregabalin 225mg capsules | 300000 |
| 94 | 4898841000033112 | Tadalafil 5mg tablets | 300000 |
| 95 | 5072541000033112 | Fortisip Bottle (Flavour Not Specified) | 300000 |
| 96 | 5234741000033116 | Ensure Plus milkshake style liquid (Flavour Not Specified) | 300000 |
| 97 | 5734741000033112 | Fortisip Compact liquid (Flavour Not Specified) | 300000 |
| 98 | 16141000033110 | Adsorbed Tetanus Vaccine Bp Injection | 200000 |
| 99 | 51641000033119 | Amphotericin B 10mg lozenges sugar free | 200000 |
| 100 | 87341000033112 | Asacol E/c tablets 400 mg | 200000 |

Supplementary Table 3 Bias Information for COVID-19 hospitalisations, with triple therapy users.

|  | variable |  | e1c1 | e0c1 | e1c0 | e0c0 | d1c1 | d0c1 | d1c0 | d0c0 | rrCE | rrCD | absLogBias |
| --- | --- | --- | --- | --- | --- | --- | --- | --- | --- | --- | --- | --- | --- |
| 1 | d2_030700_once | Mucolytics | 11152 | 2437 | 44877 | 19870 | 199 | 13390 | 463 | 64284 | 1.822 | 2.048 | 0.081 |
| 2 | d3_J44_freq | Other chronic obstructive pulmonary disease | 6276 | 1838 | 49753 | 20469 | 231 | 7883 | 431 | 69791 | 1.359 | 4.638 | 0.080 |
| 3 | d3_J44_spor | Other chronic obstructive pulmonary disease | 10472 | 3248 | 45557 | 19059 | 295 | 13425 | 367 | 64249 | 1.284 | 3.786 | 0.079 |
| 4 | d2_030700_spor | Mucolytics | 6438 | 1213 | 49591 | 21094 | 131 | 7520 | 531 | 70154 | 2.113 | 2.279 | 0.070 |
| 5 | d3_Z86_spor | Personal history of certain other diseases | 6277 | 1949 | 49752 | 20358 | 229 | 7997 | 433 | 69677 | 1.282 | 4.508 | 0.064 |
| 6 | d3_J44_once | Other chronic obstructive pulmonary disease | 17904 | 6016 | 38125 | 16291 | 370 | 23550 | 292 | 54124 | 1.185 | 2.883 | 0.060 |
| 7 | d3_J18_once | Pneumonia, organism unspecified | 2965 | 819 | 53064 | 21488 | 133 | 3651 | 529 | 74023 | 1.441 | 4.953 | 0.054 |
| 8 | d3_Z86_freq | Personal history of certain other diseases | 3703 | 1159 | 52326 | 21148 | 173 | 4689 | 489 | 72985 | 1.272 | 5.346 | 0.049 |
| 9 | d2_050101_spor | Penicillins | 15021 | 3909 | 41008 | 18398 | 222 | 18708 | 440 | 58966 | 1.530 | 1.583 | 0.048 |
| 10 | d3_J18_spor | Pneumonia, organism unspecified | 2036 | 548 | 53993 | 21759 | 106 | 2478 | 556 | 75196 | 1.479 | 5.589 | 0.047 |
| 11 | d2_060302_spor | Glucocorticoid therapy | 18726 | 3976 | 37303 | 18331 | 231 | 22471 | 431 | 55203 | 1.875 | 1.313 | 0.045 |
| 12 | d1_F17_once | Mental and behavioural disorders due to use of tobacco | 18384 | 8620 | 37645 | 13687 | 114 | 26890 | 548 | 50784 | 0.849 | 0.395 | 0.045 |
| 13 | d3_Z86_once | Personal history of certain other diseases | 10076 | 3442 | 45953 | 18865 | 275 | 13243 | 387 | 64431 | 1.165 | 3.407 | 0.044 |
| 14 | d2_050101_freq | Penicillins | 8735 | 1900 | 47294 | 20407 | 137 | 10498 | 525 | 67176 | 1.830 | 1.661 | 0.043 |
| 15 | d2_050401_once | Antimalarials | 20076 | 5445 | 35953 | 16862 | 269 | 25252 | 393 | 52422 | 1.468 | 1.417 | 0.042 |
| 16 | d3_J18_freq | Pneumonia, organism unspecified | 1037 | 261 | 54992 | 22046 | 77 | 1221 | 585 | 76453 | 1.582 | 7.812 | 0.042 |
| 17 | d2_030700_freq | Mucolytics | 3016 | 550 | 53013 | 21757 | 70 | 3496 | 592 | 74178 | 2.183 | 2.479 | 0.041 |
| 18 | d2_020202_once | Loop diuretics | 9778 | 3315 | 46251 | 18992 | 254 | 12839 | 408 | 64835 | 1.174 | 3.102 | 0.041 |
| 19 | d2_060302_freq | Glucocorticoid therapy | 8738 | 1547 | 47291 | 20760 | 122 | 10163 | 540 | 67511 | 2.249 | 1.495 | 0.041 |
| 20 | d2_060302_once | Glucocorticoid therapy | 28916 | 7628 | 27113 | 14679 | 344 | 36200 | 318 | 41474 | 1.509 | 1.237 | 0.037 |
| 21 | d2_020202_spor | Loop diuretics | 4940 | 1619 | 51089 | 20688 | 168 | 6391 | 494 | 71283 | 1.215 | 3.722 | 0.035 |
| 22 | d2_030101_spor | Adrenoceptor agonists | 30352 | 8636 | 25677 | 13671 | 365 | 38623 | 297 | 39051 | 1.399 | 1.240 | 0.033 |
| 23 | d3_Z50_once | Care involving use of rehabilitation procedures | 2603 | 796 | 53426 | 21511 | 113 | 3286 | 549 | 74388 | 1.302 | 4.538 | 0.033 |
| 24 | d3_Z50_spor | Care involving use of rehabilitation procedures | 1746 | 508 | 54283 | 21799 | 91 | 2163 | 571 | 75511 | 1.368 | 5.379 | 0.033 |
| 25 | d2_090604_once | Vitamin D | 12476 | 4135 | 43553 | 18172 | 236 | 16375 | 426 | 61299 | 1.201 | 2.059 | 0.032 |
| 26 | d2_090604_spor | Vitamin D | 6584 | 2077 | 49445 | 20230 | 156 | 8505 | 506 | 69169 | 1.262 | 2.480 | 0.031 |
| 27 | d1_F17_spor | Mental and behavioural disorders due to use of tobacco | 11329 | 5467 | 44700 | 16840 | 62 | 16734 | 600 | 60940 | 0.825 | 0.379 | 0.031 |
| 28 | d3_R29_once | Other symptoms and signs involving the nervous and musculoskeletal systems | 1497 | 443 | 54532 | 21864 | 85 | 1855 | 577 | 75819 | 1.345 | 5.801 | 0.030 |
| 29 | d3_R29_spor | Other symptoms and signs involving the nervous and musculoskeletal systems | 989 | 274 | 55040 | 22033 | 68 | 1195 | 594 | 76479 | 1.437 | 6.986 | 0.029 |
| 30 | d2_050401_spor | Antimalarials | 11722 | 2631 | 44307 | 19676 | 153 | 14200 | 509 | 63474 | 1.774 | 1.340 | 0.029 |

* e1c1 = number of exposed (ICS) with the covariate, e0c1 = number unexposed with the covariate, e1c0 = number exposed without the covariate, e0c0 = number exposed without the covariate, d1c1 = number with outcome (COVID-19 hospitalisation) with the covariate, d0c1 = number without the outcome with the covariate, d1c0 = number with the outcome without the covariate, d0c0 = number without the outcome without the covariate, rrCE = relative risk between covariate and exposure, rrCD = relative risk between covariate and outcome, absLogBias is calculated using the Bross formula. In the variable column, d1 refers to primary care clinical observations, d2 to primary care prescriptions, and d3 to hospital data.

Supplementary Table 4 Bias Information for COVID-19 hospitalisations, without triple therapy users

|  | variable |  | e1c1 | e0c1 | e1c0 | e0c0 | d1c1 | d0c1 | d1c0 | d0c0 | rrCE | rrCD | absLogBias |
| --- | --- | --- | --- | --- | --- | --- | --- | --- | --- | --- | --- | --- | --- |
| 1 | d1_F17_once | Mental and behavioural disorders due to use of tobacco | 4609 | 8620 | 10296 | 13687 | 38 | 13191 | 202 | 23781 | 0.800 | 0.341 | 0.066 |
| 2 | d1_F17_spor | Mental and behavioural disorders due to use of tobacco | 2706 | 5467 | 12199 | 16840 | 15 | 8158 | 225 | 28814 | 0.741 | 0.237 | 0.058 |
| 3 | d1_Z71_once | Persons encountering health services for other counselling and medical advice, not elsewhere classified | 7168 | 12102 | 7737 | 10205 | 79 | 19191 | 161 | 17781 | 0.886 | 0.457 | 0.046 |
| 4 | d1_Z71_spor | Persons encountering health services for other counselling and medical advice, not elsewhere classified | 3417 | 6334 | 11488 | 15973 | 27 | 9724 | 213 | 27248 | 0.807 | 0.357 | 0.042 |
| 5 | d3_R29_spor | Other symptoms and signs involving the nervous and musculoskeletal systems | 264 | 274 | 14641 | 22033 | 23 | 515 | 217 | 36457 | 1.442 | 7.225 | 0.031 |
| 6 | d3_R29_once | Other symptoms and signs involving the nervous and musculoskeletal systems | 405 | 443 | 14500 | 21864 | 28 | 820 | 212 | 36152 | 1.368 | 5.664 | 0.031 |
| 7 | d3_N39_once | Other disorders of urinary system | 342 | 365 | 14563 | 21942 | 24 | 683 | 216 | 36289 | 1.402 | 5.737 | 0.029 |
| 8 | d1_F17_freq | Mental and behavioural disorders due to use of tobacco | 1386 | 2930 | 13519 | 19377 | 10 | 4306 | 230 | 32666 | 0.708 | 0.331 | 0.028 |
| 9 | d1_J42_once | Unspecified chronic bronchitis | 1413 | 1703 | 13492 | 20604 | 47 | 3069 | 193 | 33903 | 1.242 | 2.665 | 0.027 |
| 10 | d3_J96_once | Respiratory failure, not elsewhere classified | 205 | 477 | 14700 | 21830 | 19 | 663 | 221 | 36309 | 0.643 | 4.605 | 0.026 |
| 11 | d2_030102_once | Antimuscarinic bronchodilators | 583 | 140 | 14322 | 22167 | 8 | 715 | 232 | 36257 | 6.232 | 1.740 | 0.024 |
| 12 | d3_W19_once | Unspecified fall | 172 | 168 | 14733 | 22139 | 15 | 325 | 225 | 36647 | 1.532 | 7.230 | 0.024 |
| 13 | d1_J44_once | Other chronic obstructive pulmonary disease | 4481 | 8080 | 10424 | 14227 | 61 | 12500 | 179 | 24472 | 0.830 | 0.669 | 0.023 |
| 14 | d2_010602_freq | Stimulant laxatives | 276 | 299 | 14629 | 22008 | 20 | 555 | 220 | 36417 | 1.381 | 5.792 | 0.023 |
| 15 | d2_050101_freq | Penicillins | 3030 | 3909 | 11875 | 18398 | 74 | 6865 | 166 | 30107 | 1.160 | 1.945 | 0.022 |
| 16 | d1_J42_freq | Unspecified chronic bronchitis | 391 | 425 | 14514 | 21882 | 21 | 795 | 219 | 36177 | 1.377 | 4.277 | 0.022 |
| 17 | d2_010602_spor | Stimulant laxatives | 499 | 590 | 14406 | 21717 | 28 | 1061 | 212 | 35911 | 1.266 | 4.381 | 0.022 |
| 18 | d3_F03_once | Unspecified dementia | 146 | 111 | 14759 | 22196 | 9 | 248 | 231 | 36724 | 1.969 | 5.602 | 0.021 |
| 19 | d1_Z75_once | Problems related to medical facilities and other health care | 9 | 2 | 14896 | 22305 | 3 | 8 | 237 | 36964 | 6.735 | 42.809 | 0.021 |
| 20 | d2_090604_freq | Vitamin D | 972 | 1261 | 13933 | 21046 | 46 | 2187 | 194 | 34785 | 1.154 | 3.714 | 0.020 |
| 21 | d3_N39_spor | Other disorders of urinary system | 182 | 188 | 14723 | 22119 | 15 | 355 | 225 | 36617 | 1.449 | 6.638 | 0.020 |
| 22 | d3_J45_once | Asthma | 547 | 296 | 14358 | 22011 | 10 | 833 | 230 | 36139 | 2.766 | 1.876 | 0.020 |
| 23 | d2_021200_once | Lipid-regulating drugs | 7650 | 12214 | 7255 | 10093 | 163 | 19701 | 77 | 17271 | 0.937 | 1.849 | 0.020 |
| 24 | d2_090604_spor | Vitamin D | 1572 | 2077 | 13333 | 20230 | 58 | 3591 | 182 | 33381 | 1.133 | 2.931 | 0.020 |
| 25 | d3_K59_once | Other functional intestinal disorders | 300 | 344 | 14605 | 21963 | 21 | 623 | 219 | 36349 | 1.305 | 5.445 | 0.019 |
| 26 | d1_E94_once | Bronchial tests | 1641 | 3355 | 13264 | 18952 | 19 | 4977 | 221 | 31995 | 0.732 | 0.554 | 0.019 |
| 27 | d2_010602_once | Stimulant laxatives | 919 | 1213 | 13986 | 21094 | 46 | 2086 | 194 | 34886 | 1.134 | 3.901 | 0.018 |
| 28 | d2_010604_once | Osmotic laxatives | 1804 | 2425 | 13101 | 19882 | 62 | 4167 | 178 | 32805 | 1.113 | 2.717 | 0.018 |
| 29 | d1_W19_once | Unspecified fall | 444 | 515 | 14461 | 21792 | 22 | 937 | 218 | 36035 | 1.290 | 3.815 | 0.018 |
| 30 | d3_I50_spor | Heart failure | 349 | 627 | 14556 | 21680 | 29 | 947 | 211 | 36025 | 0.833 | 5.103 | 0.017 |

* e1c1 = number of exposed (ICS) with the covariate, e0c1 = number of unexposed with the covariate, e1c0 = number exposed without the covariate, e0c0 = number exposed without the covariate, d1c1 = number with outcome (COVID-19 hospitalisation) with the covariate, d0c1 = number without the outcome with the covariate, d1c0 = number with the outcome without the covariate, d0c0 = number without the outcome without the covariate, rrCE = relative risk between covariate and exposure, rrCD = relative risk between covariate and outcome, absLogBias is calculated using the Bross formula. In the variable column, d1 refers to primary care clinical observations, d2 to primary care prescriptions, and d3 to hospital data.

Supplementary Table 5 Bias Information for COVID-19 deaths, with triple therapy users

|  | variable |  | e1c1 | e0c1 | e1c0 | e0c0 | d1c1 | d0c1 | d1c0 | d0c0 | rrCE | rrCD | absLogBias |
| --- | --- | --- | --- | --- | --- | --- | --- | --- | --- | --- | --- | --- | --- |
| 1 | d3_J44_freq | Other chronic obstructive pulmonary disease | 6276 | 1838 | 49753 | 20469 | 126 | 7988 | 240 | 69982 | 1.359 | 4.544 | 0.078 |
| 2 | d3_J44_spor | Other chronic obstructive pulmonary disease | 10472 | 3248 | 45557 | 19059 | 159 | 13561 | 207 | 64409 | 1.284 | 3.618 | 0.075 |
| 3 | d3_J44_once | Other chronic obstructive pulmonary disease | 17904 | 6016 | 38125 | 16291 | 204 | 23716 | 162 | 54254 | 1.185 | 2.865 | 0.060 |
| 4 | d3_J18_once | Pneumonia, organism unspecified | 2965 | 819 | 53064 | 21488 | 78 | 3706 | 288 | 74264 | 1.441 | 5.336 | 0.059 |
| 5 | d3_Z86_spor | Personal history of certain other diseases | 6277 | 1949 | 49752 | 20358 | 116 | 8110 | 250 | 69860 | 1.282 | 3.955 | 0.056 |
| 6 | d3_J18_spor | Pneumonia, organism unspecified | 2036 | 548 | 53993 | 21759 | 65 | 2519 | 301 | 75451 | 1.479 | 6.331 | 0.054 |
| 7 | d2_030101_spor | Adrenoceptor agonists | 30352 | 8636 | 25677 | 13671 | 213 | 38775 | 153 | 39195 | 1.399 | 1.405 | 0.053 |
| 8 | d3_Z86_freq | Personal history of certain other diseases | 3703 | 1159 | 52326 | 21148 | 96 | 4766 | 270 | 73204 | 1.272 | 5.373 | 0.049 |
| 9 | d3_J18_freq | Pneumonia, organism unspecified | 1037 | 261 | 54992 | 22046 | 48 | 1250 | 318 | 76720 | 1.582 | 8.959 | 0.048 |
| 10 | d2_030700_spor | Mucolytics | 6438 | 1213 | 49591 | 21094 | 61 | 7590 | 305 | 70380 | 2.113 | 1.848 | 0.048 |
| 11 | d2_030700_once | Mucolytics | 11152 | 2437 | 44877 | 19870 | 91 | 13498 | 275 | 64472 | 1.822 | 1.577 | 0.048 |
| 12 | d2_030700_freq | Mucolytics | 3016 | 550 | 53013 | 21757 | 42 | 3524 | 324 | 74446 | 2.183 | 2.718 | 0.047 |
| 13 | d2_090604_once | Vitamin D | 12476 | 4135 | 43553 | 18172 | 151 | 16460 | 215 | 61510 | 1.201 | 2.610 | 0.045 |
| 14 | d2_090604_spor | Vitamin D | 6584 | 2077 | 49445 | 20230 | 106 | 8555 | 260 | 69415 | 1.262 | 3.280 | 0.045 |
| 15 | d2_020202_once | Loop diuretics | 9778 | 3315 | 46251 | 18992 | 147 | 12946 | 219 | 65024 | 1.174 | 3.345 | 0.044 |
| 16 | d2_050101_spor | Penicillins | 15021 | 3909 | 41008 | 18398 | 119 | 18811 | 247 | 59159 | 1.530 | 1.512 | 0.043 |
| 17 | d1_F17_once | Mental and behavioural disorders due to use of tobacco | 18384 | 8620 | 37645 | 13687 | 67 | 26937 | 299 | 51033 | 0.849 | 0.426 | 0.042 |
| 18 | d2_090604_freq | Vitamin D | 4052 | 1261 | 51977 | 21046 | 84 | 5229 | 282 | 72741 | 1.279 | 4.094 | 0.041 |
| 19 | d2_020202_spor | Loop diuretics | 4940 | 1619 | 51089 | 20688 | 103 | 6456 | 263 | 71514 | 1.215 | 4.286 | 0.041 |
| 20 | d3_Z86_once | Personal history of certain other diseases | 10076 | 3442 | 45953 | 18865 | 144 | 13374 | 222 | 64596 | 1.165 | 3.110 | 0.040 |
| 21 | d2_030101_freq | Adrenoceptor agonists | 15018 | 3715 | 41011 | 18592 | 113 | 18620 | 253 | 59350 | 1.609 | 1.421 | 0.039 |
| 22 | d3_R29_spor | Other symptoms and signs involving the nervous and musculoskeletal systems | 989 | 274 | 55040 | 22033 | 47 | 1216 | 319 | 76754 | 1.437 | 8.991 | 0.038 |
| 23 | d3_R29_once | Other symptoms and signs involving the nervous and musculoskeletal systems | 1497 | 443 | 54532 | 21864 | 57 | 1883 | 309 | 76087 | 1.345 | 7.264 | 0.038 |
| 24 | d1_E94_once | Bronchial tests | 5620 | 3355 | 50409 | 18952 | 16 | 8959 | 350 | 69011 | 0.667 | 0.353 | 0.035 |
| 25 | d3_Z50_once | Care involving use of rehabilitation procedures | 2603 | 796 | 53426 | 21511 | 64 | 3335 | 302 | 74635 | 1.302 | 4.672 | 0.034 |
| 26 | d2_010604_once | Osmotic laxatives | 7191 | 2425 | 48838 | 19882 | 110 | 9506 | 256 | 68464 | 1.181 | 3.071 | 0.033 |
| 27 | d3_Z50_spor | Care involving use of rehabilitation procedures | 1746 | 508 | 54283 | 21799 | 50 | 2204 | 316 | 75766 | 1.368 | 5.341 | 0.033 |
| 28 | d2_020202_freq | Loop diuretics | 3239 | 1043 | 52790 | 21264 | 74 | 4208 | 292 | 73762 | 1.236 | 4.383 | 0.032 |
| 29 | d3_E87_once | Other disorders of fluid, electrolyte and acid-base balance | 1988 | 607 | 54041 | 21700 | 56 | 2539 | 310 | 75431 | 1.304 | 5.273 | 0.031 |
| 30 | d3_I10_freq | Essential (primary) hypertension | 3525 | 1168 | 52504 | 21139 | 82 | 4611 | 284 | 73359 | 1.202 | 4.531 | 0.031 |

* e1c1 = number exposed (ICS) with the covariate, e0c1 = number of unexposed with the covariate, e1c0 = number exposed without the covariate, e0c0 = number exposed without the covariate, d1c1 = number with outcome (COVID-19 death) with the covariate, d0c1 = number without the outcome with the covariate, d1c0 = number with the outcome without the covariate, d0c0 = number without the outcome without the covariate, rrCE = relative risk between covariate and exposure, rrCD = relative risk between covariate and outcome, absLogBias is calculated using the Bross formula. In the variable column, d1 refers to primary care clinical observations, d2 to primary care prescriptions, and d3 to hospital data.

Supplementary Table 6 Bias Information for COVID-19 deaths, without triple therapy users

|  | variable |  | e1c1 | e0c1 | e1c0 | e0c0 | d1c1 | d0c1 | d1c0 | d0c0 | rrCE | rrCD | absLogBias |
| --- | --- | --- | --- | --- | --- | --- | --- | --- | --- | --- | --- | --- | --- |
| 1 | d1_F17_once | Mental and behavioural disorders due to use of tobacco | 4609 | 8620 | 10296 | 13687 | 20 | 13209 | 114 | 23869 | 0.800 | 0.318 | 0.069 |
| 2 | d1_Z71_once | Persons encountering health services for other counselling and medical advice, not elsewhere classified | 7168 | 12102 | 7737 | 10205 | 37 | 19233 | 97 | 17845 | 0.886 | 0.355 | 0.059 |
| 3 | d1_F17_spor | Mental and behavioural disorders due to use of tobacco | 2706 | 5467 | 12199 | 16840 | 9 | 8164 | 125 | 28914 | 0.741 | 0.256 | 0.056 |
| 4 | d3_R29_once | Other symptoms and signs involving the nervous and musculoskeletal systems | 405 | 443 | 14500 | 21864 | 21 | 827 | 113 | 36251 | 1.368 | 7.969 | 0.044 |
| 5 | d2_041100_once | Drugs for dementia | 238 | 258 | 14667 | 22049 | 19 | 477 | 115 | 36601 | 1.381 | 12.230 | 0.043 |
| 6 | d3_R29_spor | Other symptoms and signs involving the nervous and musculoskeletal systems | 264 | 274 | 14641 | 22033 | 17 | 521 | 117 | 36557 | 1.442 | 9.905 | 0.043 |
| 7 | d2_041100_spor | Drugs for dementia | 135 | 128 | 14770 | 22179 | 13 | 250 | 121 | 36828 | 1.578 | 15.094 | 0.042 |
| 8 | d3_F03_once | Unspecified dementia | 146 | 111 | 14759 | 22196 | 9 | 248 | 125 | 36830 | 1.969 | 10.353 | 0.042 |
| 9 | d1_Z71_spor | Persons encountering health services for other counselling and medical advice, not elsewhere classified | 3417 | 6334 | 11488 | 15973 | 16 | 9735 | 118 | 27343 | 0.807 | 0.382 | 0.040 |
| 10 | d3_J45_once | Asthma | 547 | 296 | 14358 | 22011 | 8 | 835 | 126 | 36243 | 2.766 | 2.739 | 0.039 |
| 11 | d2_091316_once | Thickener | 75 | 54 | 14830 | 22253 | 7 | 122 | 127 | 36956 | 2.079 | 15.845 | 0.037 |
| 12 | d2_090604_freq | Vitamin D | 972 | 1261 | 13933 | 21046 | 38 | 2195 | 96 | 34883 | 1.154 | 6.201 | 0.034 |
| 13 | d2_090604_spor | Vitamin D | 1572 | 2077 | 13333 | 20230 | 46 | 3603 | 88 | 33475 | 1.133 | 4.808 | 0.034 |
| 14 | d1_E94_once | Bronchial tests | 1641 | 3355 | 13264 | 18952 | 5 | 4991 | 129 | 32087 | 0.732 | 0.250 | 0.034 |
| 15 | d3_N39_once | Other disorders of urinary system | 342 | 365 | 14563 | 21942 | 15 | 692 | 119 | 36386 | 1.402 | 6.508 | 0.033 |
| 16 | d3_G30_once | Alzheimer disease | 78 | 68 | 14827 | 22239 | 8 | 138 | 126 | 36940 | 1.717 | 16.119 | 0.031 |
| 17 | d1_J42_once | Unspecified chronic bronchitis | 1413 | 1703 | 13492 | 20604 | 28 | 3088 | 106 | 33990 | 1.242 | 2.890 | 0.030 |
| 18 | d3_F00_once | Dementia in Alzheimer disease | 59 | 49 | 14846 | 22258 | 7 | 101 | 127 | 36977 | 1.802 | 18.936 | 0.030 |
| 19 | d2_050101_freq | Penicillins | 3030 | 3909 | 11875 | 18398 | 46 | 6893 | 88 | 30185 | 1.160 | 2.281 | 0.029 |
| 20 | d3_F03_spor | Unspecified dementia | 89 | 67 | 14816 | 22240 | 6 | 150 | 128 | 36928 | 1.988 | 11.135 | 0.029 |
| 21 | d2_010604_spor | Osmotic laxatives | 1130 | 1443 | 13775 | 20864 | 30 | 2543 | 104 | 34535 | 1.172 | 3.883 | 0.027 |
| 22 | d1_F17_freq | Mental and behavioural disorders due to use of tobacco | 1386 | 2930 | 13519 | 19377 | 6 | 4310 | 128 | 32768 | 0.708 | 0.357 | 0.027 |
| 23 | d2_010602_spor | Stimulant laxatives | 499 | 590 | 14406 | 21717 | 18 | 1071 | 116 | 36007 | 1.266 | 5.147 | 0.026 |
| 24 | d3_W19_once | Unspecified fall | 172 | 168 | 14733 | 22139 | 9 | 331 | 125 | 36747 | 1.532 | 7.808 | 0.026 |
| 25 | d2_010604_once | Osmotic laxatives | 1804 | 2425 | 13101 | 19882 | 43 | 4186 | 91 | 32892 | 1.113 | 3.685 | 0.025 |
| 26 | d2_010602_freq | Stimulant laxatives | 276 | 299 | 14629 | 22008 | 12 | 563 | 122 | 36515 | 1.381 | 6.267 | 0.025 |
| 27 | d2_212200_spor | Emollients | 1046 | 1337 | 13859 | 20970 | 27 | 2356 | 107 | 34722 | 1.171 | 3.688 | 0.023 |
| 28 | d1_E97_once | Respiratory education | 2422 | 5044 | 12483 | 17263 | 19 | 7447 | 115 | 29631 | 0.719 | 0.658 | 0.023 |
| 29 | d2_090604_once | Vitamin D | 2979 | 4135 | 11926 | 18172 | 58 | 7056 | 76 | 30022 | 1.078 | 3.229 | 0.023 |
| 30 | d1_J42_freq | Unspecified chronic bronchitis | 391 | 425 | 14514 | 21882 | 12 | 804 | 122 | 36274 | 1.377 | 4.387 | 0.023 |

* e1c1 = number exposed (ICS) with the covariate, e0c1 = number of unexposed with the covariate, e1c0 = number exposed without the covariate, e0c0 = number exposed without the covariate, d1c1 = number with outcome (COVID-19 death) with the covariate, d0c1 = number without the outcome with the covariate, d1c0 = number with the outcome without the covariate, d0c0 = number without the outcome without the covariate, rrCE = relative risk between covariate and exposure, rrCD = relative risk between covariate and outcome, absLogBias is calculated using the Bross formula. In the variable column, d1 refers to primary care clinical observations, d2 to primary care prescriptions, and d3 to hospital data.

Supplementary Table 7 baseline characteristics (Including triple therapy users)

|  | Unweighted | |  |  | SMDs including HDPS for hospitalisations | | | | | SMDs after HDPS for deaths | | | | |
| --- | --- | --- | --- | --- | --- | --- | --- | --- | --- | --- | --- | --- | --- | --- |
|  | **LABA/LAMA**  **(n = 22318)** | **ICS/LABA**  **(n = 56049)** | **Unweighted SMD** | **SMD (weighted predefined covariates)** | **SMD (100 HDPS covariates)** | **SMD (250 HDPS covariates)** | **SMD (500 HDPS covariates)** | **SMD (750 HDPS covariates)** | **SMD (1000 HDPS covariates)** | **SMD (100 HDPS covariates)** | **SMD (250 HDPS covariates)** | **SMD (500 HDPS covariates)** | **SMD (750 HDPS covariates)** | **SMD (1000 HDPS covariates)** |
| Gender = Female | 10074 (45.1) | 26228 (46.8) | 0.033 | 0.005 | 0.003 | 0.002 | 0.001 | 0.001 | 0.001 | 0.002 | 0.001 | 0.004 | 0.004 | 0.002 |
| age (mean (SD)) | 70.82 (10.23) | 71.32 (10.47) | 0.048 | 0.002 | 0.008 | 0.01 | 0.011 | 0.009 | 0.006 | 0.008 | 0.007 | 0.011 | 0.01 | 0.009 |
| IMD |  |  | 0.044 | 0.005 | 0.009 | 0.008 | 0.006 | 0.006 | 0.007 | 0.009 | 0.008 | 0.007 | 0.008 | 0.007 |
| 1 | 3047 (13.7) | 7178 (12.8) |  |  |  |  |  |  |  |  |  |  |  |  |
| 2 | 3813 (17.1) | 9212 (16.4) |  |  |  |  |  |  |  |  |  |  |  |  |
| 3 | 4077 (18.3) | 9970 (17.8) |  |  |  |  |  |  |  |  |  |  |  |  |
| 4 | 5012 (22.5) | 12719 (22.7) |  |  |  |  |  |  |  |  |  |  |  |  |
| 5 | 6357 (28.5) | 16940 (30.2) |  |  |  |  |  |  |  |  |  |  |  |  |
| Missing | 12 ( 0.1) | 30 ( 0.1) |  |  |  |  |  |  |  |  |  |  |  |  |
| Diabetes | 5517 (24.7) | 14078 (25.1) | 0.009 | 0.003 | 0.008 | 0.01 | 0.013 | 0.012 | 0.013 | 0.008 | 0.011 | 0.012 | 0.011 | 0.012 |
| Hypertension | 11318 (50.7) | 28598 (51.0) | 0.006 | 0.003 | 0.006 | 0.002 | 0.004 | 0.003 | 0.001 | 0.006 | 0.006 | 0.003 | 0.002 | 0.002 |
| Cardiovascular disease | 6539 (29.3) | 16768 (29.9) | 0.014 | 0.002 | 0.004 | 0.003 | 0.004 | 0.003 | 0.002 | 0.005 | 0.005 | 0.008 | 0.007 | 0.005 |
| Cancer | 4415 (19.8) | 10542 (18.8) | 0.025 | 0.001 | 0.004 | 0.005 | 0.006 | 0.006 | 0.003 | 0.005 | 0.008 | 0.01 | 0.011 | 0.01 |
| Past asthma | 2665 (11.9) | 15336 (27.4) | 0.396 | 0.002 | 0.013 | 0.022 | 0.022 | 0.02 | 0.02 | 0.013 | 0.012 | 0.014 | 0.012 | 0.015 |
| Chronic kidney disease | 6737 (30.2) | 16722 (29.8) | 0.008 | 0.007 | 0.012 | 0.012 | 0.013 | 0.011 | 0.009 | 0.013 | 0.012 | 0.013 | 0.012 | 0.012 |
| Immunosuppression | 277 ( 1.2) | 665 ( 1.2) | 0.005 | <0.001 | <0.001 | 0.002 | 0.001 | <0.001 | 0.001 | 0.001 | 0.004 | 0.003 | 0.005 | 0.003 |
| Influenza vaccine | 17961 (80.5) | 44898 (80.1) | 0.009 | 0.001 | <0.001 | <0.001 | <0.001 | 0.002 | 0.005 | 0.001 | 0.002 | 0.003 | 0.001 | 0.003 |
| Pneumococcal vaccine | 3236 (14.5) | 5985 (10.7) | 0.115 | 0.001 | <0.001 | 0.001 | <0.001 | <0.001 | 0.001 | 0.001 | <0.001 | <0.001 | 0.001 | 0.001 |
| COPD exacerbation past 12 months) | 6222 (27.9) | 22799 (40.7) | 0.272 | 0.002 | 0.003 | 0.005 | 0.006 | 0.008 | 0.009 | 0.002 | <0.001 | 0.007 | 0.006 | 0.011 |
| Former smoking | 12246 (54.9) | 33287 (59.4) | 0.091 | 0.002 | 0.01 | 0.01 | 0.009 | 0.008 | 0.007 | 0.009 | 0.008 | 0.006 | 0.008 | 0.008 |
| Ethnicity (%) |  |  | 0.05 | 0.009 | 0.009 | 0.017 | 0.016 | 0.014 | 0.014 | 0.011 | 0.01 | 0.011 | 0.013 | 0.011 |
| White | 19584 (87.7) | 49390 (88.1) |  |  |  |  |  |  |  |  |  |  |  |  |
| South Asian | 197 ( 0.9) | 732 ( 1.3) |  |  |  |  |  |  |  |  |  |  |  |  |
| Black | 130 ( 0.6) | 351 ( 0.6) |  |  |  |  |  |  |  |  |  |  |  |  |
| Mixed | 49 ( 0.2) | 142 ( 0.3) |  |  |  |  |  |  |  |  |  |  |  |  |
| Unknown | 2358 (10.6) | 5434 ( 9.7) |  |  |  |  |  |  |  |  |  |  |  |  |
| BMI category(%) |  |  | 0.07 | 0.004 | 0.007 | 0.008 | 0.008 | 0.006 | 0.007 | 0.007 | 0.009 | 0.007 | 0.006 | 0.006 |
| Underweight (<18.5) | 970 ( 4.3) | 3140 ( 5.6) |  |  |  |  |  |  |  |  |  |  |  |  |
| Normal (18.5-24.9) | 6926 (31.0) | 18155 (32.4) |  |  |  |  |  |  |  |  |  |  |  |  |
| Overweight (25-29.9) | 7171 (32.1) | 17349 (31.0) |  |  |  |  |  |  |  |  |  |  |  |  |
| Obese (>=30) | 7251 (32.5) | 17405 (31.1) |  |  |  |  |  |  |  |  |  |  |  |  |

Supplementary Table 8 baseline characteristics (No triple therapy users)

|  | Unweighted | |  |  | SMDs after HDPS for hospitalisations | | | | | SMDs after HDPS for deaths | | | | |
| --- | --- | --- | --- | --- | --- | --- | --- | --- | --- | --- | --- | --- | --- | --- |
|  | **LABA/LAMA**  **(n = 22308)** | **ICS/LABA**  **(n = 14905)** | **SMD unweighted** | **SMD (predefined covariates)** | **SMD (100 HDPS covariates)** | **SMD (250 HDPS covariates)** | **SMD (500 HDPS covariates)** | **SMD (750 HDPS covariates)** | **SMD (1000 HDPS covariates)** | **SMD (100 HDPS covariates)** | **SMD (250 HDPS covariates)** | **SMD (500 HDPS covariates)** | **SMD (750 HDPS covariates)** | **SMD (1000 HDPS covariates)** |
| Gender = Female | 10068 (45.1) | 7089 (47.6) | 0.049 | 0.001 | 0.001 | 0.003 | 0.005 | 0.006 | 0.006 | <0.001 | 0.002 | 0.003 | 0.006 | 0.005 |
| age (mean (SD)) | 70.83 (10.23) | 71.37 (11.29) | 0.051 | 0.002 | 0.002 | 0.002 | <0.001 | 0.002 | 0.001 | 0.002 | 0.003 | 0.005 | 0.004 | 0.004 |
| IMD |  |  | 0.028 | 0.016 | 0.017 | 0.016 | 0.017 | 0.017 | 0.018 | 0.022 | 0.019 | 0.019 | 0.018 | 0.018 |
| 1 | 3048 (13.7) | 2021 (13.6) |  |  |  |  |  |  |  |  |  |  |  |  |
| 2 | 3813 (17.1) | 2562 (17.2) |  |  |  |  |  |  |  |  |  |  |  |  |
| 3 | 4077 (18.3) | 2759 (18.5) |  |  |  |  |  |  |  |  |  |  |  |  |
| 4 | 5012 (22.5) | 3473 (23.3) |  |  |  |  |  |  |  |  |  |  |  |  |
| 5 | 6357 (28.5) | 4089 (27.4) |  |  |  |  |  |  |  |  |  |  |  |  |
| Missing | 1 ( 0.0) | 1 ( 0.0) |  |  |  |  |  |  |  |  |  |  |  |  |
| Diabetes | 5515 (24.7) | 3685 (24.7) | <0.001 | 0.001 | 0.002 | 0.001 | 0.001 | 0.001 | 0.001 | 0.002 | <0.001 | 0.001 | 0.002 | 0.003 |
| Hypertension | 11313 (50.7) | 7688 (51.6) | 0.017 | 0.001 | 0.001 | 0.001 | <0.001 | 0.002 | 0.002 | <0.001 | 0.001 | 0.001 | 0.003 | 0.004 |
| Cardiovascular disease | 6538 (29.3) | 4387 (29.4) | 0.003 | <0.001 | 0.001 | 0.001 | 0.003 | 0.004 | 0.002 | 0.001 | 0.001 | 0.001 | 0.001 | 0.001 |
| Cancer | 4414 (19.8) | 2827 (19.0) | 0.021 | <0.001 | 0.002 | 0.003 | 0.006 | 0.007 | 0.006 | 0.001 | 0.004 | 0.004 | 0.005 | 0.005 |
| Past asthma | 2665 (11.9) | 4134 (27.7) | 0.404 | <0.001 | <0.001 | 0.001 | 0.002 | 0.005 | 0.006 | <0.001 | 0.001 | 0.001 | 0.002 | 0.004 |
| Kidney disease | 6737 (30.2) | 4566 (30.6) | 0.009 | 0.001 | 0.004 | 0.006 | 0.01 | 0.01 | 0.006 | 0.002 | 0.006 | 0.005 | 0.007 | 0.007 |
| Immunosuppression | 274 ( 1.2) | 188 ( 1.3) | 0.003 | <0.001 | 0.001 | <0.001 | 0.003 | 0.004 | 0.004 | <0.001 | 0.006 | 0.005 | 0.007 | 0.007 |
| Influenza vaccine | 17956 (80.5) | 11390 (76.4) | 0.099 | <0.001 | 0.002 | 0.004 | 0.002 | 0.006 | 0.005 | 0.002 | 0.004 | 0.003 | 0.004 | 0.004 |
| Pneumococcal vaccine | 3236 (14.5) | 1497 (10.0) | 0.136 | <0.001 | 0.002 | 0.002 | 0.002 | 0.002 | 0.001 | 0.003 | 0.001 | <0.001 | 0.002 | 0.002 |
| COPD exacerbation past 12 months) | 6221 (27.9) | 4637 (31.1) | 0.071 | 0.001 | 0.004 | 0.003 | 0.003 | 0.007 | 0.006 | 0.002 | 0.005 | 0.005 | 0.007 | 0.008 |
| Former smoking | 12240 (54.9) | 8941 (60.0) | 0.104 | 0.001 | 0.003 | 0.002 | 0.002 | 0.005 | 0.002 | 0.002 | 0.002 | 0.005 | 0.006 | 0.006 |
| Ethnicity (%) |  |  | 0.101 | 0.002 | 0.012 | 0.014 | 0.013 | 0.013 | 0.015 | 0.004 | 0.004 | 0.005 | 0.005 | 0.005 |
| White | 19575 (87.7) | 12891 (86.5) |  |  |  |  |  |  |  |  |  |  |  |  |
| South Asian | 197 ( 0.9) | 291 ( 2.0) |  |  |  |  |  |  |  |  |  |  |  |  |
| Black | 130 ( 0.6) | 138 ( 0.9) |  |  |  |  |  |  |  |  |  |  |  |  |
| Mixed | 49 ( 0.2) | 44 ( 0.3) |  |  |  |  |  |  |  |  |  |  |  |  |
| Unknown | 2357 (10.6) | 1541 (10.3) |  |  |  |  |  |  |  |  |  |  |  |  |
| BMI category (%) |  |  | 0.038 | 0.001 | 0.004 | 0.004 | 0.005 | 0.006 | 0.005 | 0.003 | 0.003 | 0.001 | 0.003 | 0.002 |
| Underweight (<18.5) | 969 ( 4.3) | 619 ( 4.2) |  |  |  |  |  |  |  |  |  |  |  |  |
| Normal (18.5-24.9) | 6923 (31.0) | 4628 (31.0) |  |  |  |  |  |  |  |  |  |  |  |  |
| Overweight (25-29.9) | 7167 (32.1) | 5021 (33.7) |  |  |  |  |  |  |  |  |  |  |  |  |
| Obese (>=30) | 7249 (32.5) | 4637 (31.1) |  |  |  |  |  |  |  |  |  |  |  |  |

2. Supplementary figures
   2.1. Concept plots for outcome COVID-19 hospitalisation, including triple therapy users

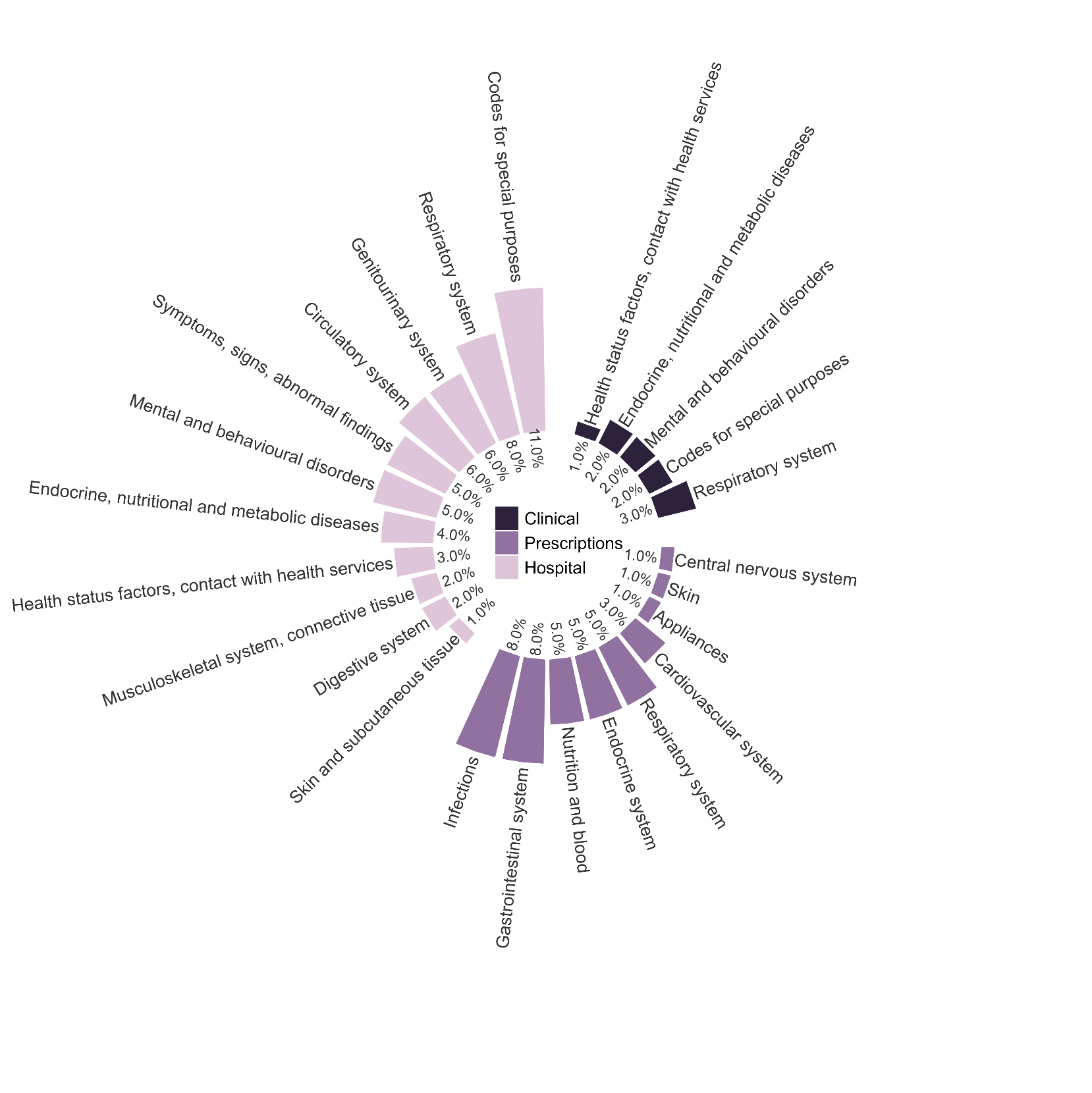

Supplementary Figure 1 Summary of high-level concepts captured in the top 100 ranked high-dimensional propensity score covariates by data dimension for COVID-19 hospitalisations, including triple therapy users

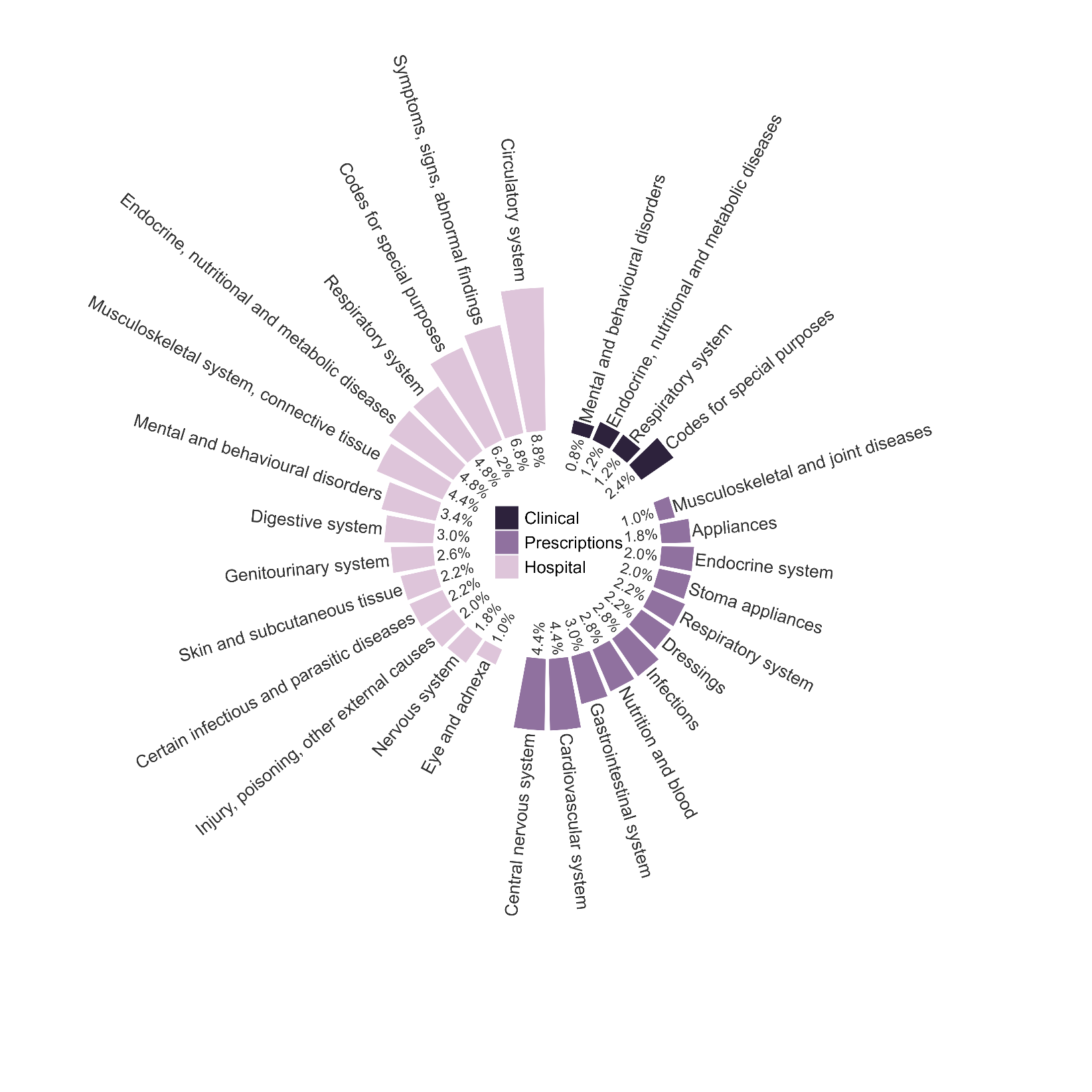

Supplementary Figure 2 Summary of high-level concepts captured in the top 500 ranked high-dimensional propensity score covariates by data dimension for COVID-19 hospitalisations, including triple therapy users

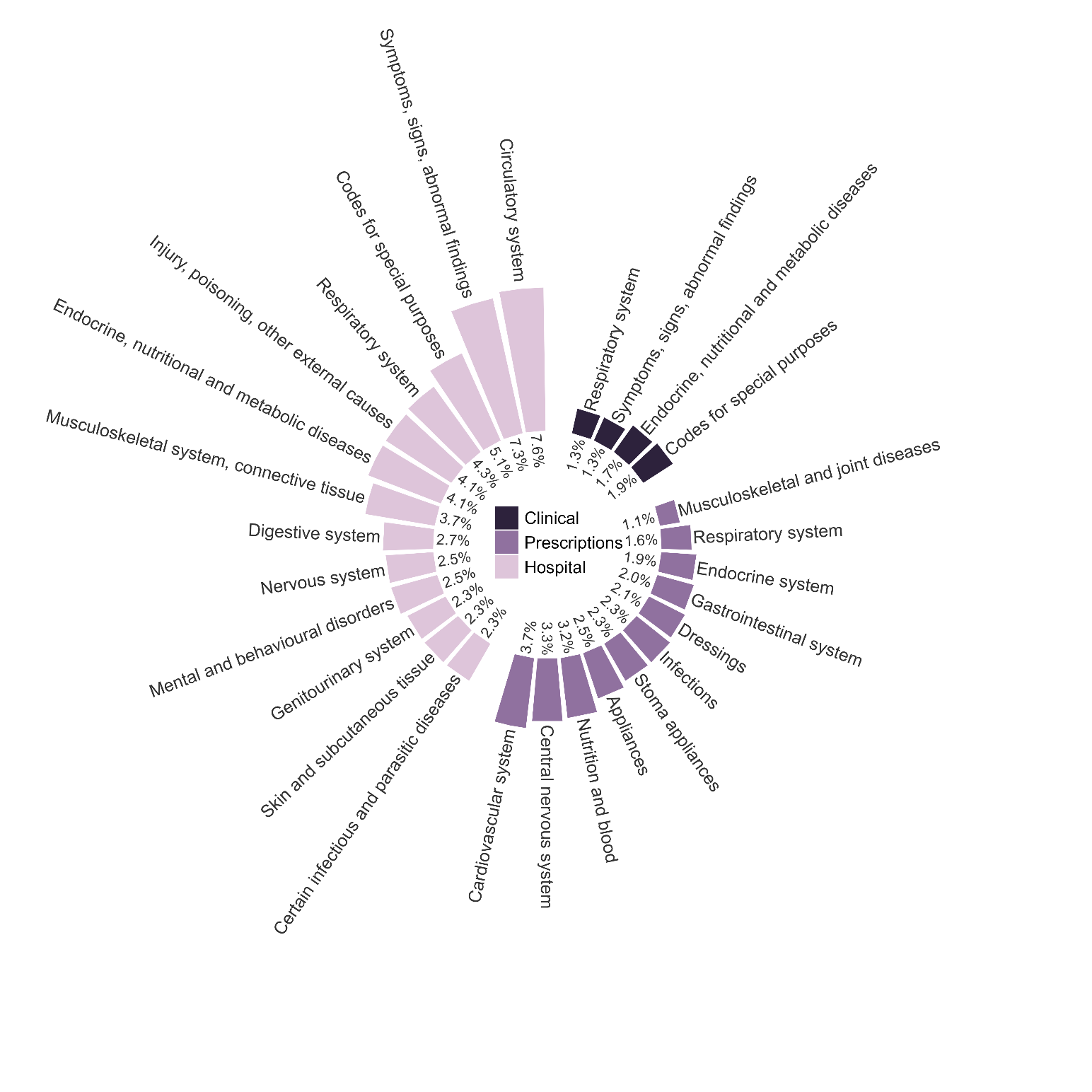

Supplementary Figure 3 Summary of high-level concepts captured in the top 750 ranked high-dimensional propensity score covariates by data dimension for COVID-19 hospitalisations, including triple therapy users

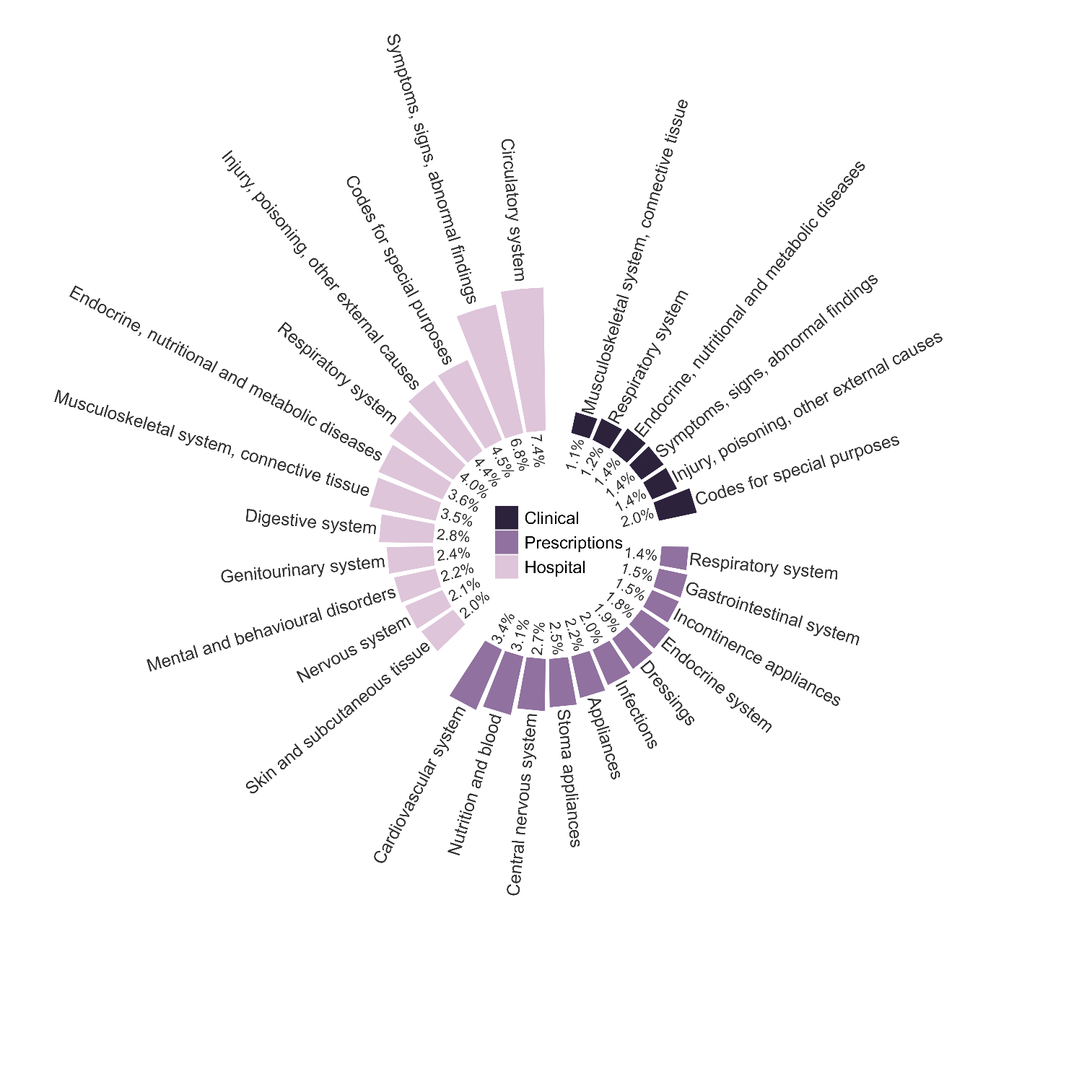

Supplementary Figure 4 Summary of high-level concepts captured in the top 1000 ranked high-dimensional propensity score covariates by data dimension for COVID-19 hospitalisations, including triple therapy users

2.2. Concept plots for outcome COVID-19 hospitalisation, excluding triple therapy users

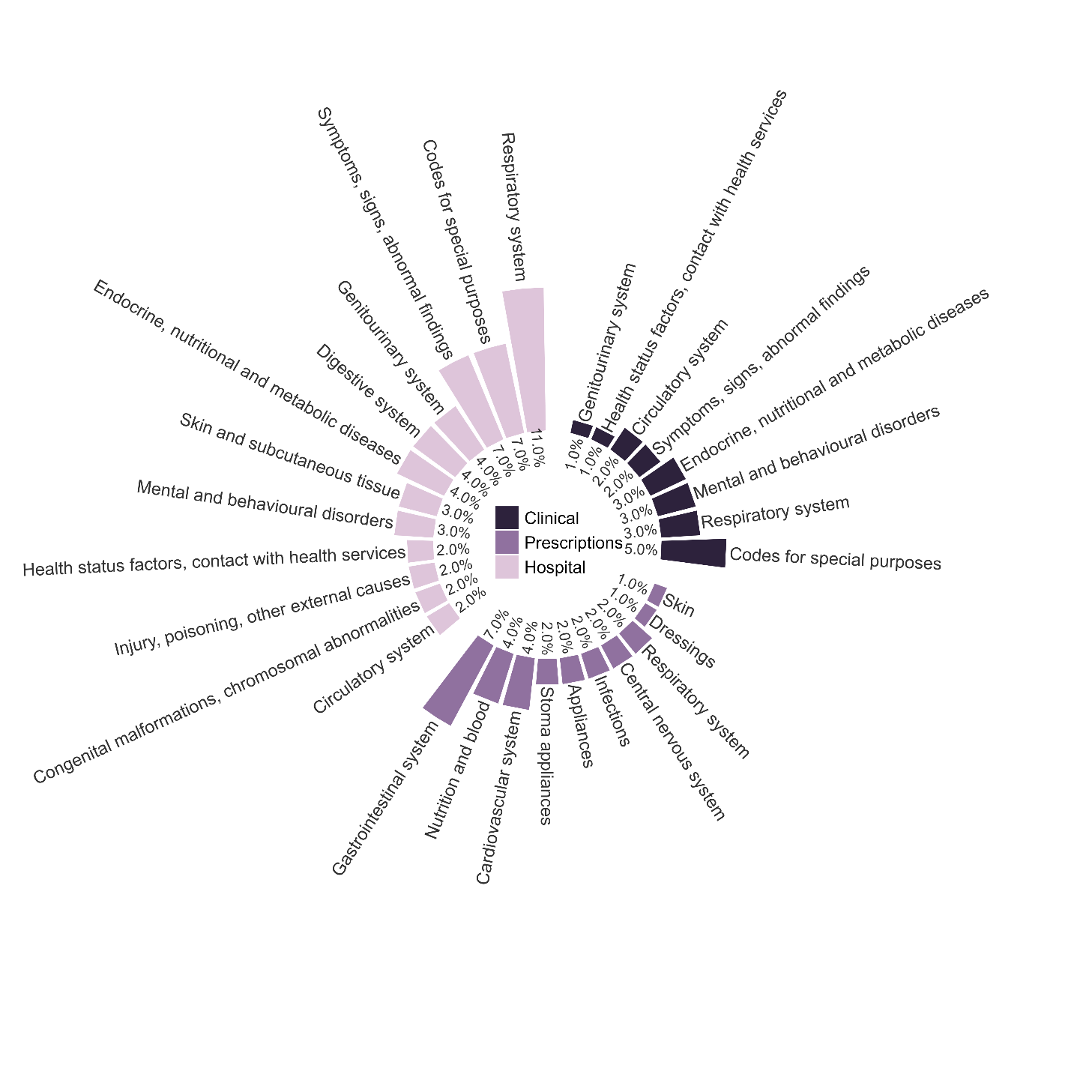

Supplementary Figure 5 Summary of high-level concepts captured in the top 100 ranked high-dimensional propensity score covariates by data dimension for COVID-19 hospitalisations, excluding triple therapy users

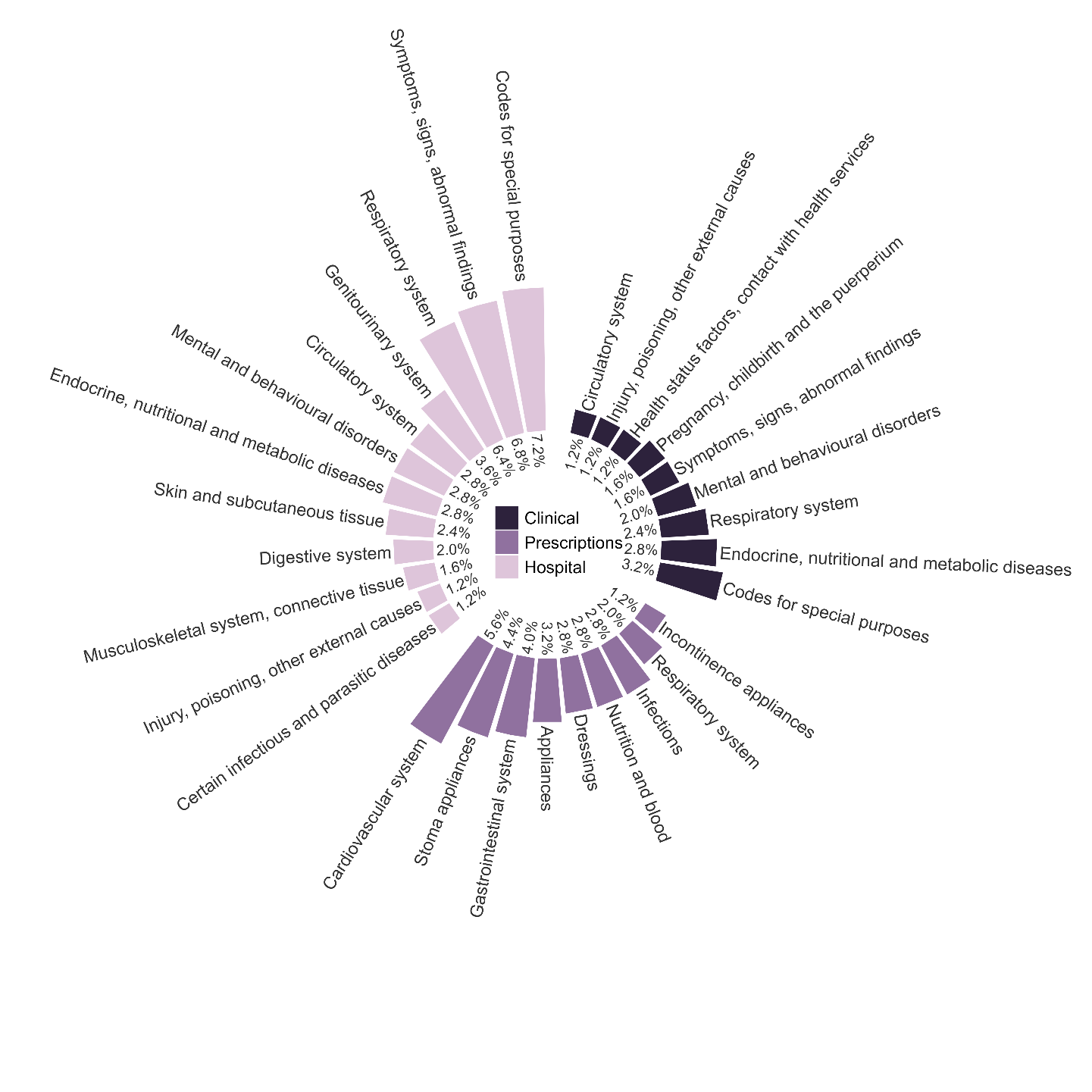

Supplementary Figure 6 Summary of high-level concepts captured in the top 250 ranked high-dimensional propensity score covariates by data dimension for COVID-19 hospitalisations, excluding triple therapy users

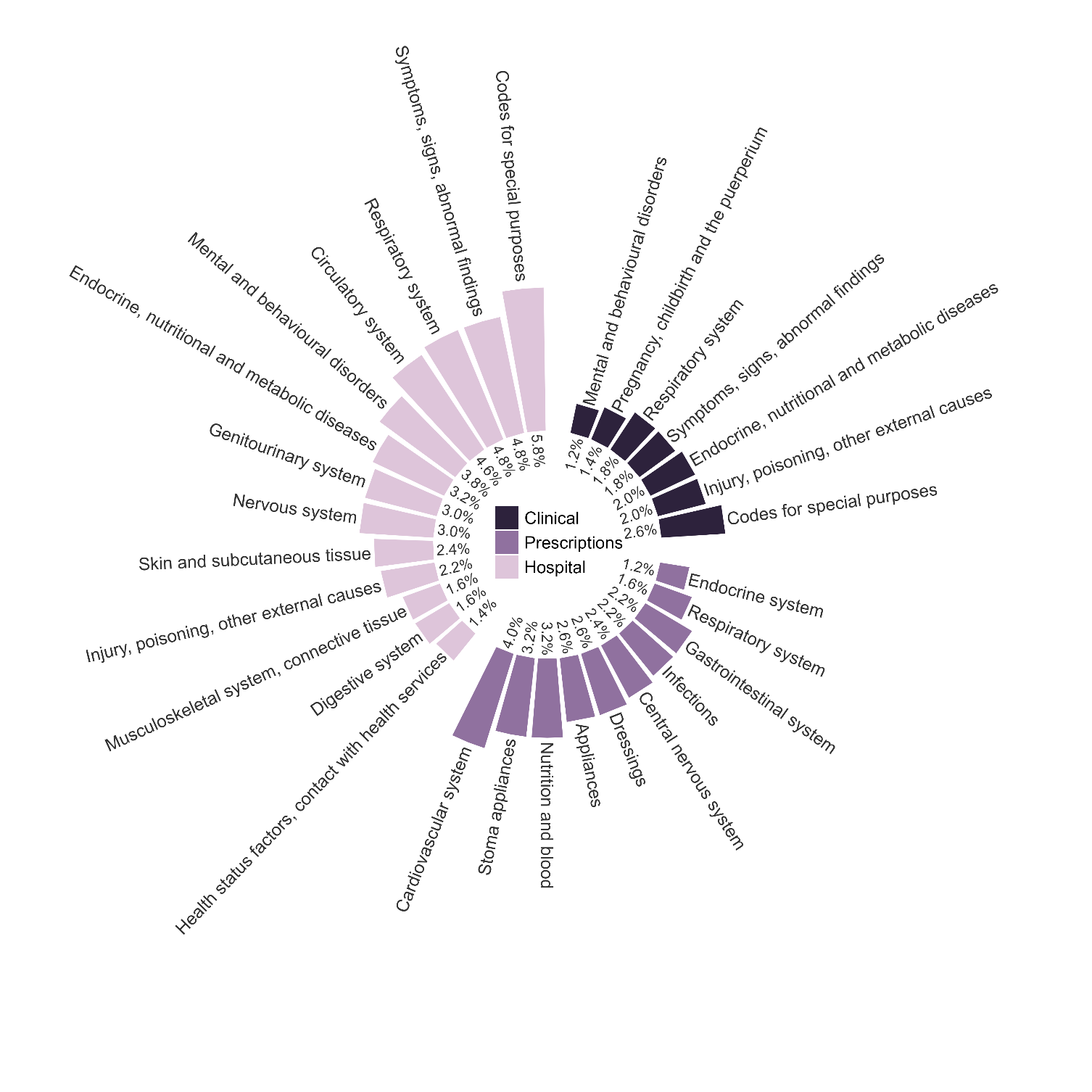

Supplementary Figure 7 Summary of high-level concepts captured in the top 500 ranked high-dimensional propensity score covariates by data dimension for COVID-19 hospitalisations, excluding triple therapy users

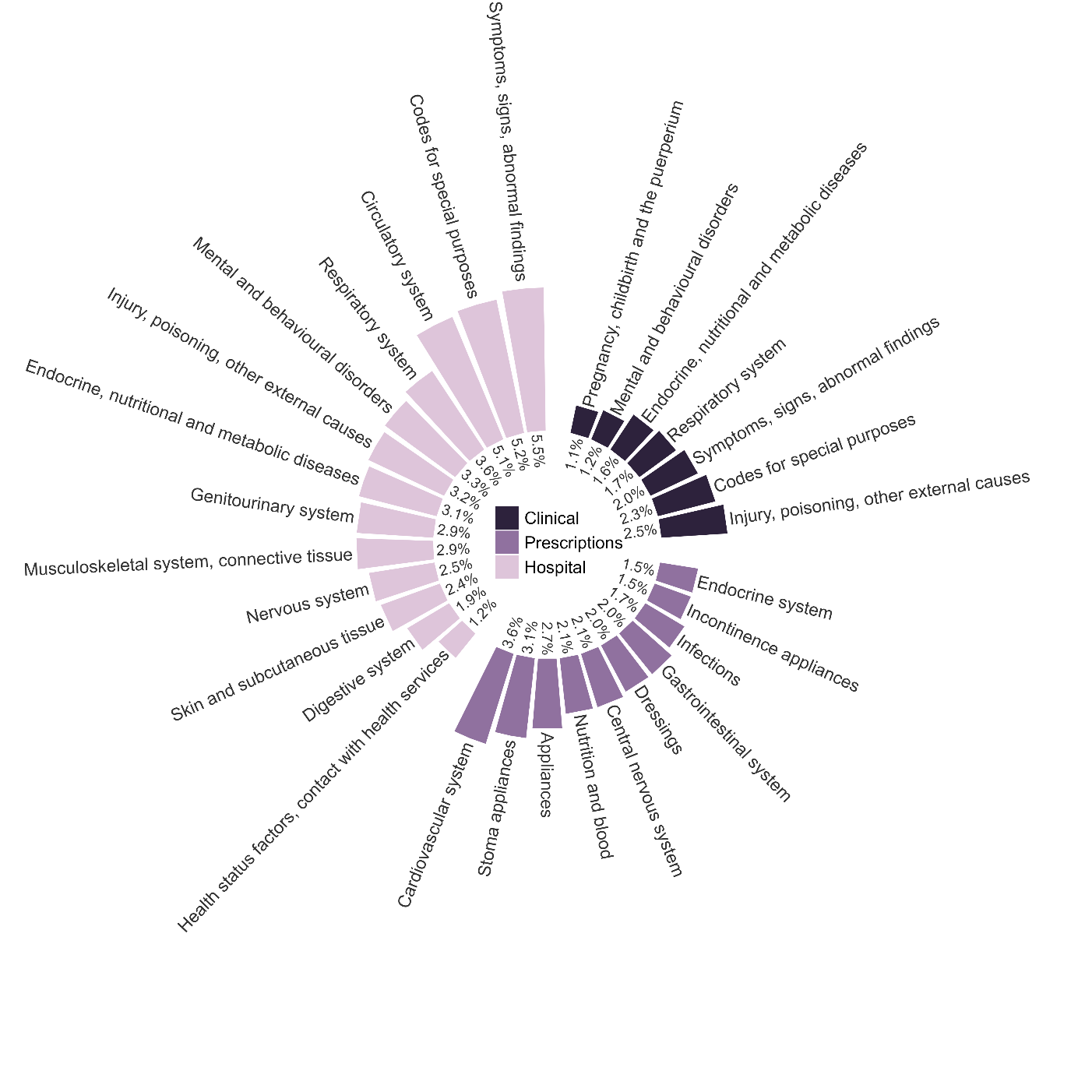

Supplementary Figure 8 Summary of high-level concepts captured in the top 750 ranked high-dimensional propensity score covariates by data dimension for COVID-19 hospitalisations, excluding triple therapy users

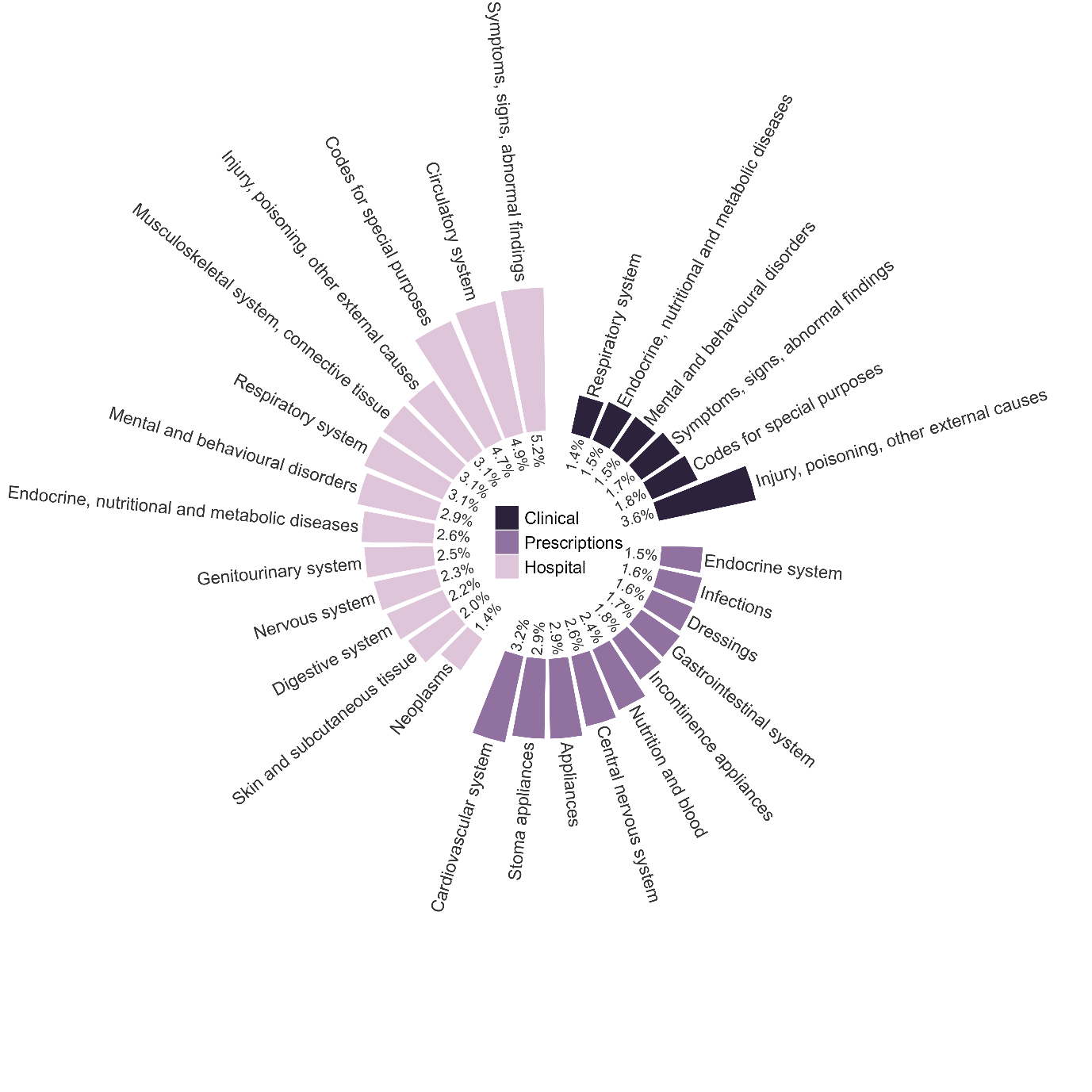

Supplementary Figure 9 Summary of high-level concepts captured in the top 1000 ranked high-dimensional propensity score covariates by data dimension for COVID-19 hospitalisations, excluding triple therapy users

2.3. Concept plots for outcome COVID-19 death, including triple therapy users

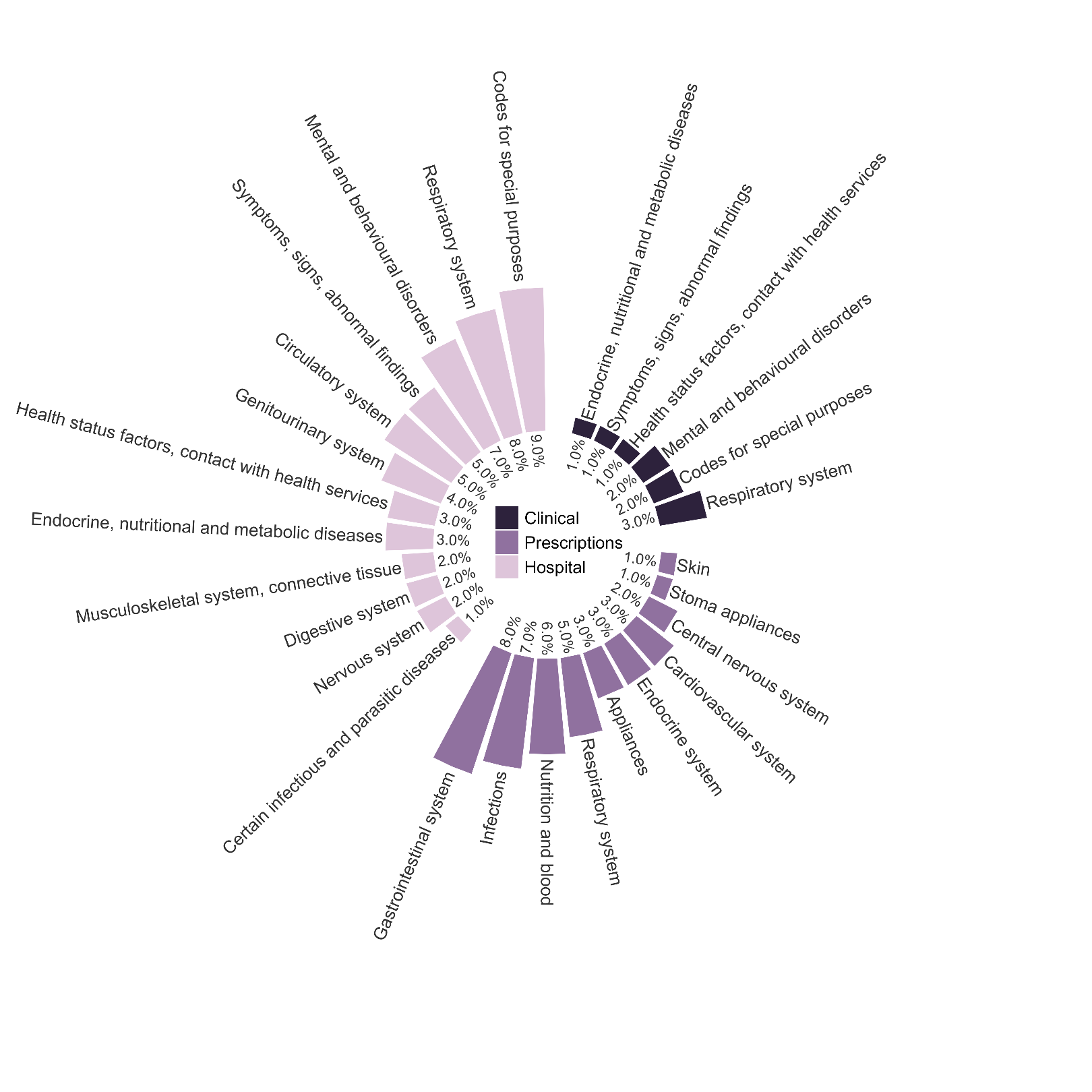

Supplementary Figure 10 Summary of high-level concepts captured in the top 100 ranked high-dimensional propensity score covariates by data dimension for COVID-19 death, including triple therapy users

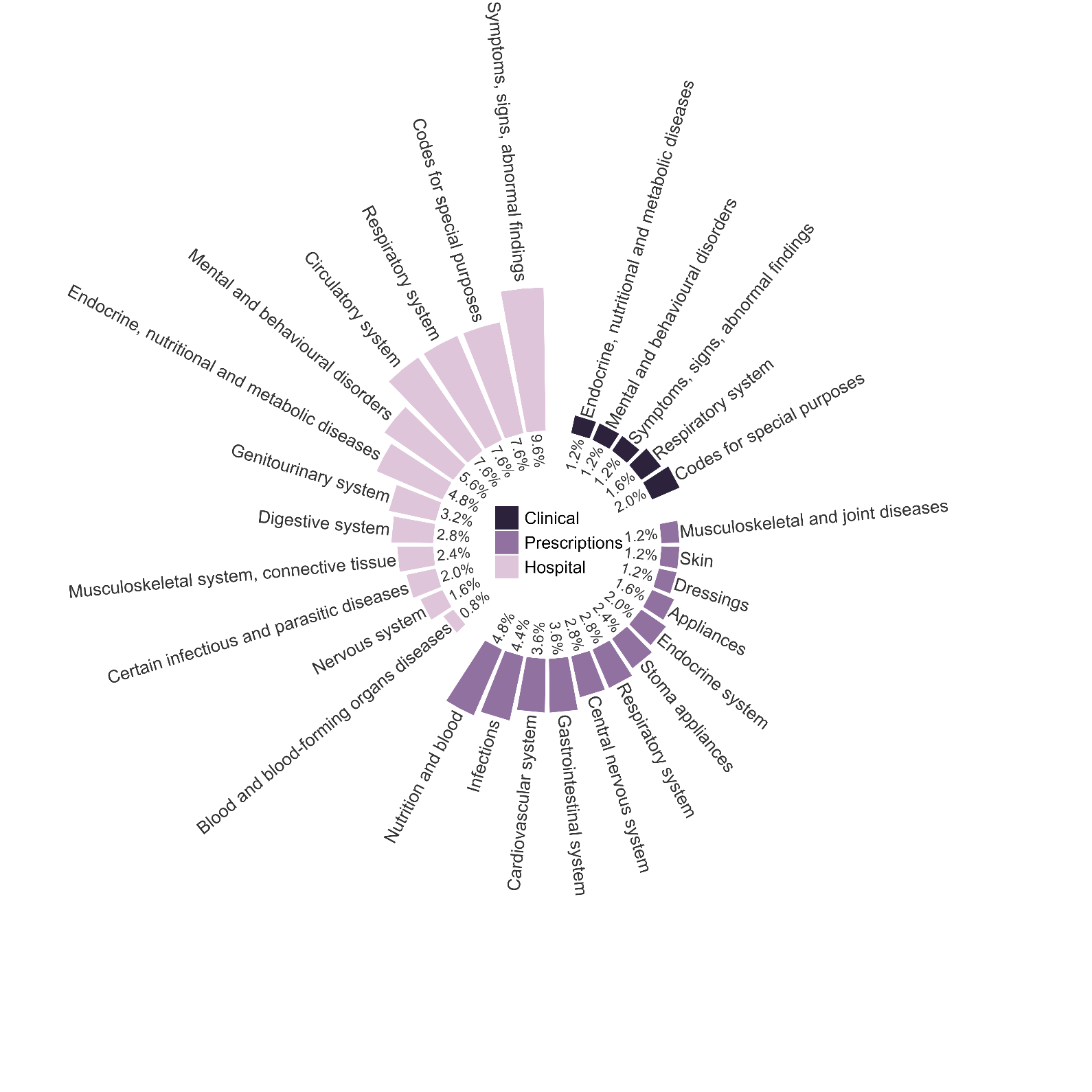

Supplementary Figure 11 Summary of high-level concepts captured in the top 250 ranked high-dimensional propensity score covariates by data dimension for COVID-19 death, including triple therapy users

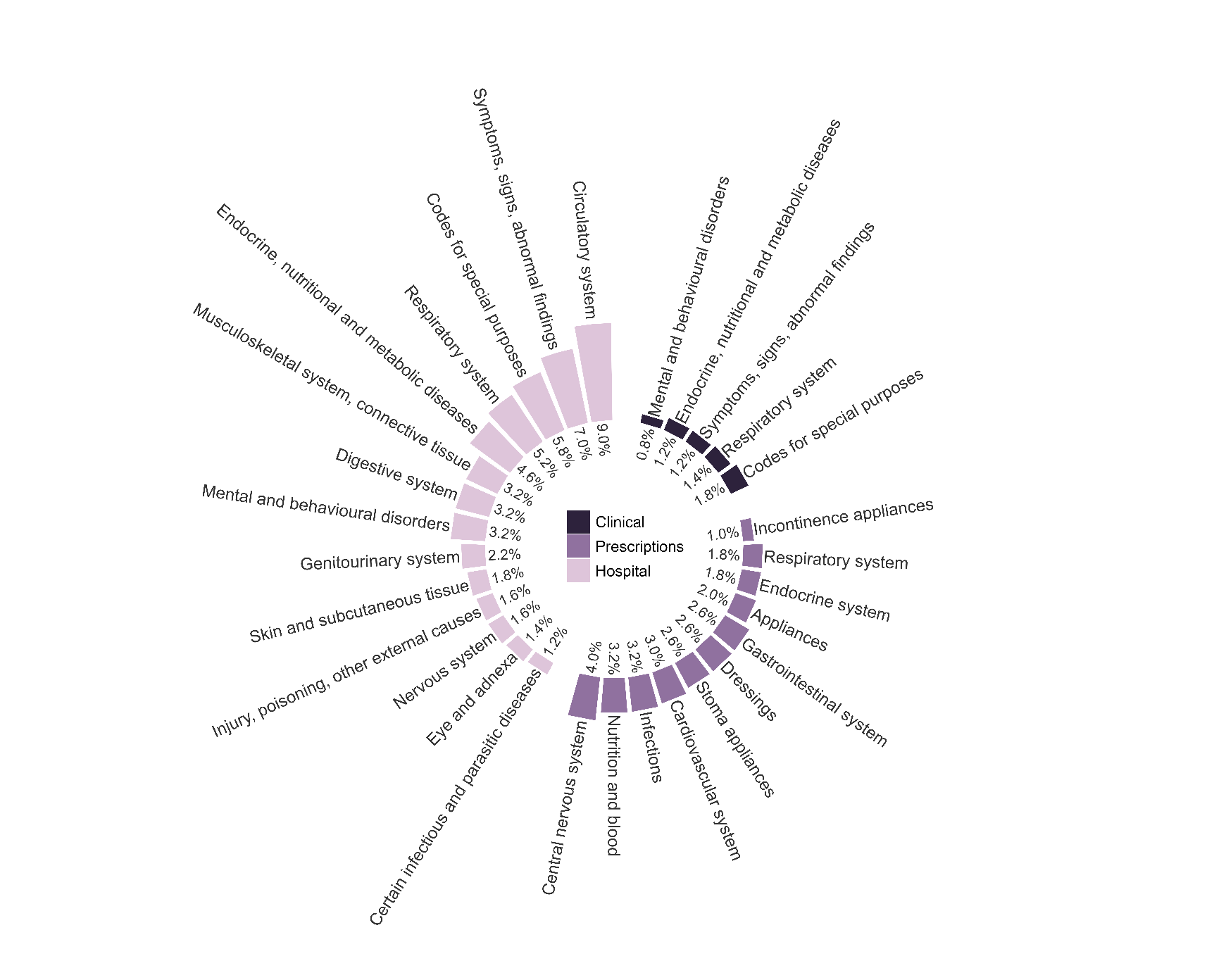

Supplementary Figure 12 Summary of high-level concepts captured in the top 500 ranked high-dimensional propensity score covariates by data dimension for COVID-19 death, including triple therapy users

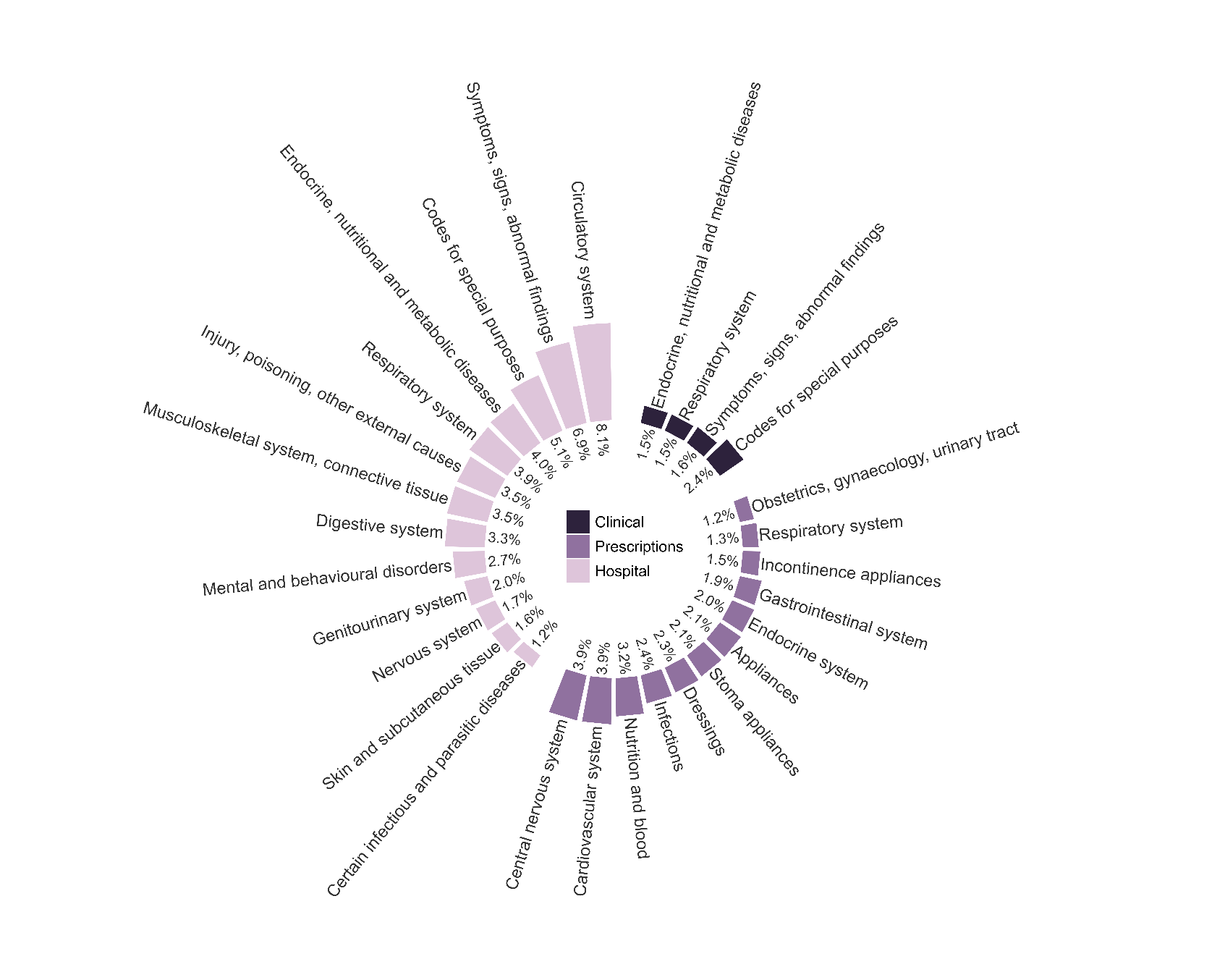

Supplementary Figure 13 Summary of high-level concepts captured in the top 750 ranked high-dimensional propensity score covariates by data dimension for COVID-19 death, including triple therapy users

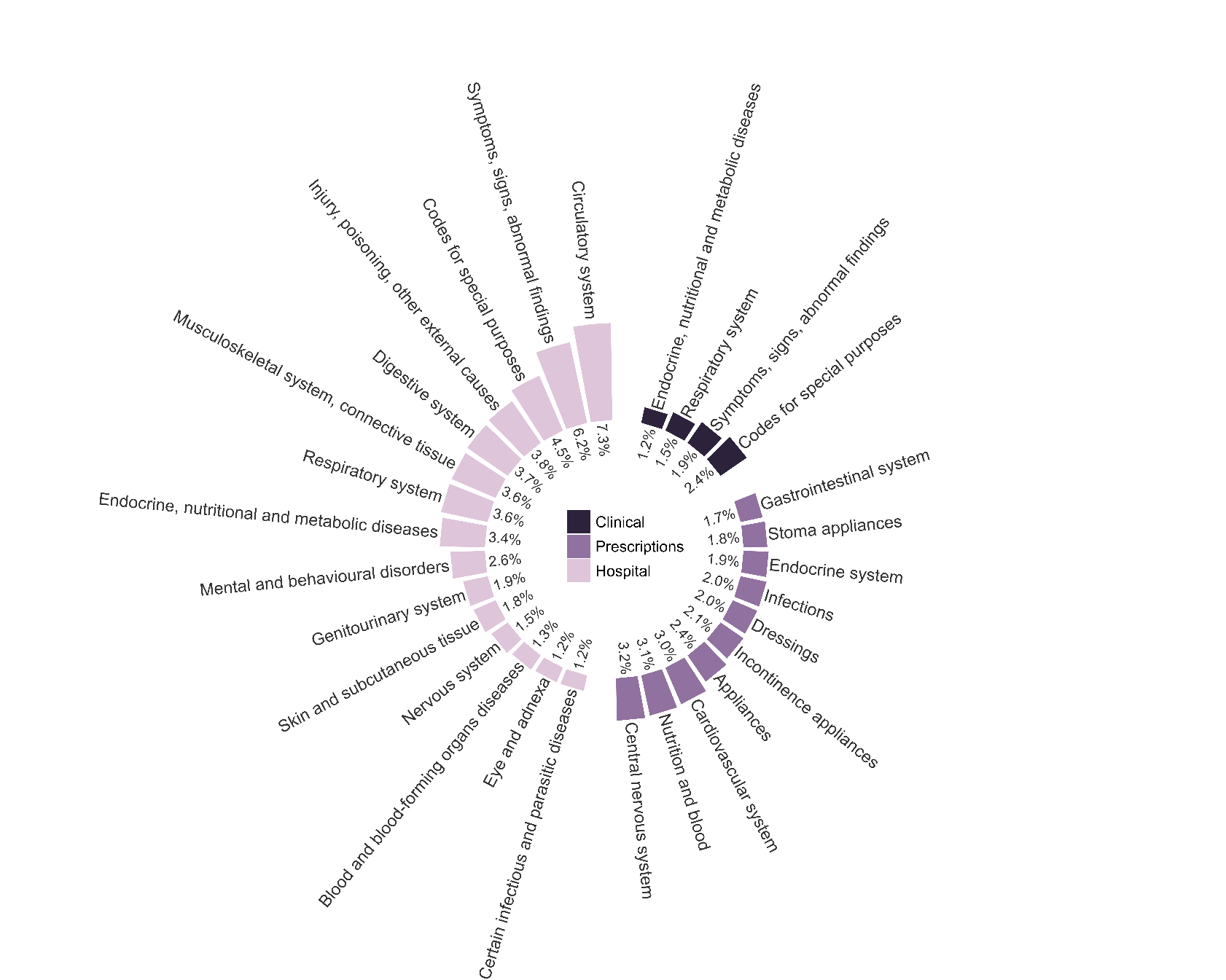

Supplementary Figure 14 Summary of high-level concepts captured in the top 1000 ranked high-dimensional propensity score covariates by data dimension for COVID-19 death, including triple therapy users

2.4. Concept plots for outcome COVID-19 death, excluding triple therapy users

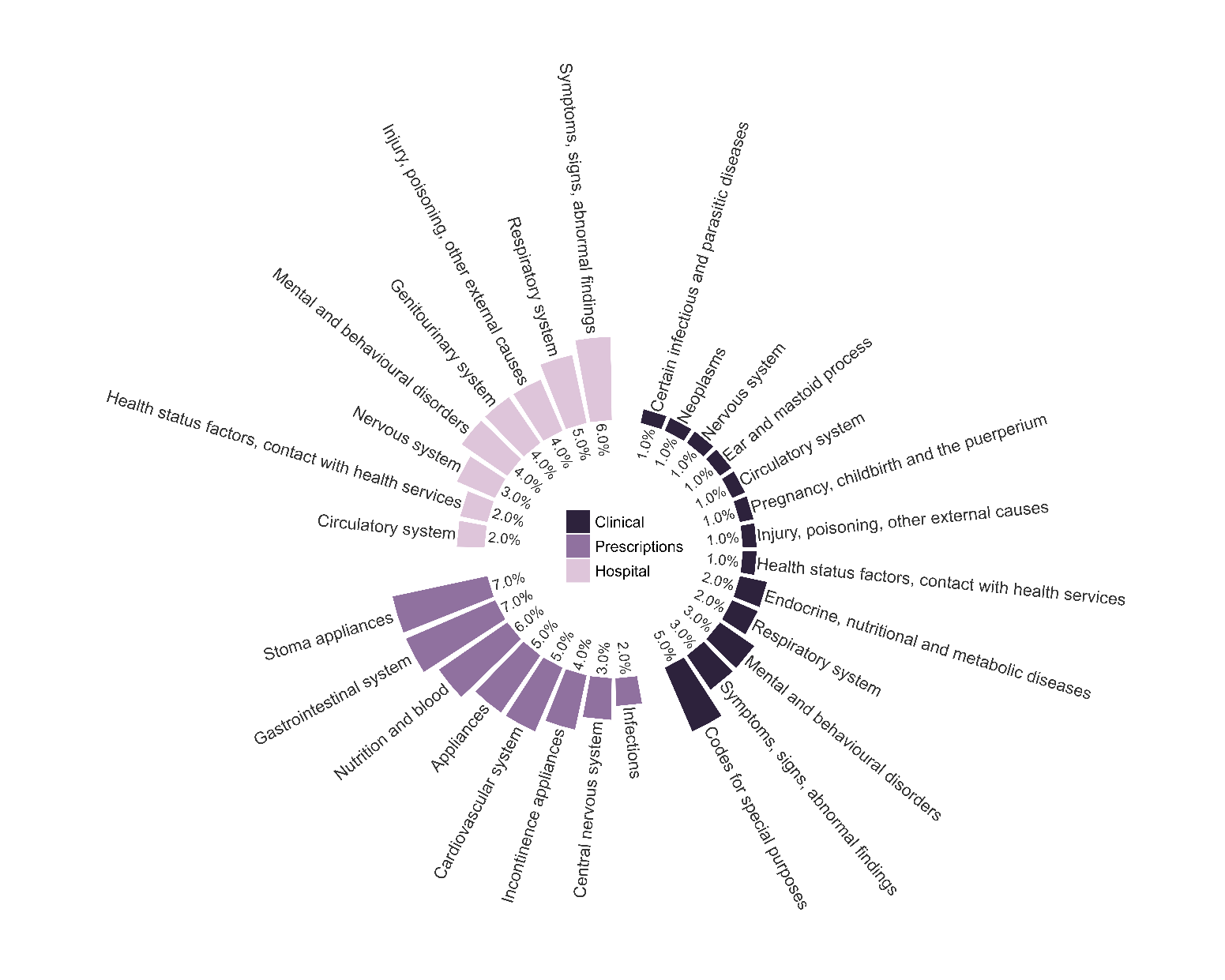

Supplementary Figure 15 Summary of high-level concepts captured in the top 100 ranked high-dimensional propensity score covariates by data dimension for COVID-19 death, excluding triple therapy users

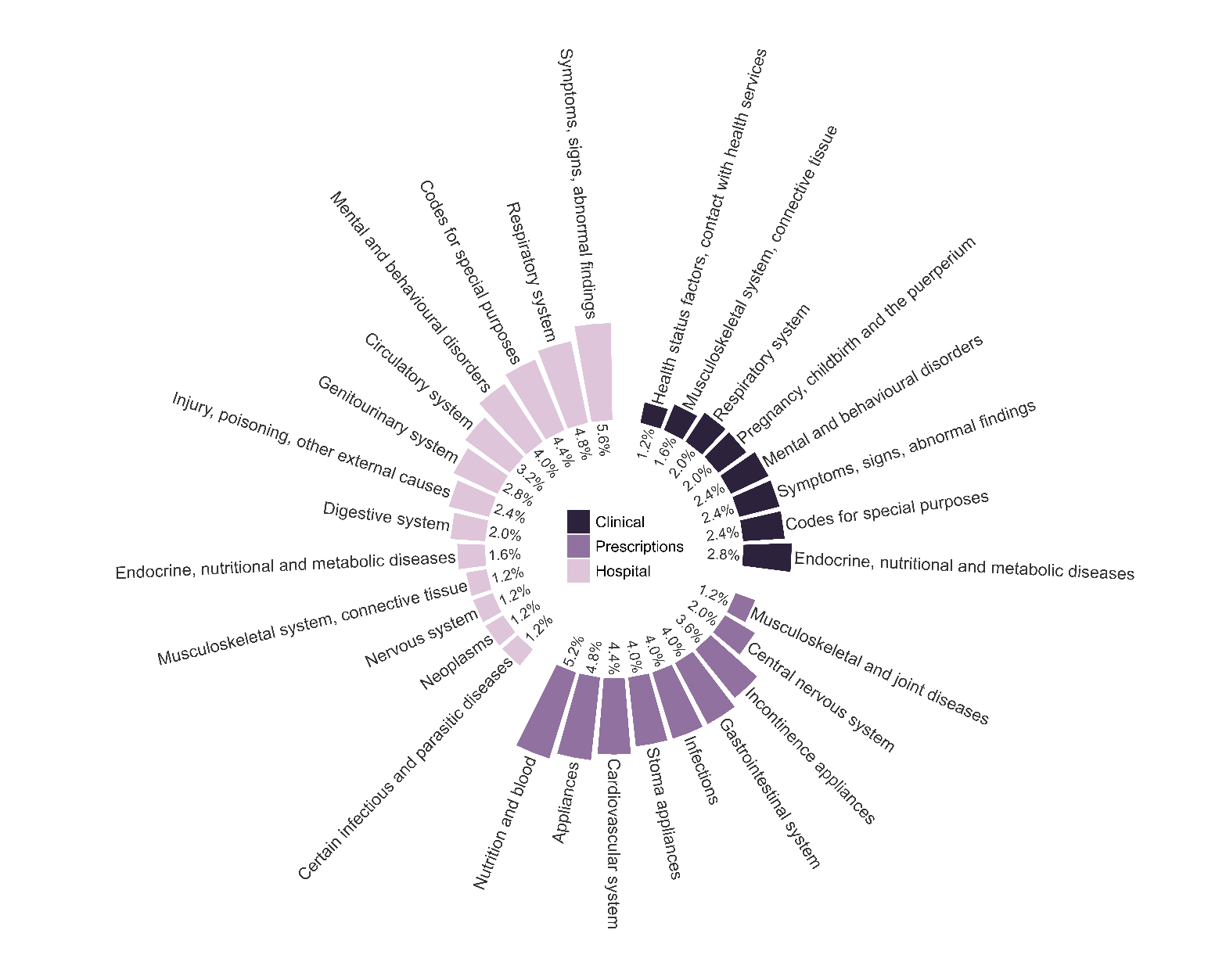

Supplementary Figure 16 Summary of high-level concepts captured in the top 250 ranked high-dimensional propensity score covariates by data dimension for COVID-19 death, excluding triple therapy users

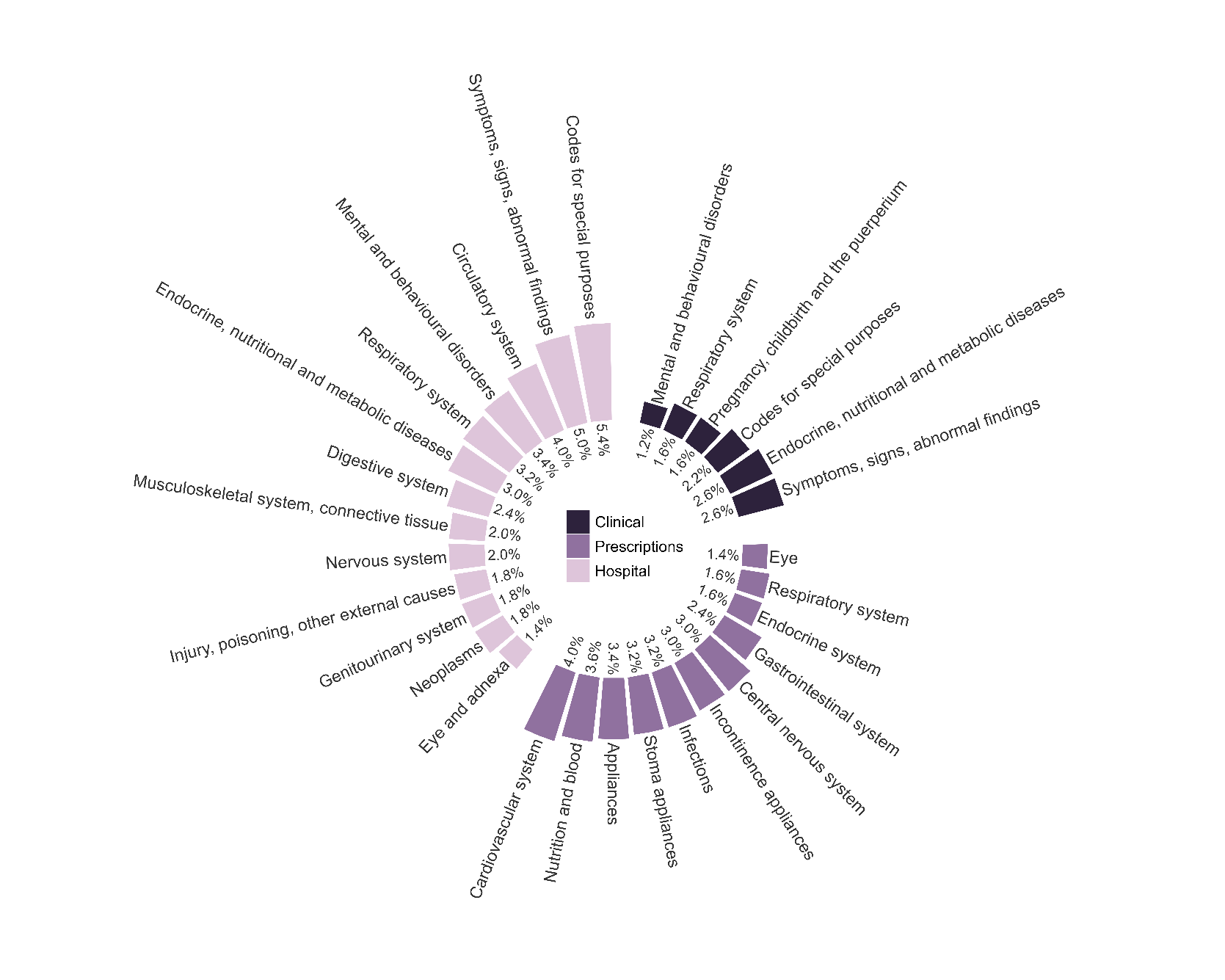

Supplementary Figure 17 Summary of high-level concepts captured in the top 500 ranked high-dimensional propensity score covariates by data dimension for COVID-19 death, including triple therapy users

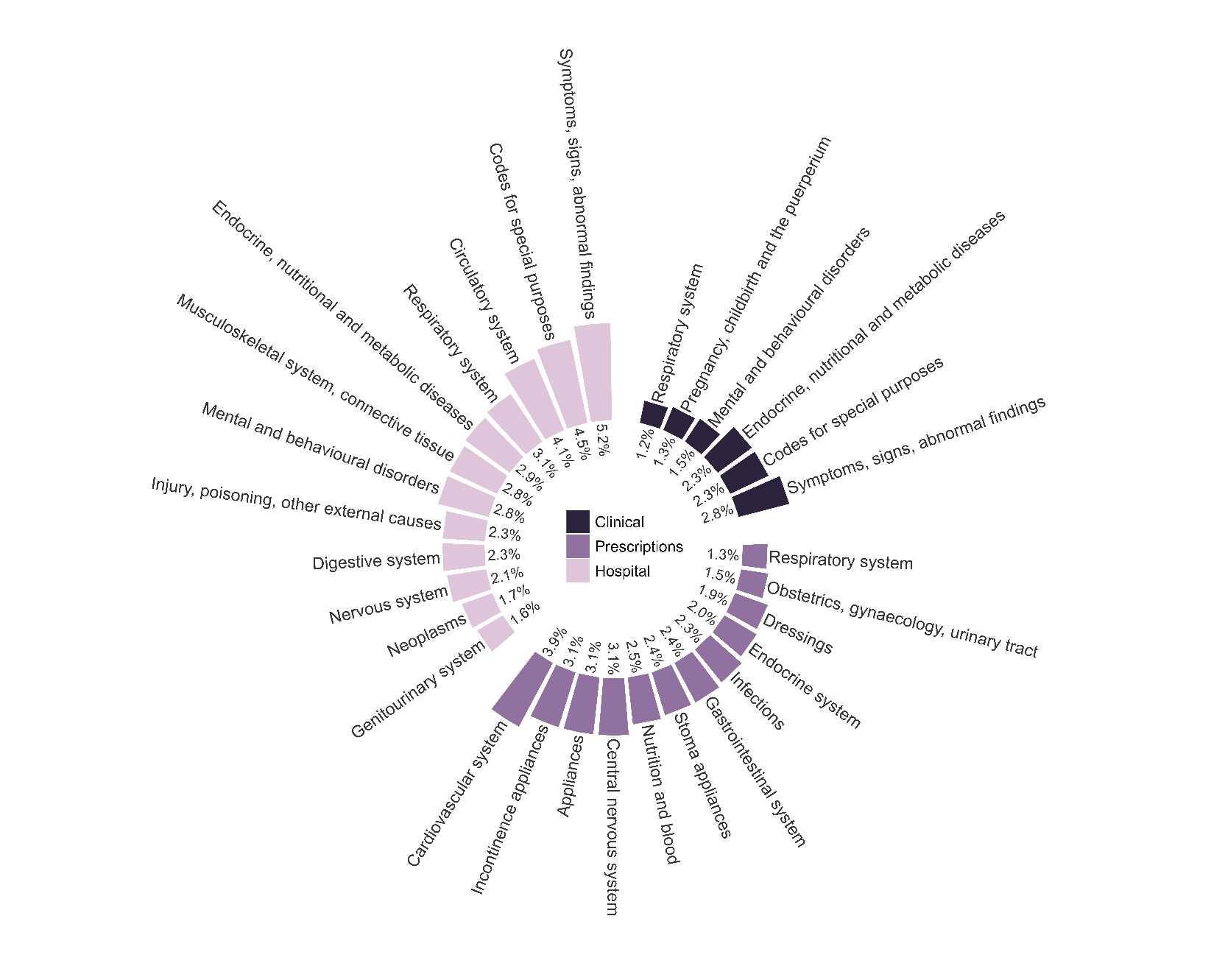

Supplementary Figure 18 Summary of high-level concepts captured in the top 750 ranked high-dimensional propensity score covariates by data dimension for COVID-19 death, excluding triple therapy users

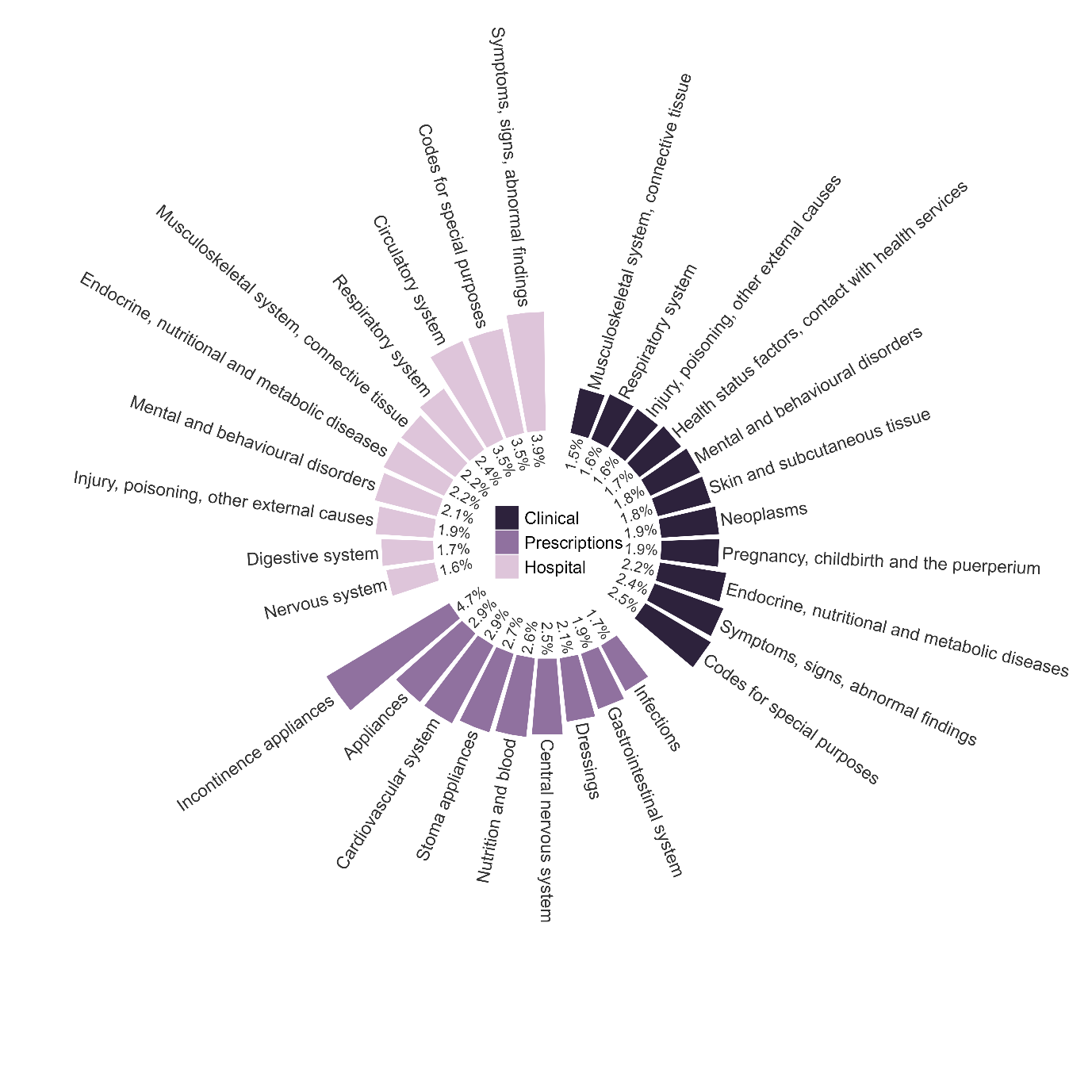

Supplementary Figure 19 Summary of high-level concepts captured in the top 1000 ranked high-dimensional propensity score covariates by data dimension for COVID-19 death, excluding triple therapy users

2.5. Diagnostic plots for HDPS
     2.5.1. COVID-19 hospitalisation, including triple therapy users

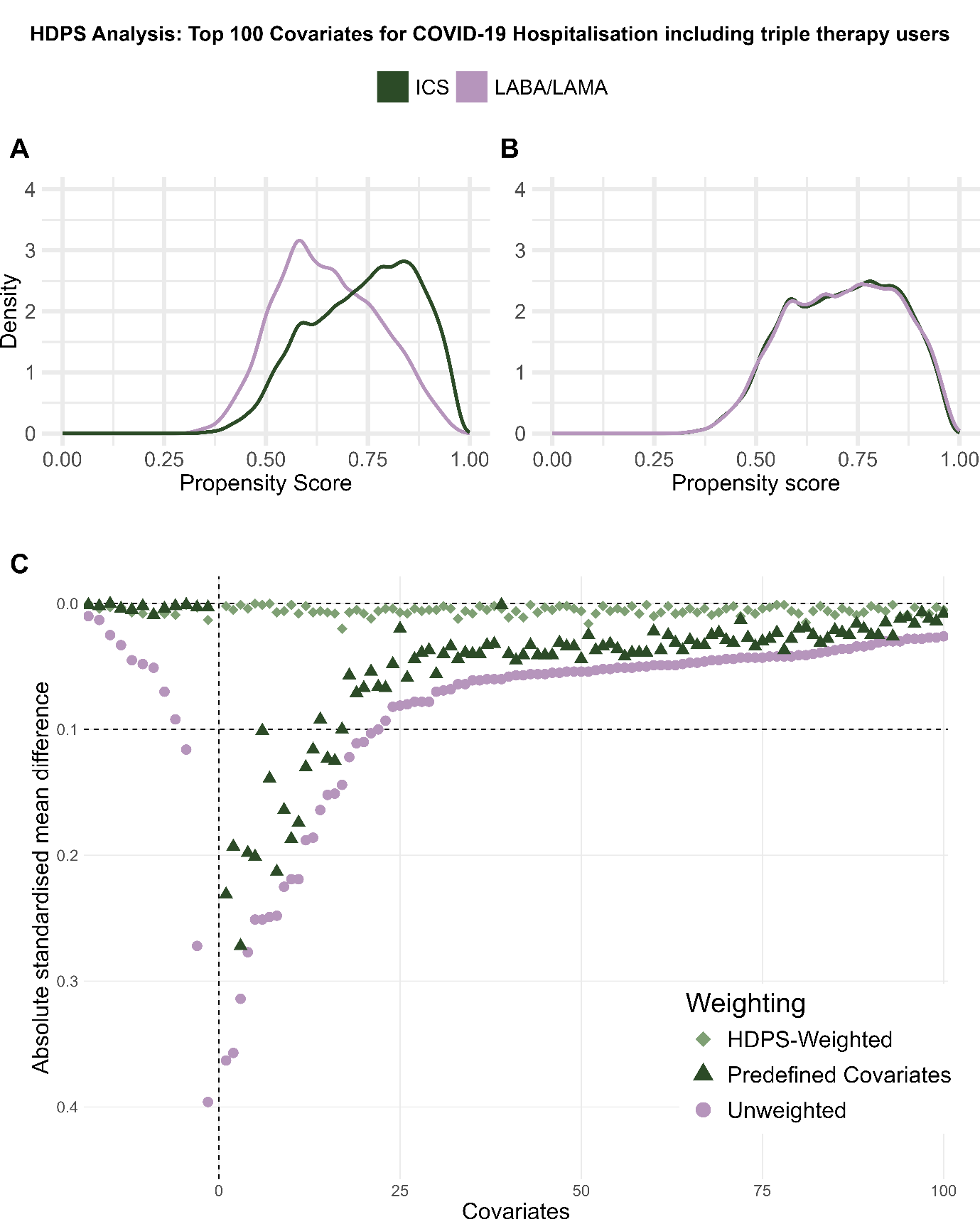

Supplementary Figure 20 Diagnostic plots for high-dimensional propensity score weighted analysis for COVID-19 hospitalisations, including triple therapy users, including the top 100 ranked covariates.

A) high-dimensional propensity score (HDPS) distribution, B) weighted HDPS distribution, C) Comparison of absolute standardised differences in the pre-defined and high-dimensional propensity score covariates between unweighted, predefined and HDPS weighted cohort.

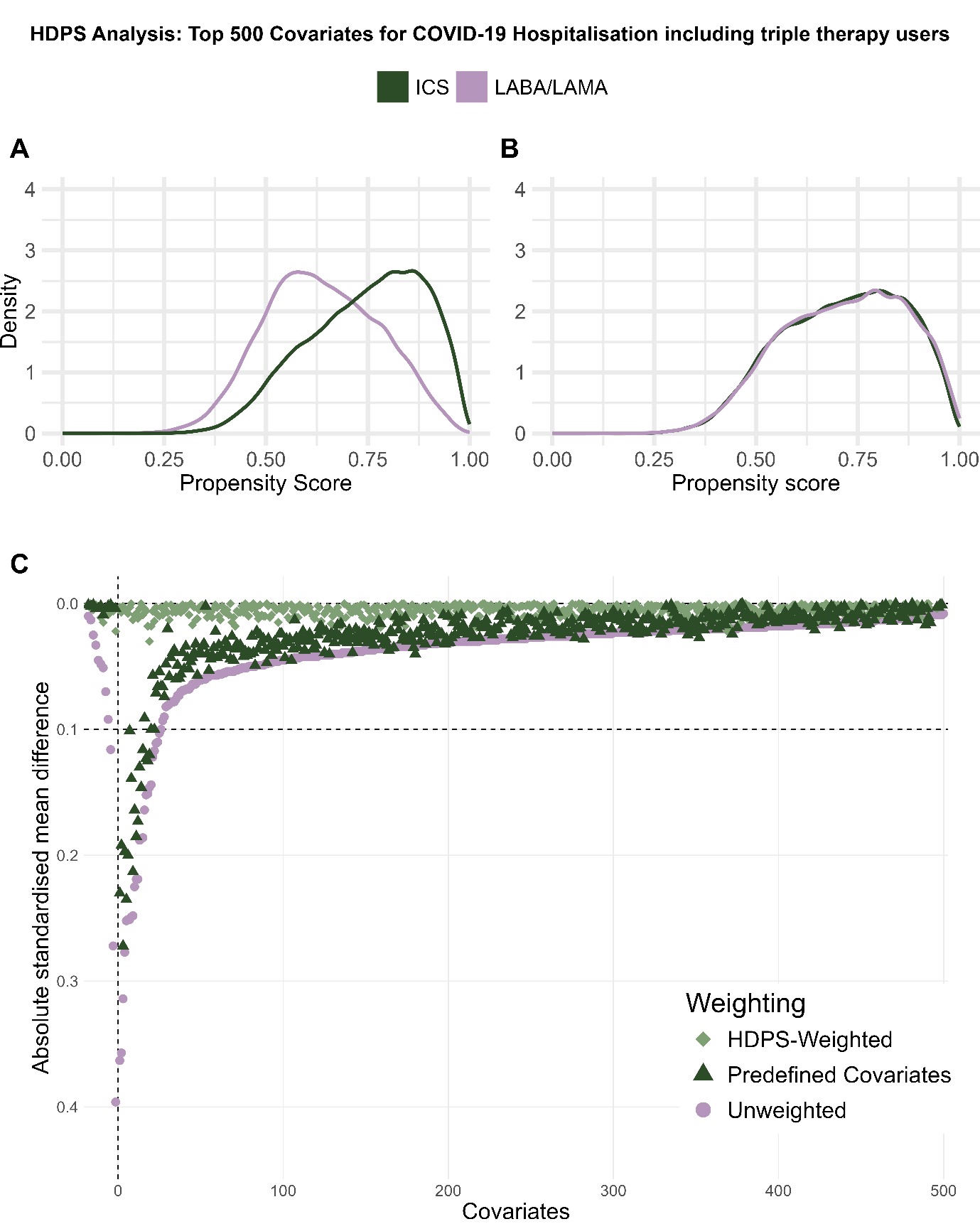

Supplementary Figure 21 Diagnostic plots for high-dimensional propensity score weighted analysis for COVID-19 hospitalisations, including triple therapy users, including the top 500 ranked covariates.

A) high-dimensional propensity score (HDPS) distribution, B) weighted HDPS distribution, C) Comparison of absolute standardised differences in the pre-defined and high-dimensional propensity score covariates between unweighted, predefined and HDPS weighted cohort.

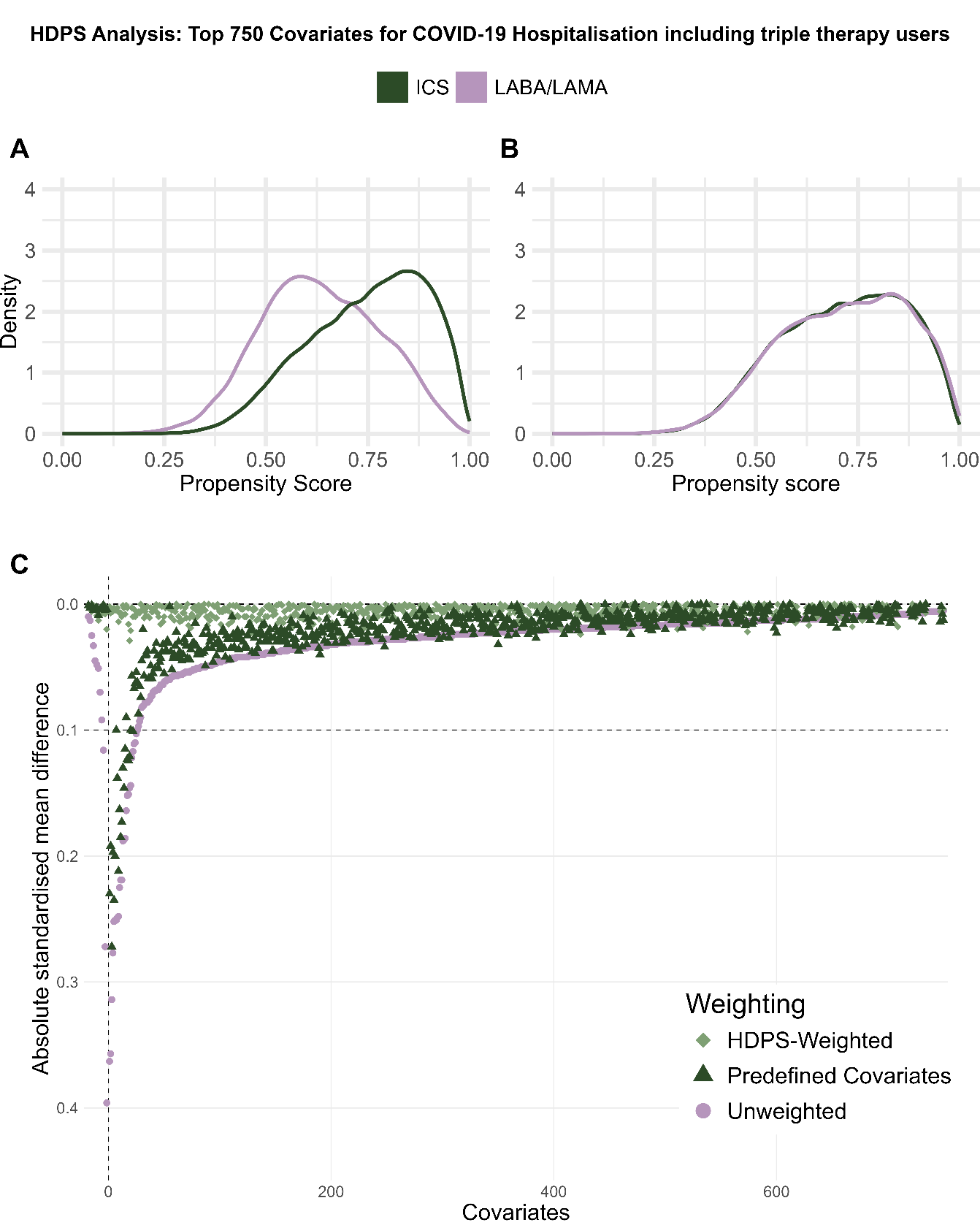

Supplementary Figure 22 Diagnostic plots for high-dimensional propensity score weighted analysis for COVID-19 hospitalisations, including triple therapy users, including the top 750 ranked covariates.

A) high-dimensional propensity score (HDPS) distribution, B) weighted HDPS distribution, C) Comparison of absolute standardised differences in the pre-defined and high-dimensional propensity score covariates between unweighted, predefined and HDPS weighted cohort.

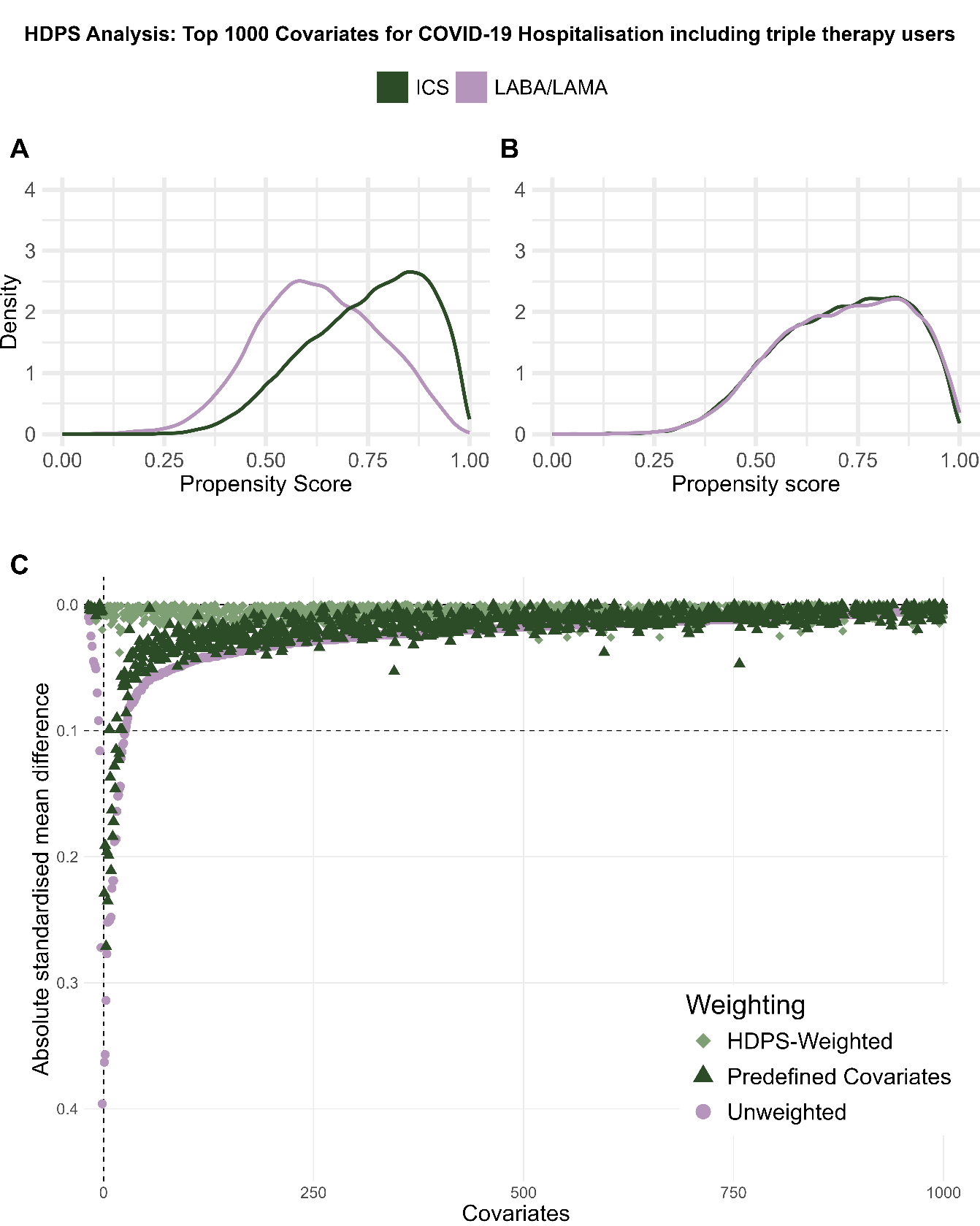

Supplementary Figure 23 Diagnostic plots for high-dimensional propensity score weighted analysis for COVID-19 hospitalisations, including triple therapy users, including the top 1000 ranked covariates.

A) high-dimensional propensity score (HDPS) distribution, B) weighted HDPS distribution, C) Comparison of absolute standardised differences in the pre-defined and high-dimensional propensity score covariates between unweighted, predefined and HDPS weighted cohort.

2.5.2. COVID-19 hospitalisation, excluding triple therapy users

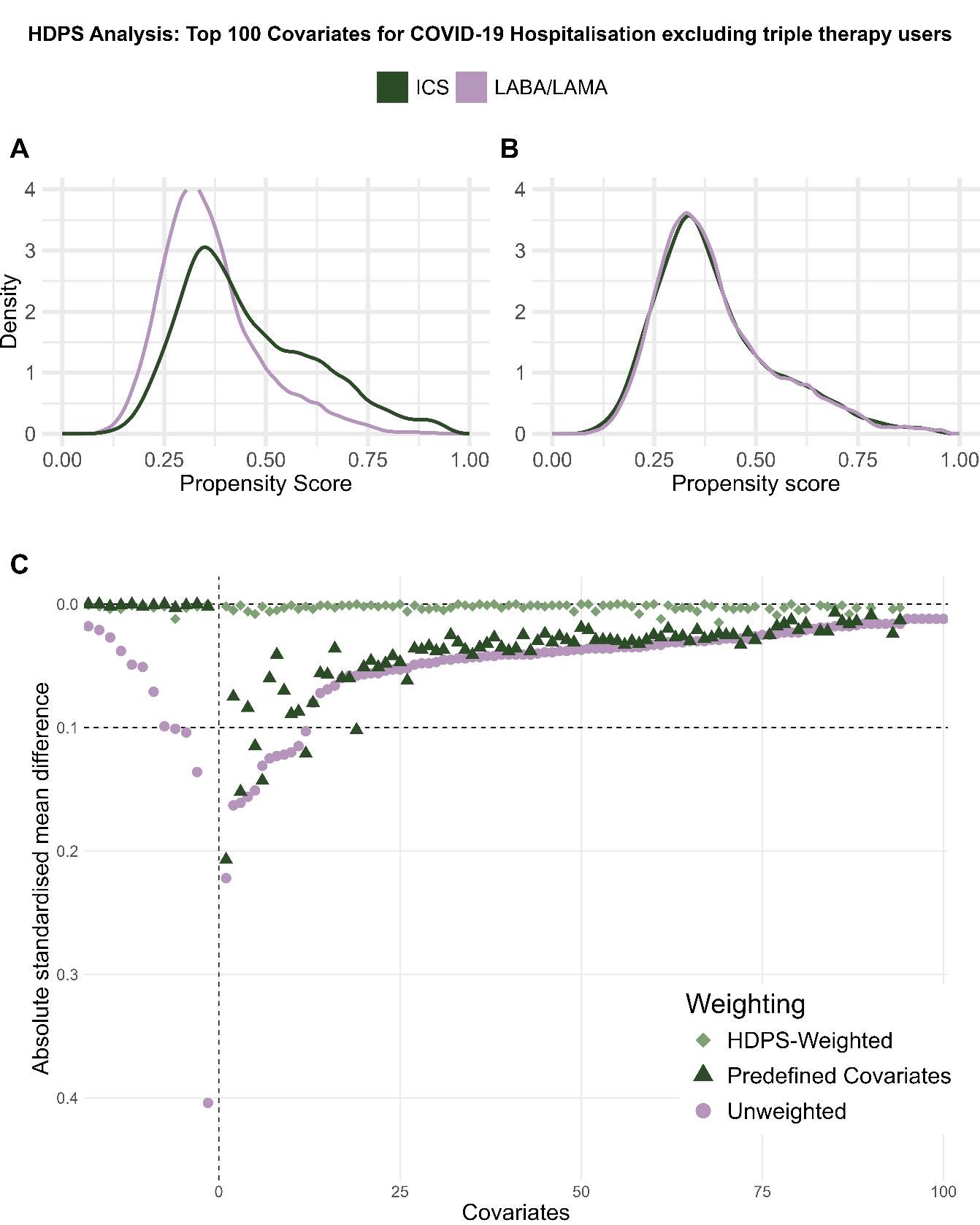

Supplementary Figure 24 Diagnostic plots for high-dimensional propensity score weighted analysis for COVID-19 hospitalisations, excluding triple therapy users, including the top 100 ranked covariates.

A) high-dimensional propensity score (HDPS) distribution, B) weighted HDPS distribution, C) Comparison of absolute standardised differences in the pre-defined and high-dimensional propensity score covariates between unweighted, predefined and HDPS weighted cohort.

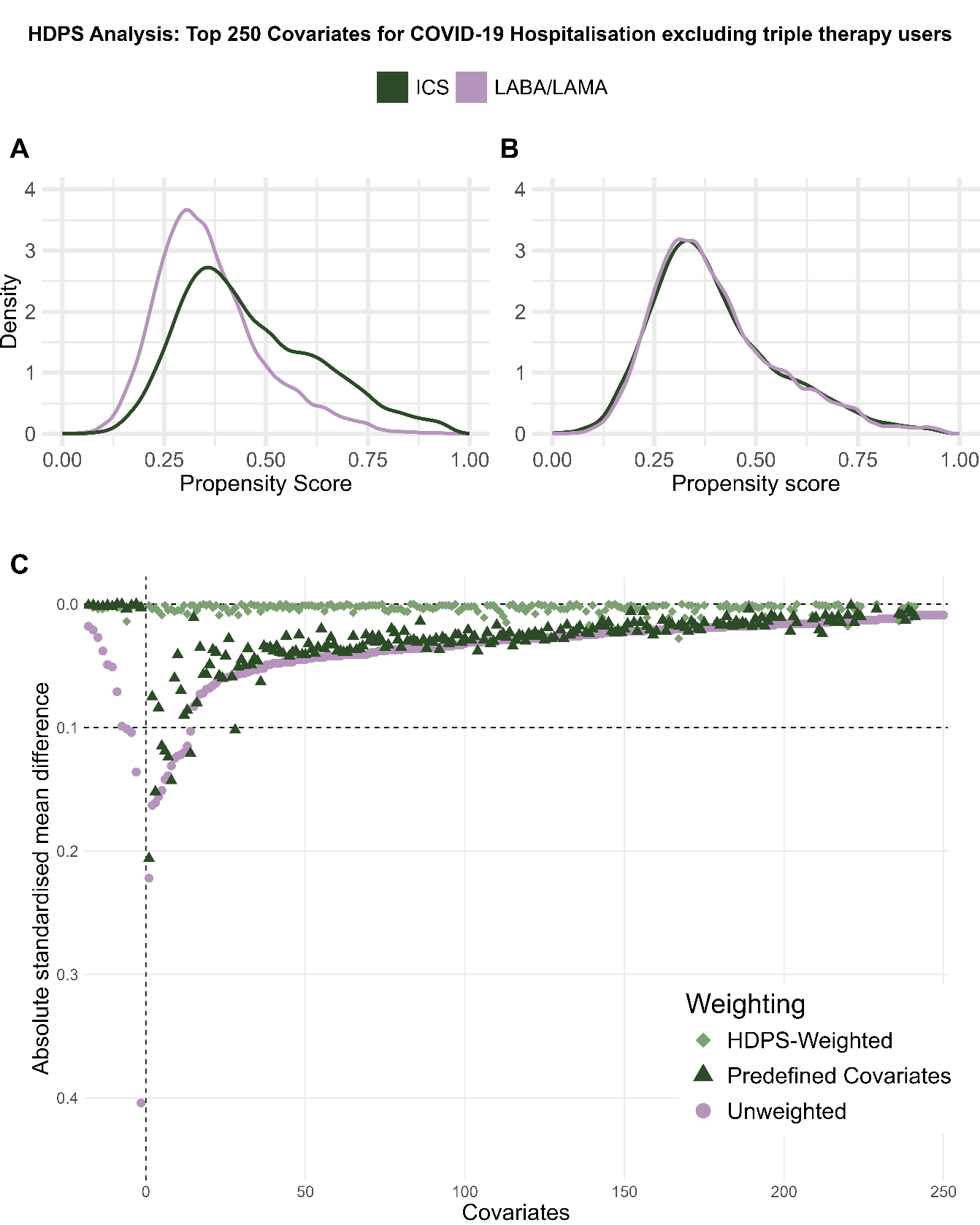

Supplementary Figure 25 Diagnostic plots for high-dimensional propensity score weighted analysis for COVID-19 hospitalisations, excluding triple therapy users, including the top 250 ranked covariates.

A) high-dimensional propensity score (HDPS) distribution, B) weighted HDPS distribution, C) Comparison of absolute standardised differences in the pre-defined and high-dimensional propensity score covariates between unweighted, predefined and HDPS weighted cohort.

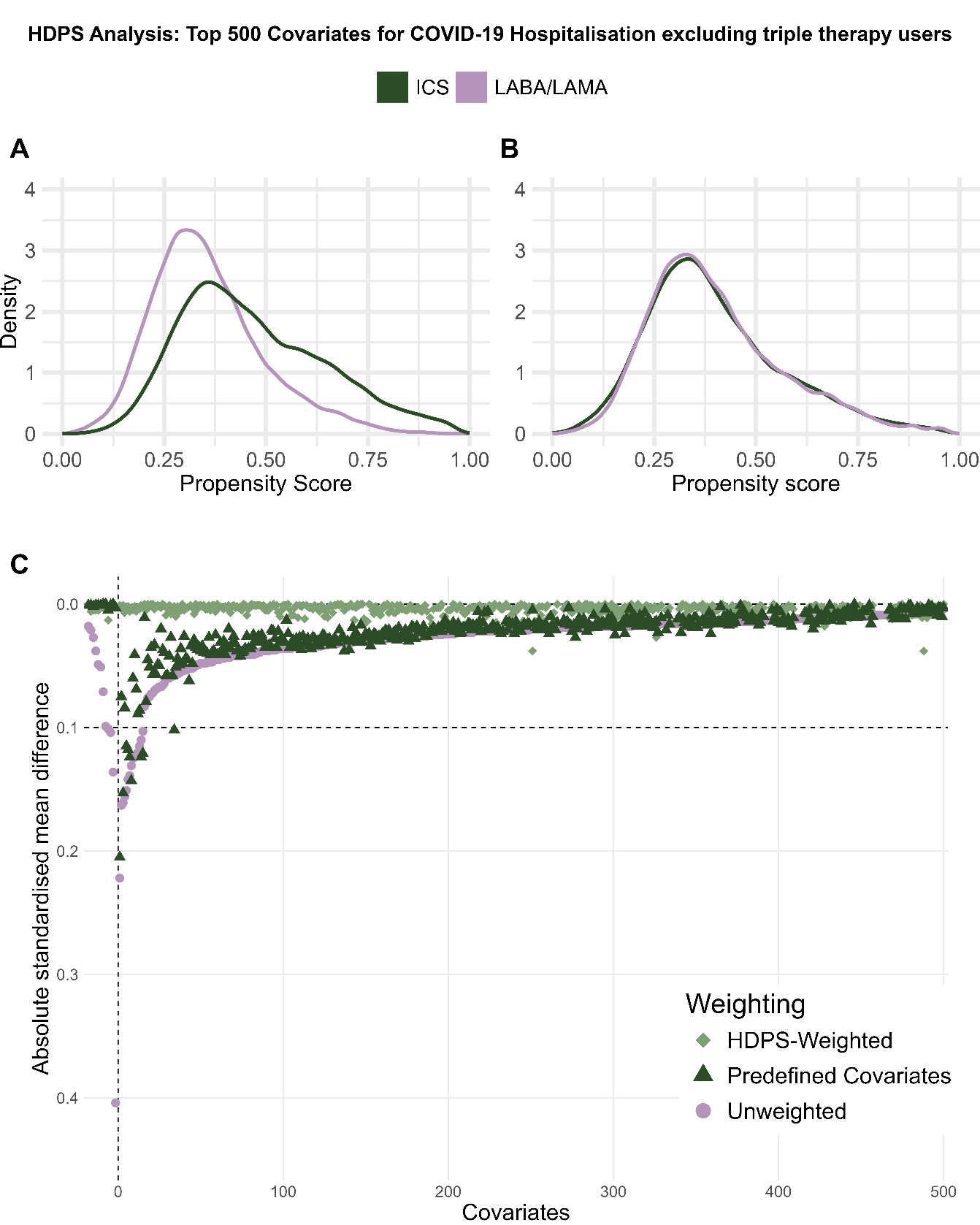

Supplementary Figure 26 Diagnostic plots for high-dimensional propensity score weighted analysis for COVID-19 hospitalisations, excluding triple therapy users, including the top 500 ranked covariates.

A) high-dimensional propensity score (HDPS) distribution, B) weighted HDPS distribution, C) Comparison of absolute standardised differences in the pre-defined and high-dimensional propensity score covariates between unweighted, predefined and HDPS weighted cohort.

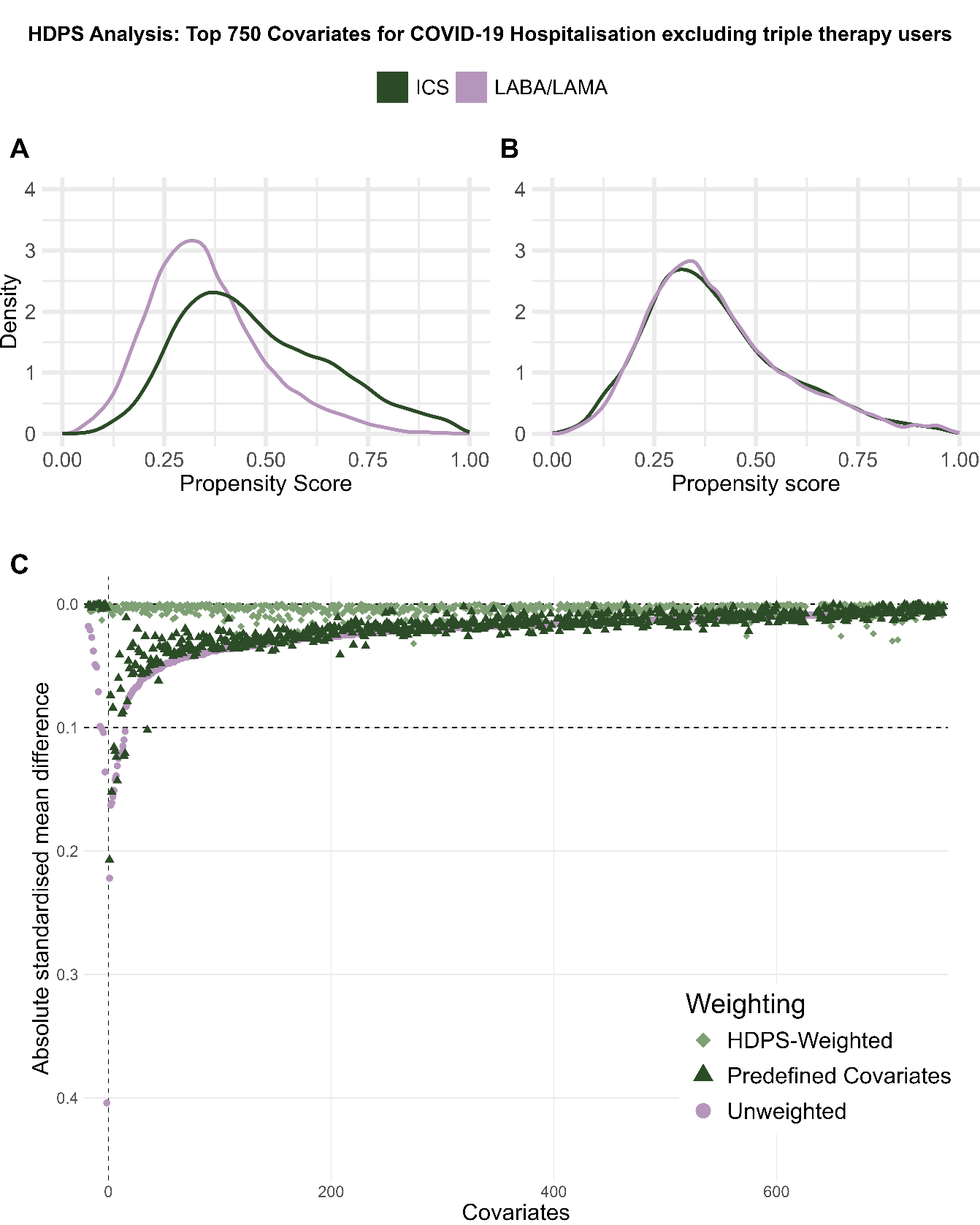

Supplementary Figure 27 Diagnostic plots for high-dimensional propensity score weighted analysis for COVID-19 hospitalisations, excluding triple therapy users, including the top 750 ranked covariates.

A) high-dimensional propensity score (HDPS) distribution, B) weighted HDPS distribution, C) Comparison of absolute standardised differences in the pre-defined and high-dimensional propensity score covariates between unweighted, predefined and HDPS weighted cohort.

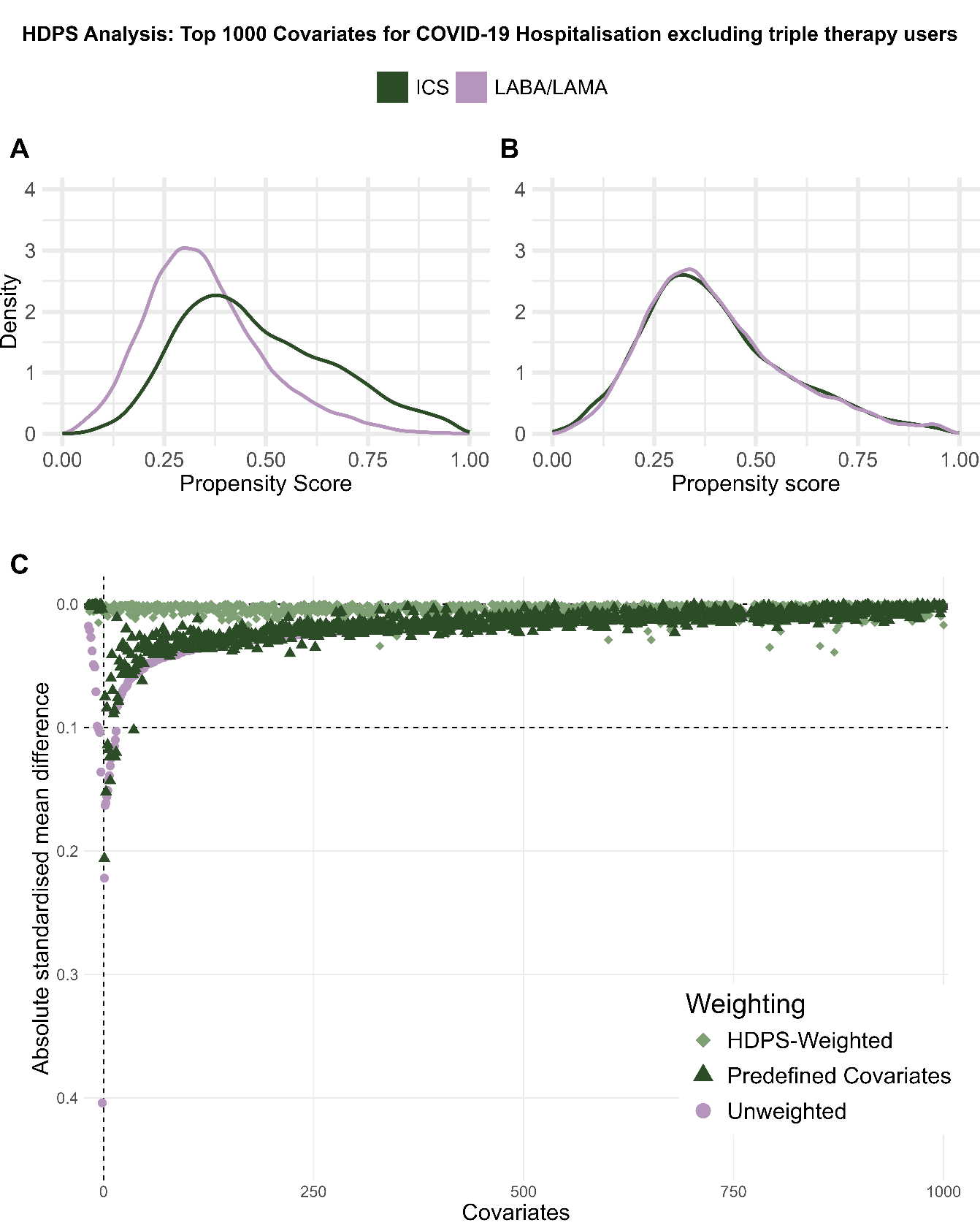

Supplementary Figure 28 Diagnostic plots for high-dimensional propensity score weighted analysis for COVID-19 hospitalisations, excluding triple therapy users, including the top 1000 ranked covariates.

A) high-dimensional propensity score (HDPS) distribution, B) weighted HDPS distribution, C) Comparison of absolute standardised differences in the pre-defined and high-dimensional propensity score covariates between unweighted, predefined and HDPS weighted cohort.

2.5.3. COVID-19 death, including triple therapy users

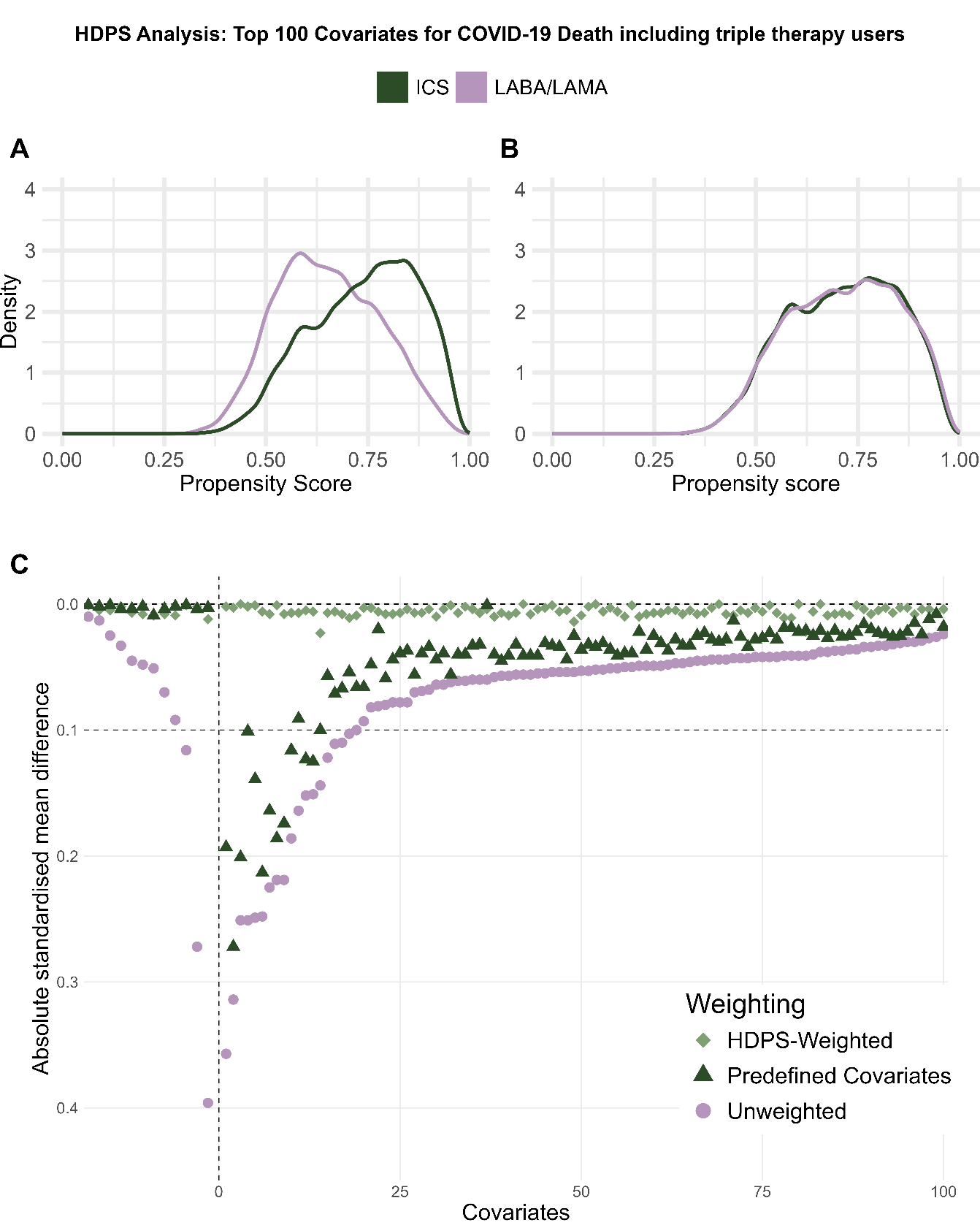

Supplementary Figure 29 Diagnostic plots for high-dimensional propensity score weighted analysis for COVID-19 deaths, including triple therapy users, including the top 100 ranked covariates.

A) high-dimensional propensity score (HDPS) distribution, B) weighted HDPS distribution, C) Comparison of absolute standardised differences in the pre-defined and high-dimensional propensity score covariates between unweighted, predefined and HDPS weighted cohort.

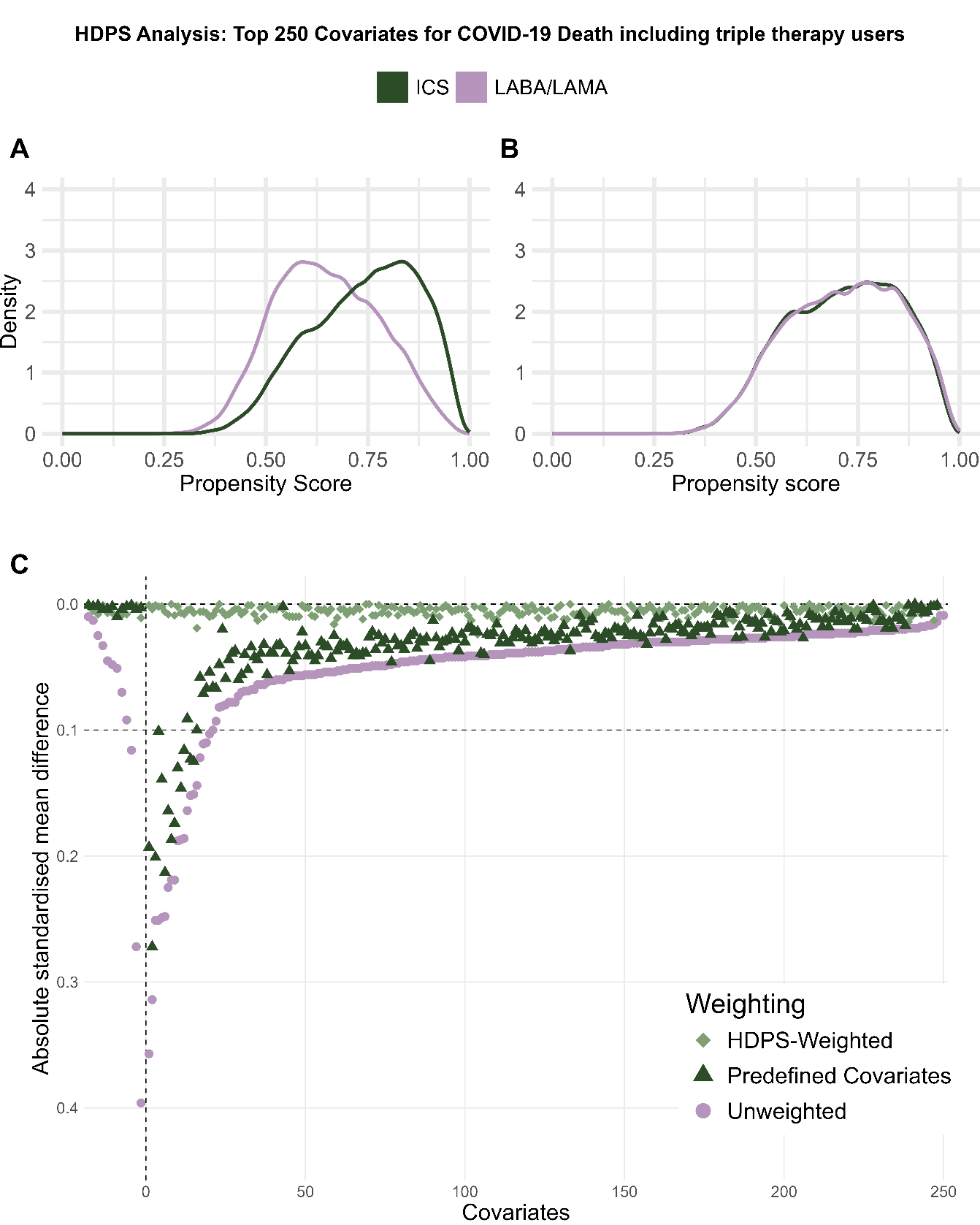

Supplementary Figure 30 Diagnostic plots for high-dimensional propensity score weighted analysis for COVID-19 deaths, including triple therapy users, including the top 250 ranked covariates.

A) high-dimensional propensity score (HDPS) distribution, B) weighted HDPS distribution, C) Comparison of absolute standardised differences in the pre-defined and high-dimensional propensity score covariates between unweighted, predefined and HDPS weighted cohort.

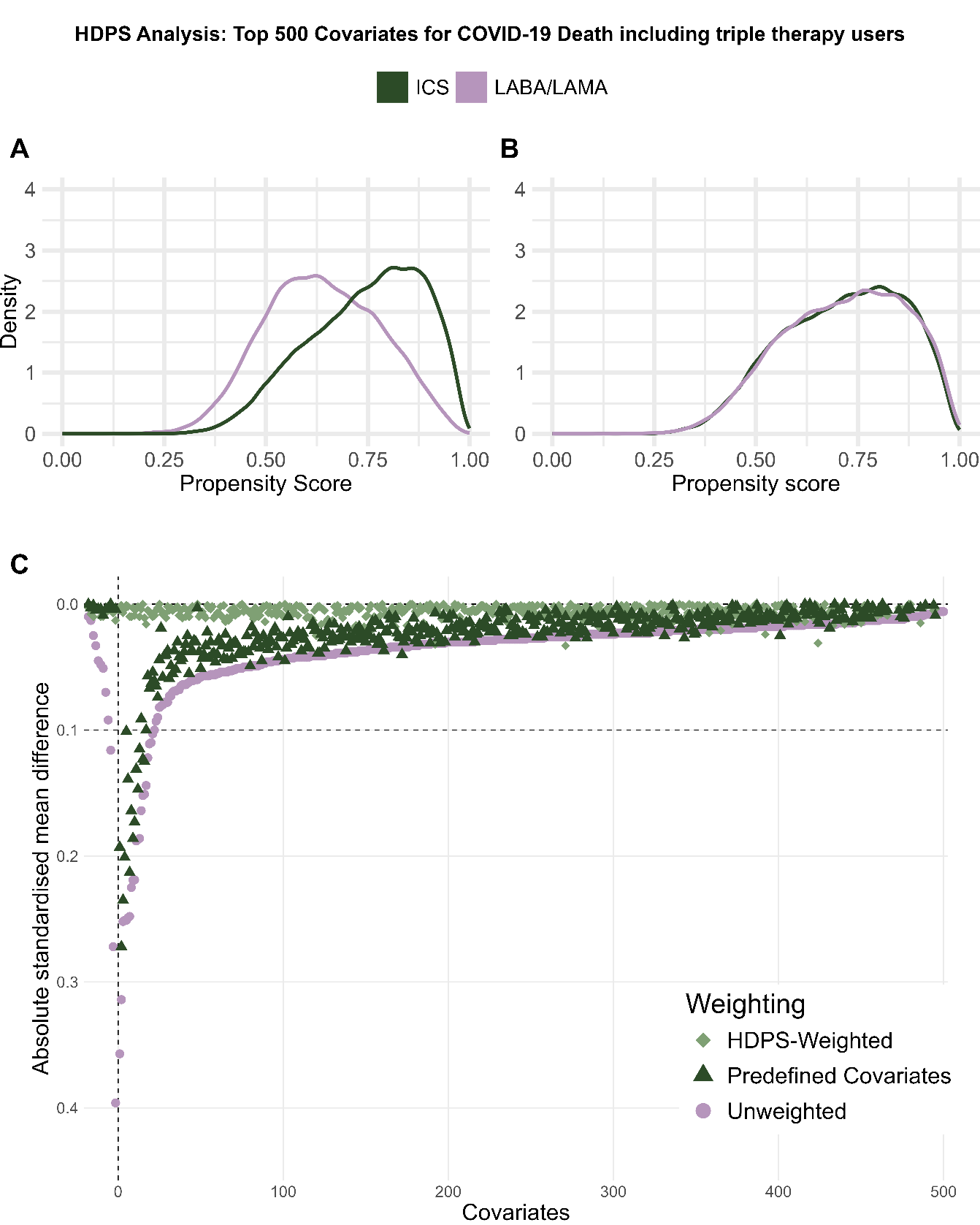

Supplementary Figure 31 Diagnostic plots for high-dimensional propensity score weighted analysis for COVID-19 deaths, including triple therapy users, including the top 500 ranked covariates.

A) high-dimensional propensity score (HDPS) distribution, B) weighted HDPS distribution, C) Comparison of absolute standardised differences in the pre-defined and high-dimensional propensity score covariates between unweighted, predefined and HDPS weighted cohort.

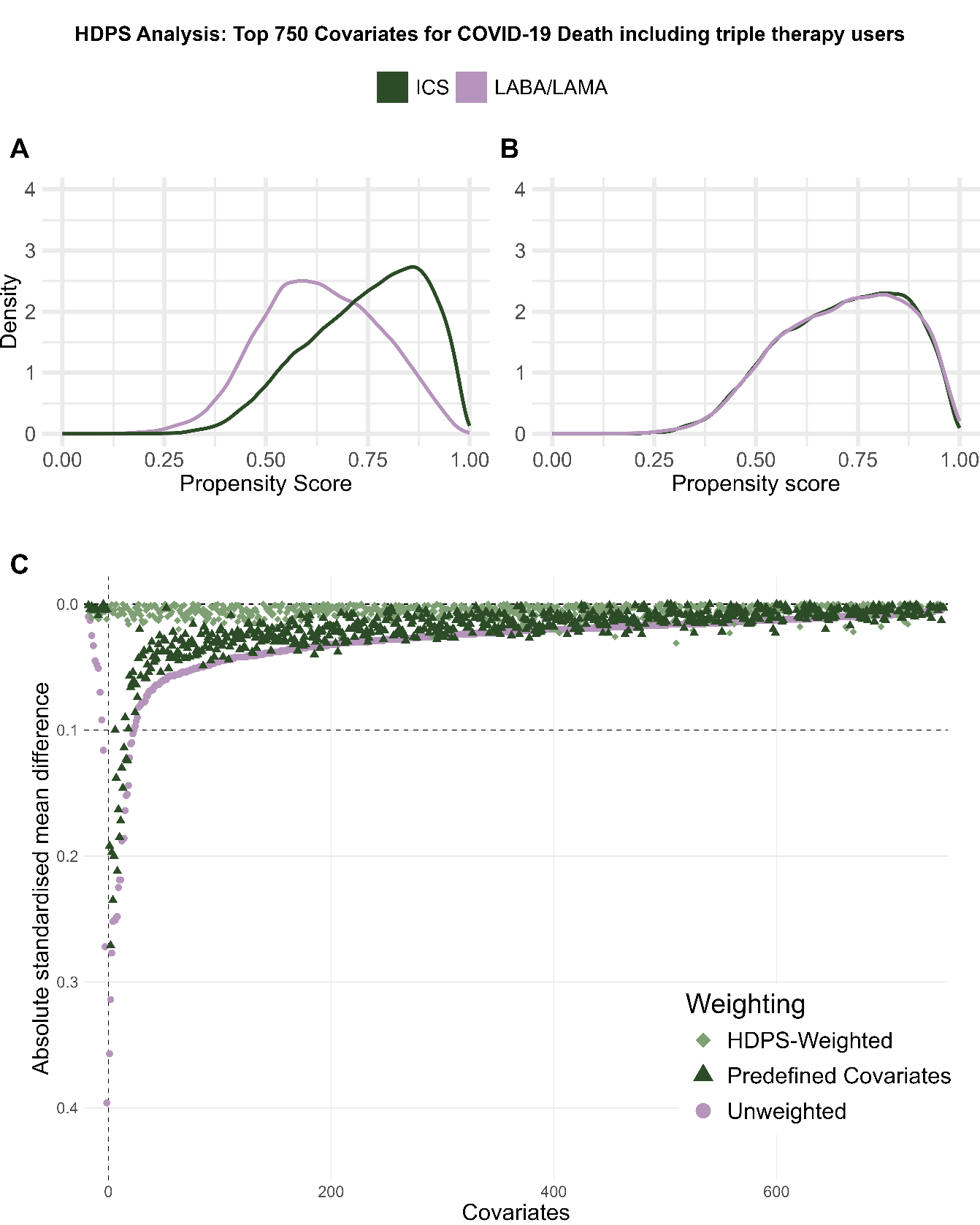

Supplementary Figure 32 Diagnostic plots for high-dimensional propensity score weighted analysis for COVID-19 deaths, including triple therapy users, including the top 750 ranked covariates.

A) high-dimensional propensity score (HDPS) distribution, B) weighted HDPS distribution, C) Comparison of absolute standardised differences in the pre-defined and high-dimensional propensity score covariates between unweighted, predefined and HDPS weighted cohort.

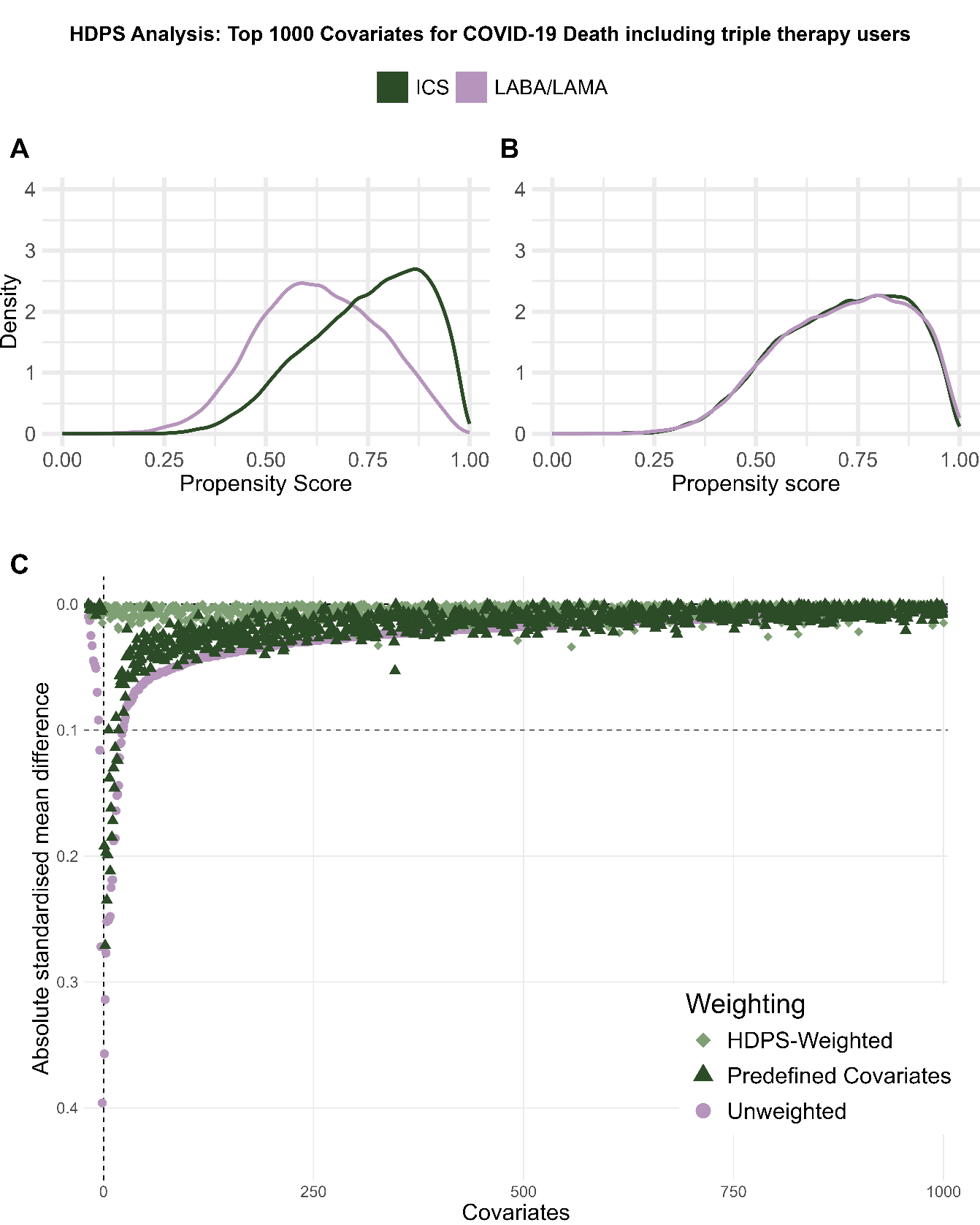

Supplementary Figure 33 Diagnostic plots for high-dimensional propensity score weighted analysis for COVID-19 deaths, including triple therapy users, including the top 1000 ranked covariates.

A) high-dimensional propensity score (HDPS) distribution, B) weighted HDPS distribution, C) Comparison of absolute standardised differences in the pre-defined and high-dimensional propensity score covariates between unweighted, predefined and HDPS weighted cohort.

2.5.4. COVID-19 death, excluding triple therapy users

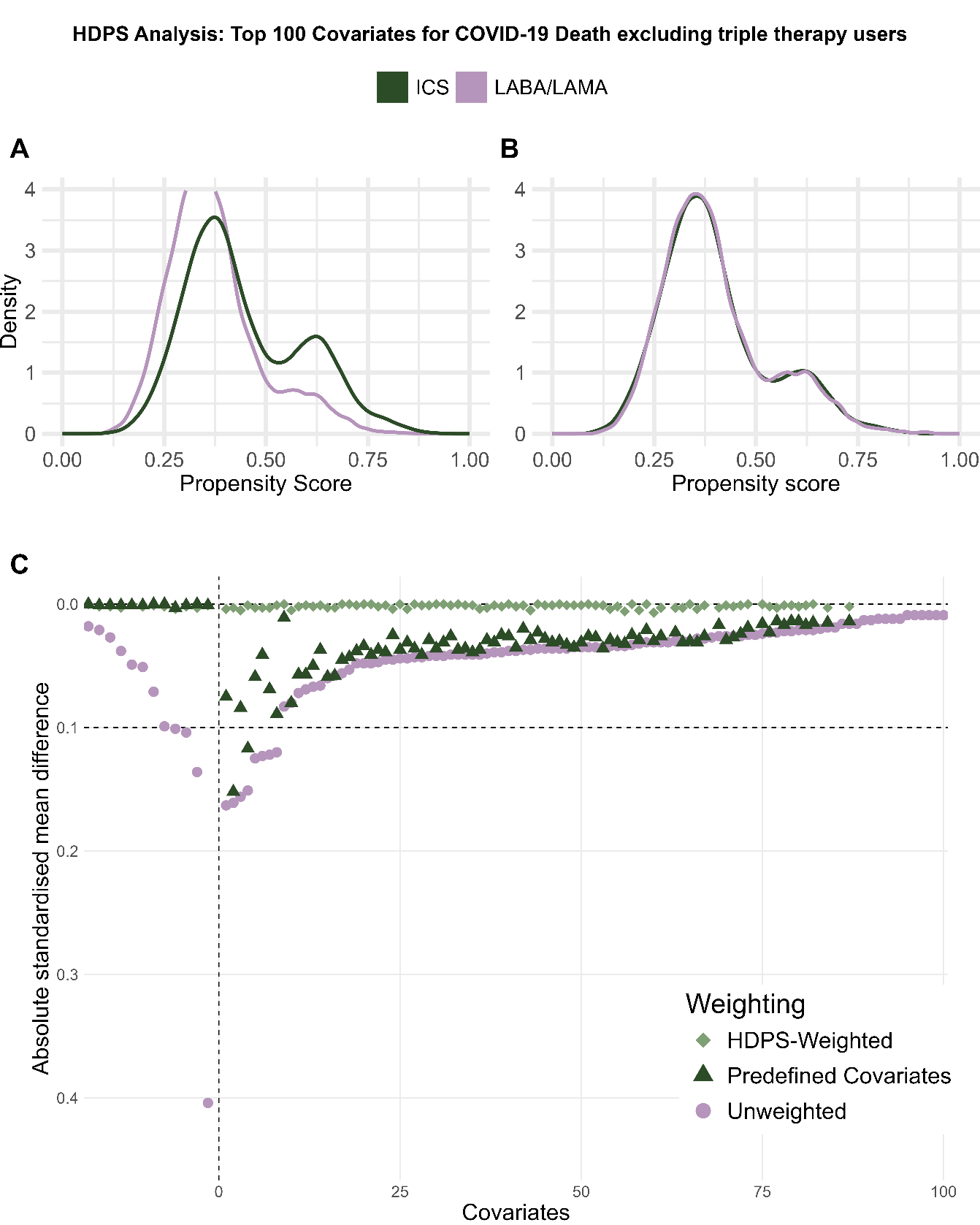

Supplementary Figure 34 Diagnostic plots for high-dimensional propensity score weighted analysis for COVID-19 deaths, excluding triple therapy users, including the top 100 ranked covariates.

A) high-dimensional propensity score (HDPS) distribution, B) weighted HDPS distribution, C) Comparison of absolute standardised differences in the pre-defined and high-dimensional propensity score covariates between unweighted, predefined and HDPS weighted cohort.

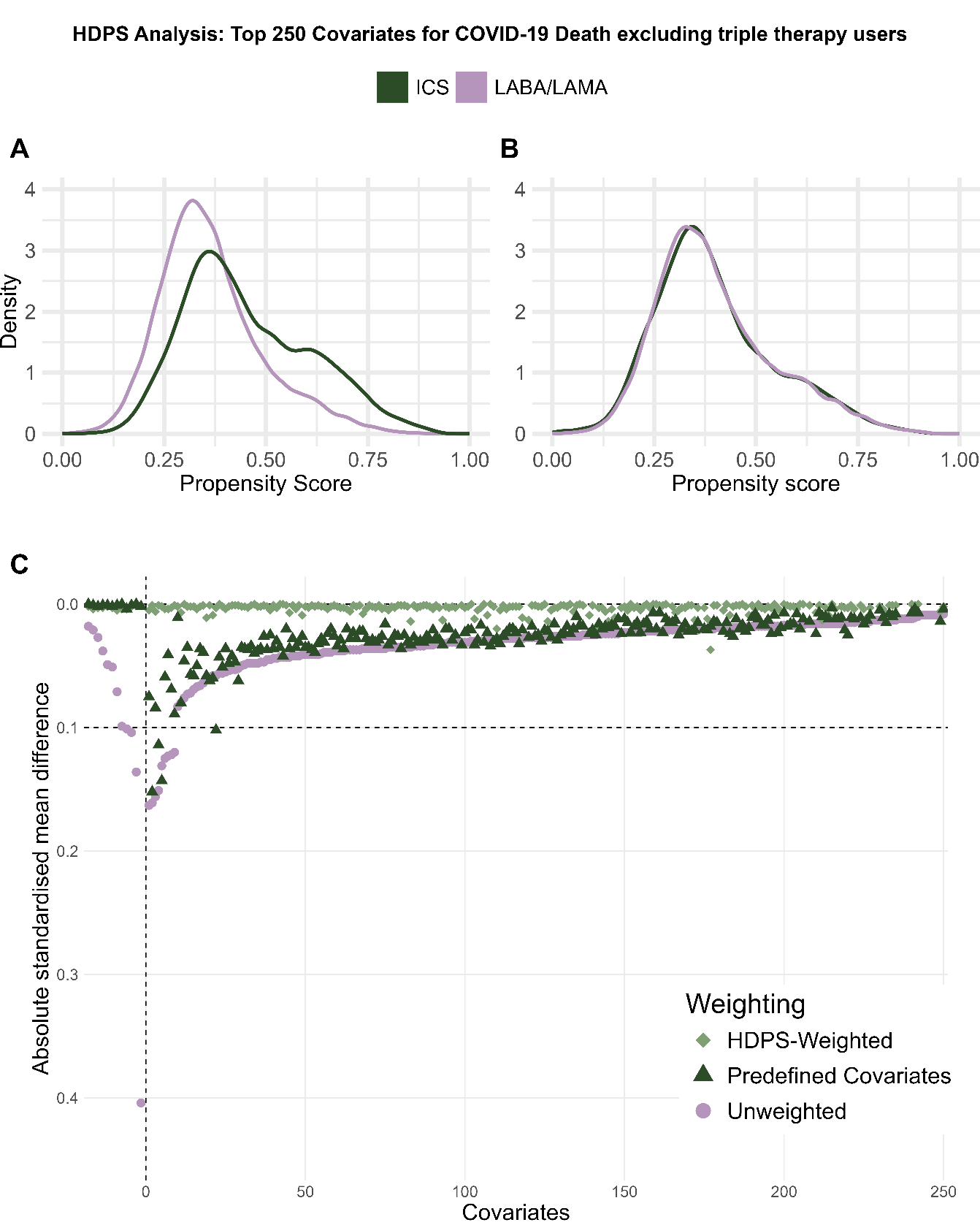

Supplementary Figure 35 Diagnostic plots for high-dimensional propensity score weighted analysis for COVID-19 deaths, excluding triple therapy users, including the top 250 ranked covariates.

A) high-dimensional propensity score (HDPS) distribution, B) weighted HDPS distribution, C) Comparison of absolute standardised differences in the pre-defined and high-dimensional propensity score covariates between unweighted, predefined and HDPS weighted cohort.

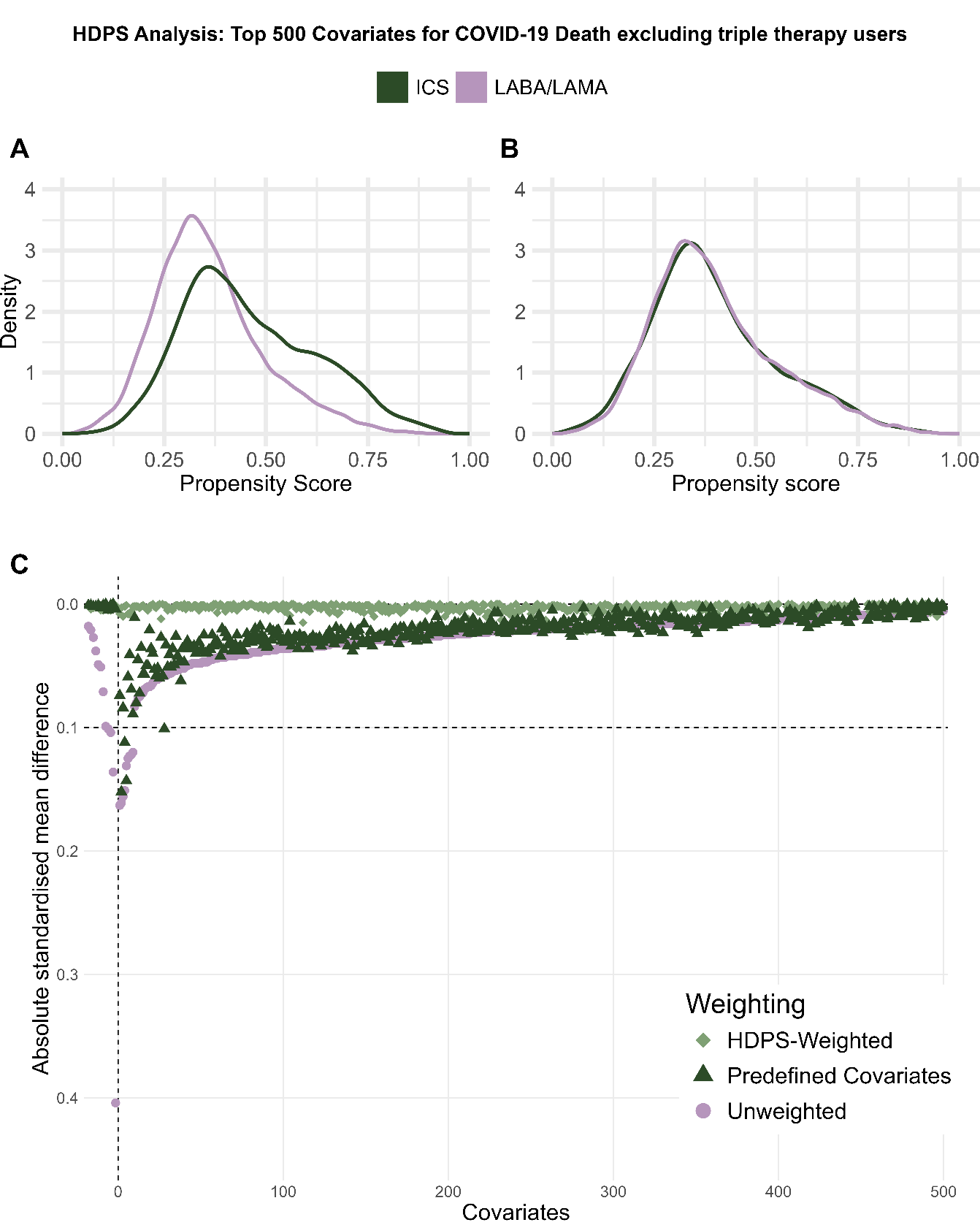

Supplementary Figure 36 Diagnostic plots for high-dimensional propensity score weighted analysis for COVID-19 deaths, excluding triple therapy users, including the top 500 ranked covariates.

A) high-dimensional propensity score (HDPS) distribution, B) weighted HDPS distribution, C) Comparison of absolute standardised differences in the pre-defined and high-dimensional propensity score covariates between unweighted, predefined and HDPS weighted cohort.

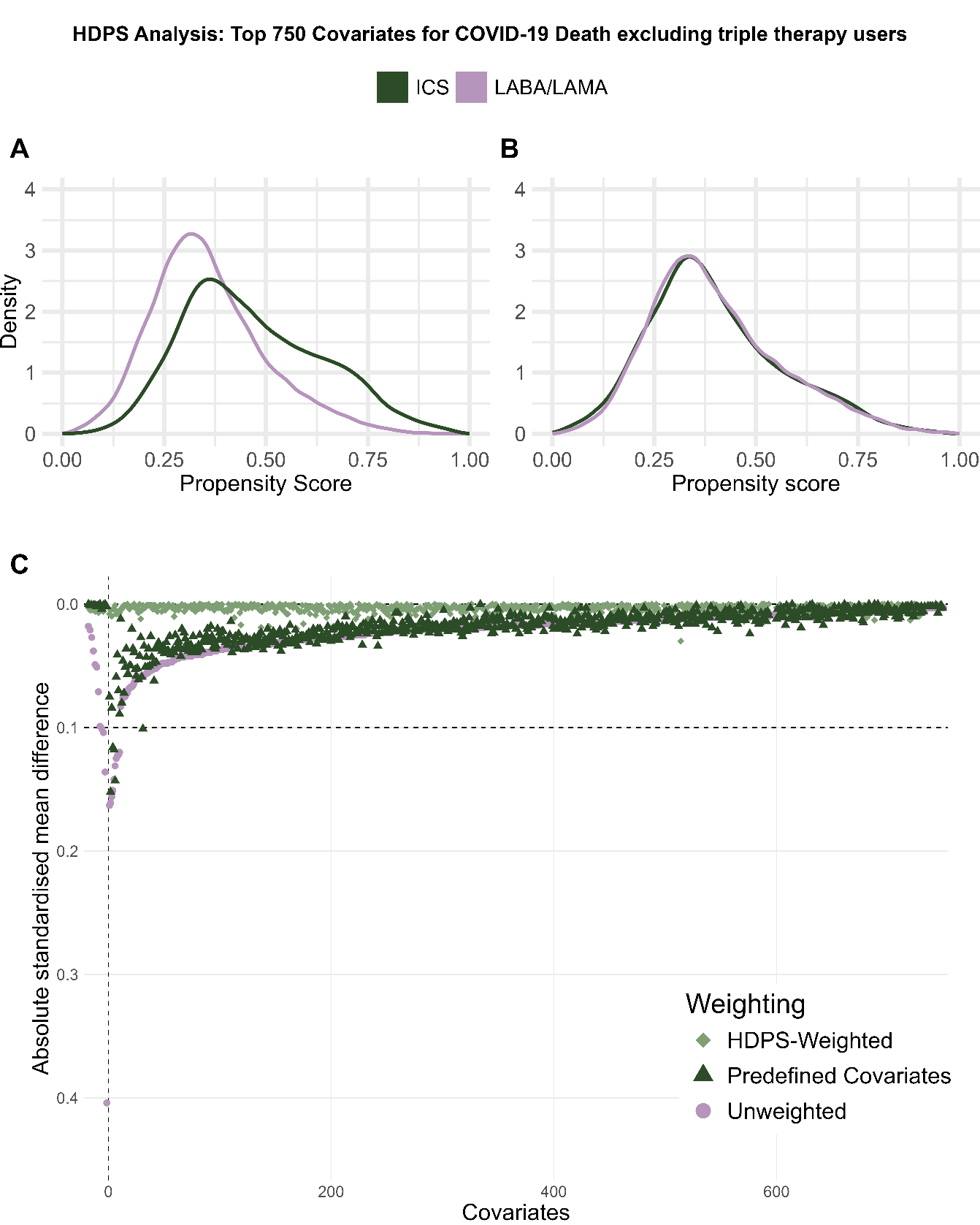

Supplementary Figure 37 Diagnostic plots for high-dimensional propensity score weighted analysis for COVID-19 deaths, excluding triple therapy users, including the top 750 ranked covariates.

A) high-dimensional propensity score (HDPS) distribution, B) weighted HDPS distribution, C) Comparison of absolute standardised differences in the pre-defined and high-dimensional propensity score covariates between unweighted, predefined and HDPS weighted cohort.

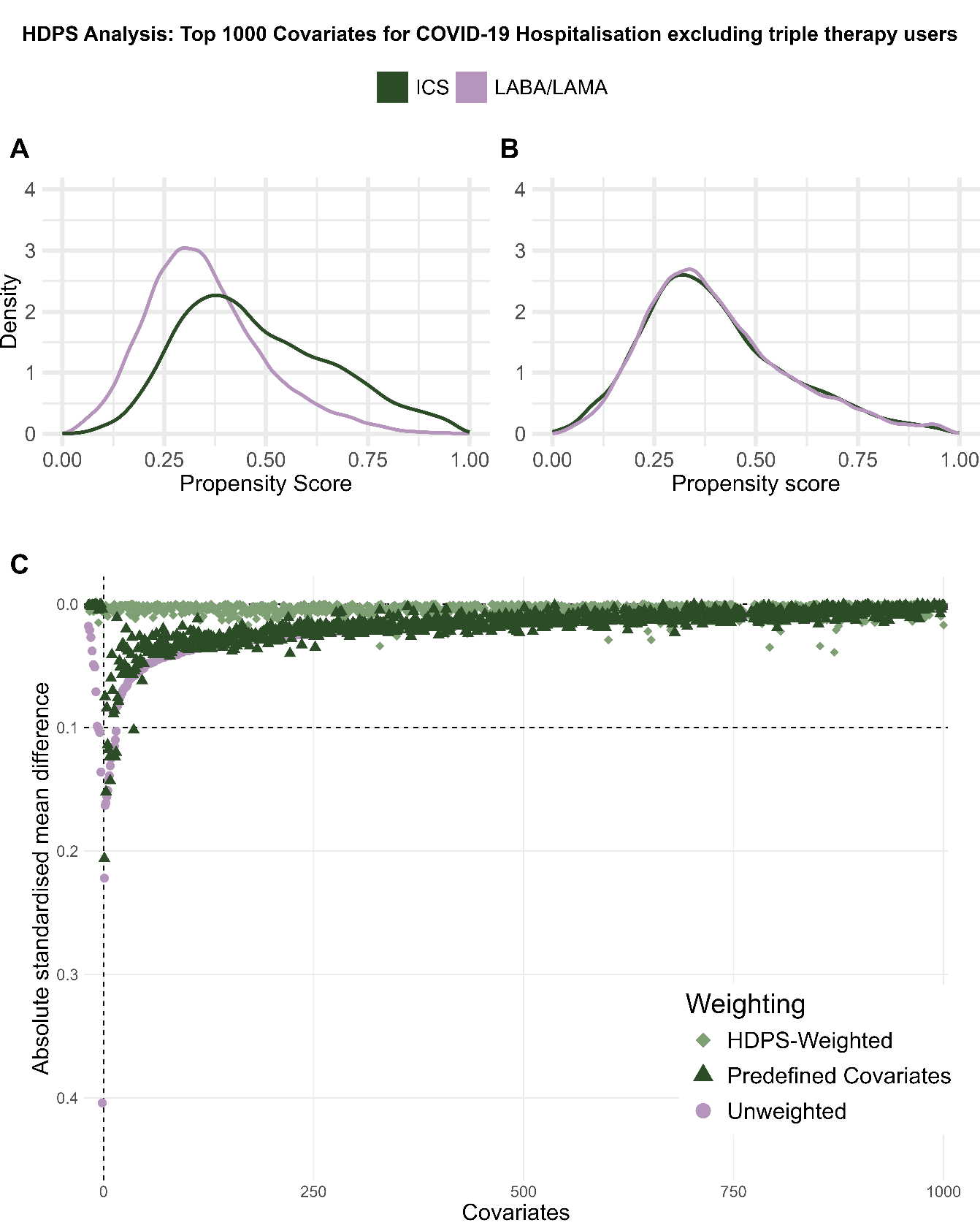

Supplementary Figure 38 Diagnostic plots for high-dimensional propensity score weighted analysis for COVID-19 deaths, excluding triple therapy users, including the top 1000 ranked covariates.

A) high-dimensional propensity score (HDPS) distribution, B) weighted HDPS distribution, C) Comparison of absolute standardised differences in the pre-defined and high-dimensional propensity score covariates between unweighted, predefined and HDPS weighted cohort.

2.6. Cox proportional hazards models
     2.6.1. Kaplan-Meier plots

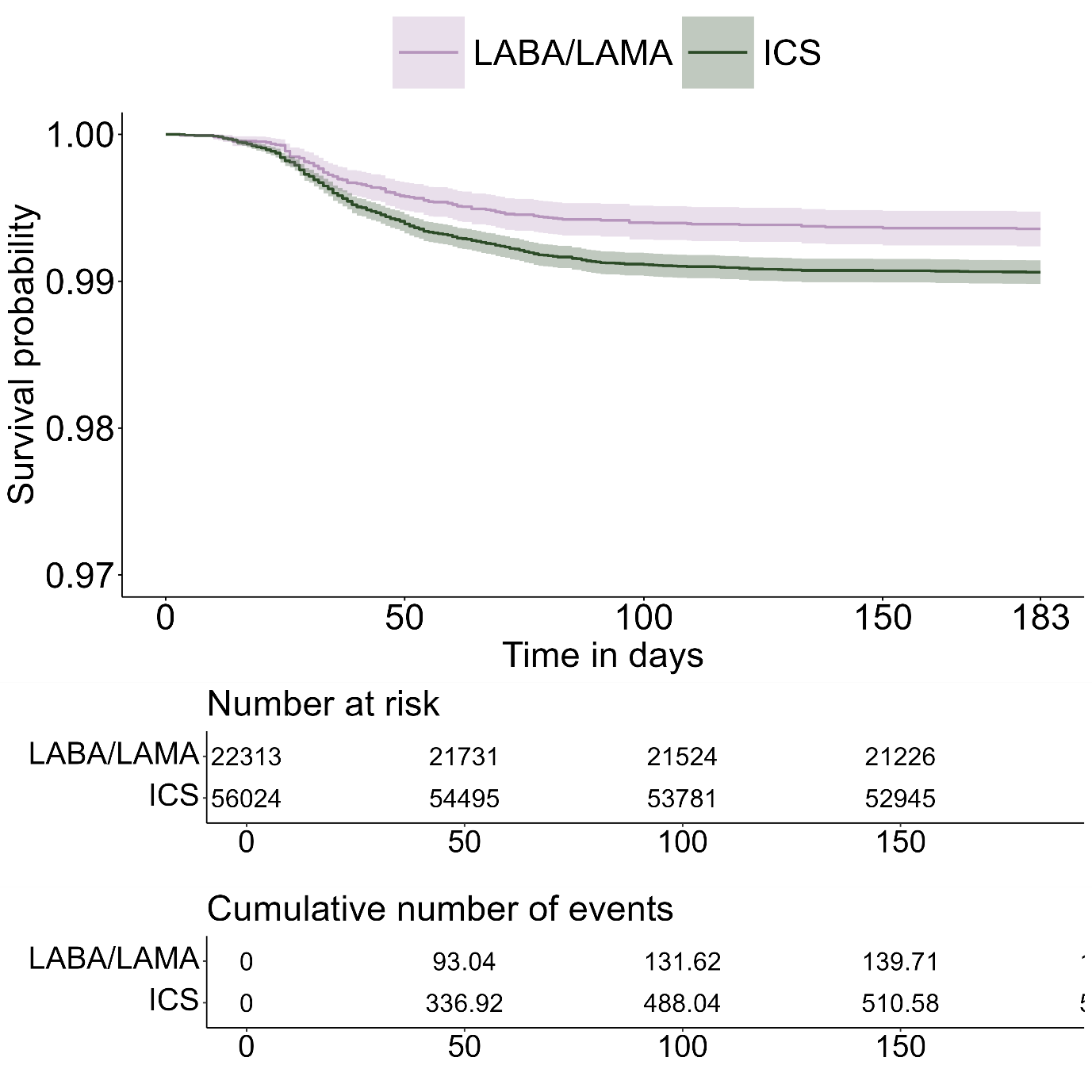

Supplementary Figure 39 Kaplan-Meier curves for COVID-19 hospitalisation, including triple therapy users, weighted using prespecified covariates

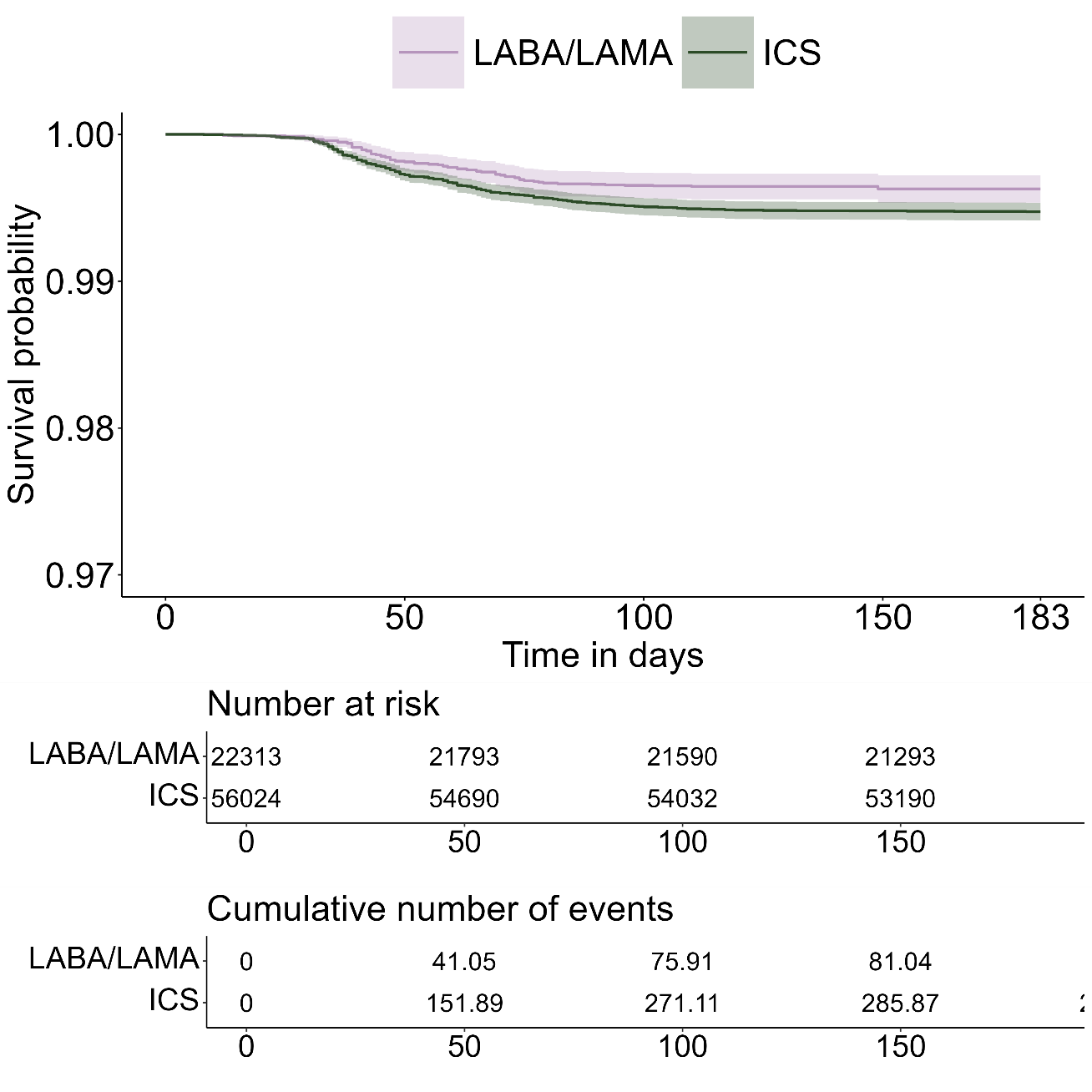

Supplementary Figure 40 Kaplan-Meier curves for COVID-19 death, including triple therapy users, weighted using prespecified covariates

2.7. Logistic regression models

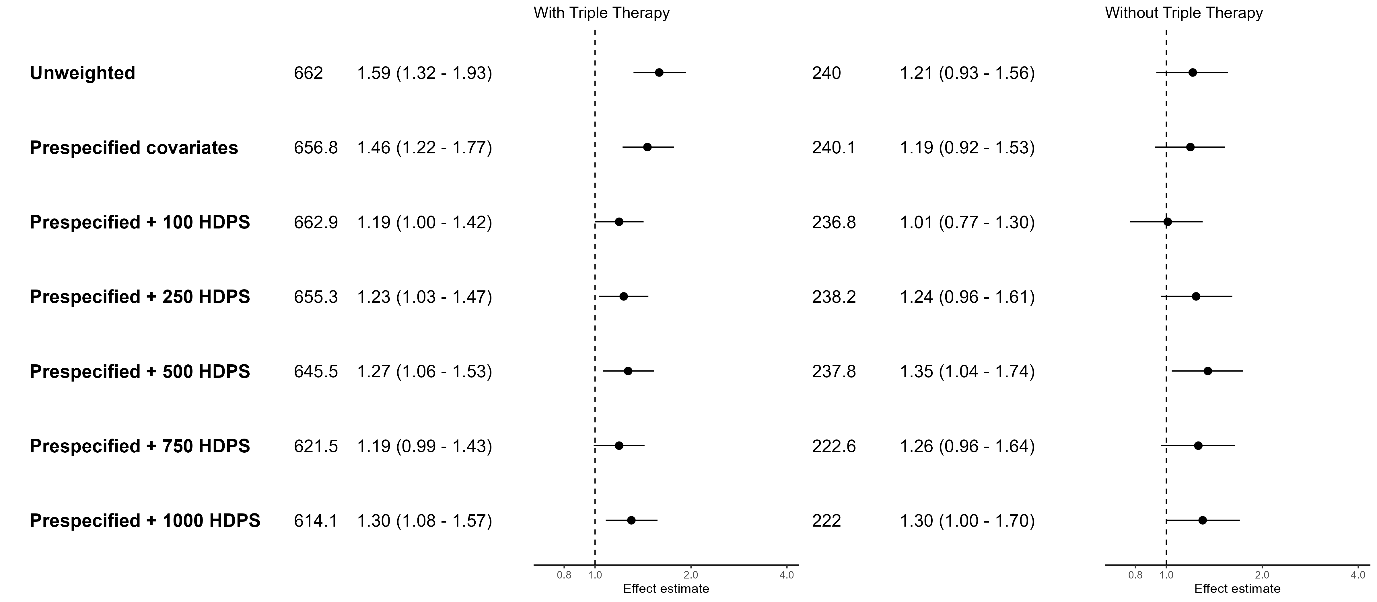

Supplementary Figure 41 Forest plot of odds ratios and 95% confidence intervals for COVID-19 hospitalisations, comparing ICS/LABA (+/- LAMA) users to LABA/LAMA users.

Effect estimates >1 indicate an increased risk in the ICS group.

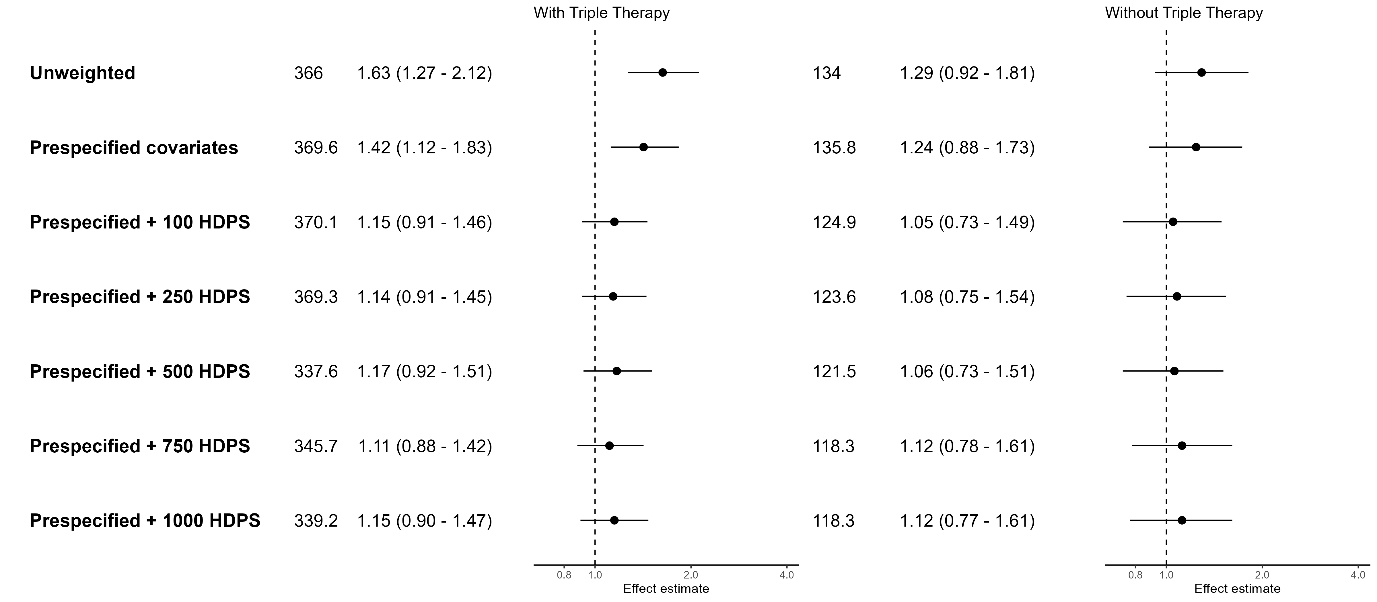

Supplementary Figure 42 Forest plot of odds ratios and 95% confidence intervals for COVID-19 deaths, comparing ICS/LABA (+/- LAMA) users to LABA/LAMA users.

Effect estimates >1 indicate an increased risk in the ICS group.
